# Integrative multi-omics profiling of colorectal cancer from a Hispanic/Latino cohort of patients

**DOI:** 10.1101/2024.11.03.24316599

**Authors:** B Waldrup, F Carranza, Y Jin, Y Amzaleg, M Postel, DW Craig, JD Carpten, B Salhia, CN Ricker, JO Culver, CE Chavez, MC Stern, L Baezconde-Garbanati, HJ Lenz, EI Velazquez-Villarreal

**Affiliations:** City of Hope, Beckman Research Institute, Department of Integrative Translational Sciences, Duarte, CA; University of Southern California, Keck School of Medicine of USC, Department of Translational Genomics, Los Angeles, CA; City of Hope Comprehensive Cancer Center, Duarte, CA; University of Southern California, USC Norris Comprehensive Cancer Center, Los Angeles, CA; University of Southern California, Keck School of Medicine of USC, Division of Medical Oncology, Los Angeles, CA; University of Southern California, Keck School of Medicine of USC, Department of Population and Public Health Sciences, Los Angeles, CA

## Abstract

Colorectal cancer contributes to cancer-related deaths and health disparities in the Hispanic and Latino community. To probe both the biological and genetic bases of the disparities, we characterized features of colorectal cancer in terms of somatic alterations and genetic similarity. Specifically, we conducted a comprehensive genome-scale analysis of 67 Hispanic and Latino samples. We performed DNA exome sequencing for somatic mutations, somatic copy number alterations, and genetic similarity. We also performed RNA sequencing for differential gene expression, cellular pathways, and gene fusions. We analyzed all samples for 22 important CRC gene mutations, 8 gene amplifications, and 25 CRC gene fusions. Then, we compared our data from the Hispanic and Latino samples to publicly available, Non-Hispanic White (NHW) cohorts. According to our analyses, twenty-four percent of colorectal carcinomas were hypermutated when patients were of Peruvians-from-Lima-like (1KG-PEL-like) genetic similarity population from the 1000 genome project. Moreover, most of these cases occurred in patients who were less than fifty years old age at diagnosis. Excluding hypermutated tumors, approximately 55% of colon cancers and 58% of rectum cancers exhibited two similar features: 1) the patterns of genomic alterations; 2) percentage of 1KG-PEL-like. We analyzed all samples -- which had a median 1KG-PEL-like proportion of 55% -- for 22 important CRC gene mutations, 8 gene amplifications, and 25 CRC gene fusions. One notable example of a frequently observed gene mutation was SMAD4. Samples with SMAD4 alterations, which are known to support tumor growth and progression, had the highest 1KG-PEL-like proportion (63%). According to our results from risk association analyses and differential gene expression, SMAD4 alterations were significant when we compared Hispanic and Latino samples to NHW cohorts. Of the 8 drug-targetable amplifications, PIK3CA and PI3K exhibited an average 1KG-PEL-like of over 55%. Of the 25 relevant CRC gene fusions, targetable genes included ALK, FGFR1, RAF1, and PTPRK; PTPRK was observed in a sample with the highest 1KG-PEL-like proportion (95%). Using integrative analysis, we also detected recurrent alterations in the WNT, TGFB, TP53, IGF2/PI3K, and RTK/RAS pathways. Importantly, these alterations mostly occurred in young patients with high 1KG-PEL-like. These findings highlight the potential for tailoring precision medicine therapeutics to an underrepresented population. Our study advances the molecular profiling of CRC in Hispanics and Latinos. In toto, genetic similarity appears to be an important component in understanding colorectal carcinogenesis and has the potential to advance cancer health disparities research.

**Significance:** Our study provides molecular profiles of colorectal cancer in an underserved community, probes the impact of genetic similarity, and sheds light on cancer health disparities.

This study provides a comprehensive, genome-scale analysis of colorectal cancers (CRCs) taken from 67 Hispanic and Latino patients. We performed DNA exome sequencing for somatic mutations, somatic copy number alterations, and genetic similarity. We also performed RNA sequencing for differential gene expression, cellular pathways, and gene fusions. Our data from the Hispanic and Latino samples was compared to publicly available, Non-Hispanic White cohorts. In general, colorectal carcinomas from our cohort presented a high proportion (55%) of Peruvians-from-Lima-like (1KG-PEL-like) population from the 1000 genome project. Moreover, hypermutated colorectal carcinomas (24% of cases) predominantly occurred in individuals with 1KG-PEL-like. Most hypermutated cases occurred in young (<50) patients. We assessed our cohort for 22 important CRC gene mutations, 8 gene amplifications, and 25 CRC gene fusions. We cross-referenced results with percentage of 1KG-PEL-like. For example, frequent mutations were observed in a tumor suppressor called SMAD4; samples with SMAD4 alterations had 63% 1KG-PEL-like. Of the 8 drug-targetable amplifications, PIK3CA and PI3K exhibited an average 1KG-PEL-like of over 55%. Of the 25 CRC gene fusions, targetable genes included ALK, FGFR1, RAF1, and PTPRK. PTPRK was observed in a sample with 95% 1KG-PEL-like proportion. Using integrative analysis, we also detected recurrent alterations in the WNT, TGFB, TP53, IGF2/PI3K, and RTK/RAS pathways. Our study thus advances the molecular profiling of CRC in Hispanics and Latinos; suggests precision medicine therapeutics can be tailored to an underrepresented community; and demonstrates genetic similarity can be an important component in understanding colorectal carcinogenesis.

## INTRODUCTION

Widely adopted screening programs, as well as greatly improved therapies, have decreased the overall burden of colorectal cancer (CRC).[1–4][5] Nevertheless, such progress has not been uniformly observed across all demographic groups.[6–8] Both the largest and fastest-growing minority group in the U.S. – the Hispanic and Latino community – continue to experience disparities in CRC incidence and outcomes.[9] The underlying reasons for these disparities are multifaceted: they involve complex interplay between socioeconomic factors, lifestyle, and genetics.[10] In emerging studies, these disparities appear to be linked to both genetic similarity and the specific molecular characteristics of tumors.[11,12] Although some studies have provided comprehensive, molecular characterizations of CRC, most of this work underrepresents the Hispanic and Latino population. They predominantly use data from Non-Hispanic White individuals. [13,14] They also typically categorize CRC tumors based on their hypermutated and non-hypermutated status. For colorectal malignancies in the Hispanic and Latino community, little to no data is available about somatic mutations and genetic similarity. Altogether, significant gaps remain in our understanding of the molecular landscape of CRC in Hispanic and Latino individuals. Genetic similarity populations from the 1000 genome project of Hispanic and Latino populations plays a crucial role in understanding colorectal cancer tumorigenesis. [15,16,17] A study has shown that genetic similarity influences the molecular characteristics of CRC; genetic similarity affects both the mutation spectrum and the response to treatment.[17] For instance, certain genetic variants prevalent in admixed populations may contribute to higher rates of BRAF mutations in Hispanic/Latino populations. [17] Understanding these genetic differences is vital for addressing health disparities in CRC outcomes. Here, we aim to provide a comprehensive, genome-scale analysis of colorectal cancers (CRCs) taken from 67 Hispanic and Latino patients. We performed DNA exome sequencing for somatic mutations, somatic copy number alterations, and genetic similarity. We also performed RNA sequencing for differential gene expression, cellular pathways, and gene fusions. Our data from the Hispanic and Latino samples was compared to publicly available, Non-Hispanic White cohorts. We looked for several genes and pathways critical for the initiation and progression of CRC: six examples are WNT, RAS-MAPK, PI3K, TGF-β, P53, and DNA mismatch-repair [15,16]. Moreover, we assessed our data for identified recurrently mutated genes and a recurrent chromosomal translocation. The goal is to provide a deeper understanding of CRC disparities at the molecular level and identify potential targeted therapies for precision medicine.

## RESULTS

### Description of the study population

The study population for the Colorectal Cancer (CRC) Moonshot project consisted of 67 Hispanic and Latino patients (**Table 1**). Almost half of the patients (44.8%) were diagnosed under 50 years old. The gender distribution was similar: 53.7% male vs. 46.3% female. Most tumors (80.6%) were microsatellite stable (MSS) as opposed to microsatellite instable (19.4%, MSI). In terms of tumor mutational burden, fewer tumors (23.9%) were classified as hypermutated. The majority of tumors (62.7%) were located in the colon. Pathological staging revealed that 28.4% were in the early stages (Stage I or II), 43.3% were in the late stages (Stage III or IV), and 28.4% had no TNM applicable or an unknown stage.

**Table 1.**
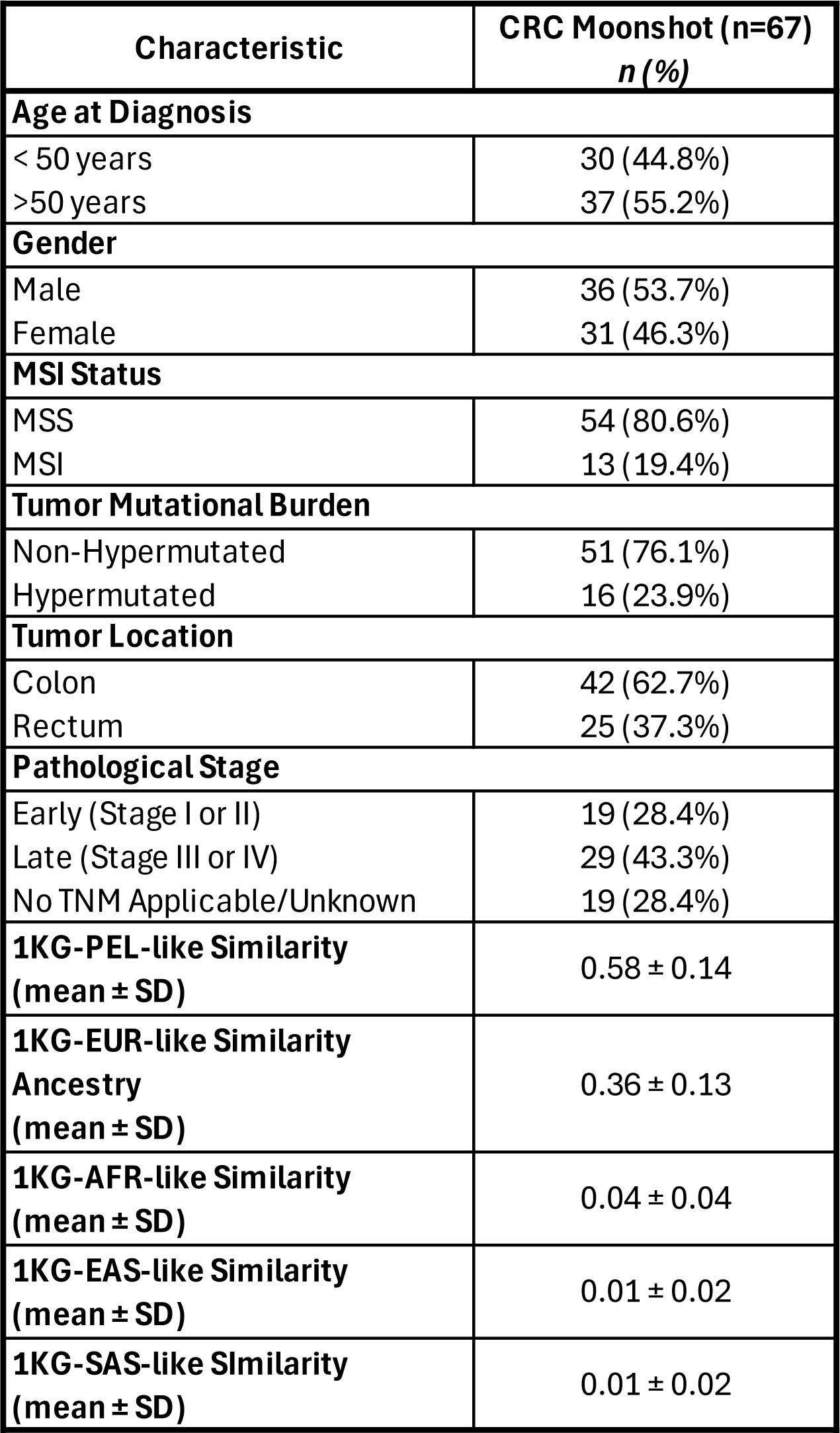
Patient Demographics, Clinical Characteristics, and Genetic Similarity Proportions. This table provides an overview of patient demographics (with race and ethnicity self-reported), clinical characteristics, and genetic similarity proportions relative to the 1000 Genomes Project reference populations. These genetic similarity proportions are categorized as 1KG-PEL-like (Peruvian-in-Lima-like), 1KG-EUR-like (European-like), 1KG-AFR-like (African-like), 1KG-EAS-like (East Asian-like), and 1KG-SAS-like (South Asian-like). The genetic similarity proportions offer insights into the continental origins of the patients’ genomes, enhancing our understanding of their genetic background.

### Whole exome sequence (WES) analysis

To define the genetic similarity, mutational spectrum and chromosomal and sub-chromosomal changes, we performed whole exome sequencing on 67 tumors and normal pairs. Tumor and normal pairs were analyzed using the same platform. WES data summary quality metrics and statistics are provided in **Supplementary Table 1**.

### Genetic similarity and tumor characteristics

Among our cohort, 100% self-reported as Hispanic and Latino. We deduced genetic similarity proportions and admixture for each patient using germline genetic similarity informative markers from WES data (**Fig 1a**). The mean of each genetic similarity population (1000 Genomes Project Peruvian-in-Lima-like-1KG-PEL-like, 1000 Genomes Project European-like-1KG-EUR-like, 1000 Genomes Project African-like-1KG-AFR-like, 1000 Genomes Project South-Asian-like-1KG-SAS-like, 1000 Genomes Project East-Asian-like --1KG-EAS-like) was calculated according to participant status (**Table 1**). CRC cases had 58% 1KG-PEL-like, 36% 1KG-EUR-like, 4% 1KG-AFR-like, 1% 1KG-SAS-like, and 1% 1KG-EAS-like.

**Figure 1:**
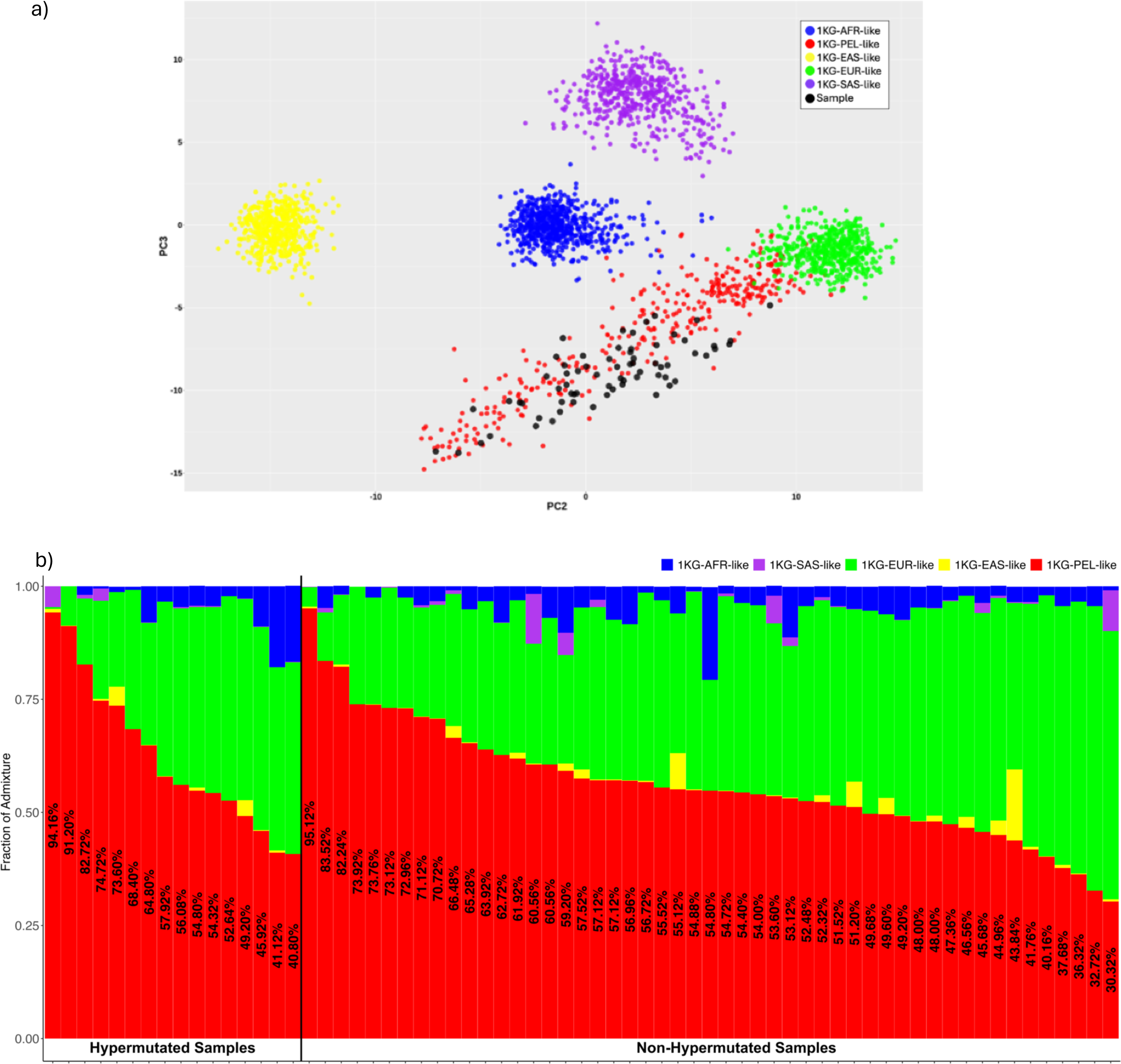
Genetic Similarity and Chromosome-Level Patterns. **a)** Genetic similarity principal component analysis (PCA) plot. This PCA plot illustrates the genetic similarity of our Hispanic/Latino colorectal cancer cohort of 67 individuals, represented by black points, in the context of five super populations: 1000 Genomes Project African-like (1KG-AFR-like, blue), 1000 Genomes Project Peruvian-in-Lima-like (1KG-PEL-like, red), 1000 Genomes Project East-Asian-like (1KG-EAS-like, yellow), 1000 Genomes Project European-like (1KG-EUR-like, green), and 1000 Genomes Project South-Asian-like (1KG-SAS-like, purple). The plot displays the distribution of these samples along two principal components, PC2 and PC3, which capture the majority of the variance in the genetic data. This visualization helps to contextualize the genetic similarity composition of our cohort relative to these major global populations. **b)** This figure illustrates the frequencies in each tumor sample from a cohort of 67 Hispanic/Latino (H/L) colorectal cancer (CRC) patients. The genomes are categorized into five super populations, same as section a. Samples are stratified into hypermutated and non-hypermutated groups, with the proportion of 1KG-PEL-like similarity included.

At the genetic similarity level within our cohort, all participants self-identified as Hispanic and Latino. Among hypermutated tumors, 100% showed a 1KG-PEL-like similarity of over 55% (**Fig. 1b**). In non-hypermutated tumors, 88% also exhibited a higher proportion of 1KG-PEL-like similarity, while 12% showed a 1KG-EUR-like similarity (**Fig. 1b**). These findings indicate a predominance of 1KG-PEL-like similarity in this population, as expected.

We measured the contribution of genetic similarity in the Hispanic and Latino population. Specifically, tumors from both 1KG-PEL-like and 1KG-EUR-like genetic similarity were characterized clinically and at the molecular level (**Supplementary Table 2**). Clinical characteristics included tumor location and tumor stage. Neither were significantly associated with either 1KG-PEL-like or 1KG-EUR-like. We also assessed the following 3 molecular tumor characteristics: microsatellite status, tumor mutational burden, and the mutation status of previously reported CRC genes in our study population. SMAD4-mutated tumors were found more frequently (∼7 times) on patients with 1KG-PEL-like of >55%. However, the exact magnitude of risk increase is uncertain due to the wide confidence interval observed in the data (**Supplementary Table 3**).

### Gene mutations

To define the mutational spectrum, we performed WES on 67 tumors and normal tissue. The somatic mutation rates varied considerably among the samples. Two samples had mutations rates of < 1 per 10^6^ bases; two other samples had mutations rates of > 100 per 10^6^. We separated cases (76%) with a mutation rate of < 9.23 per 10^6^ (median number of total mutations, 91) and those with mutation rates of > 22.31 per 10^6^ (median number of total mutations, 1852). The latter we designated as hypermutated **(Fig. 2a**, **Table 1)**. Compared to a previous study of CRC (N=224)[13], we observed similar percentages of hypermutated and MSI tumors. Our data showed that 24% of the tumors (vs. 13% in [13]) were hypermutated. 81% of our hypermutated tumors (vs. 77% in [13]) exhibited MSI.

**Figure 2:**
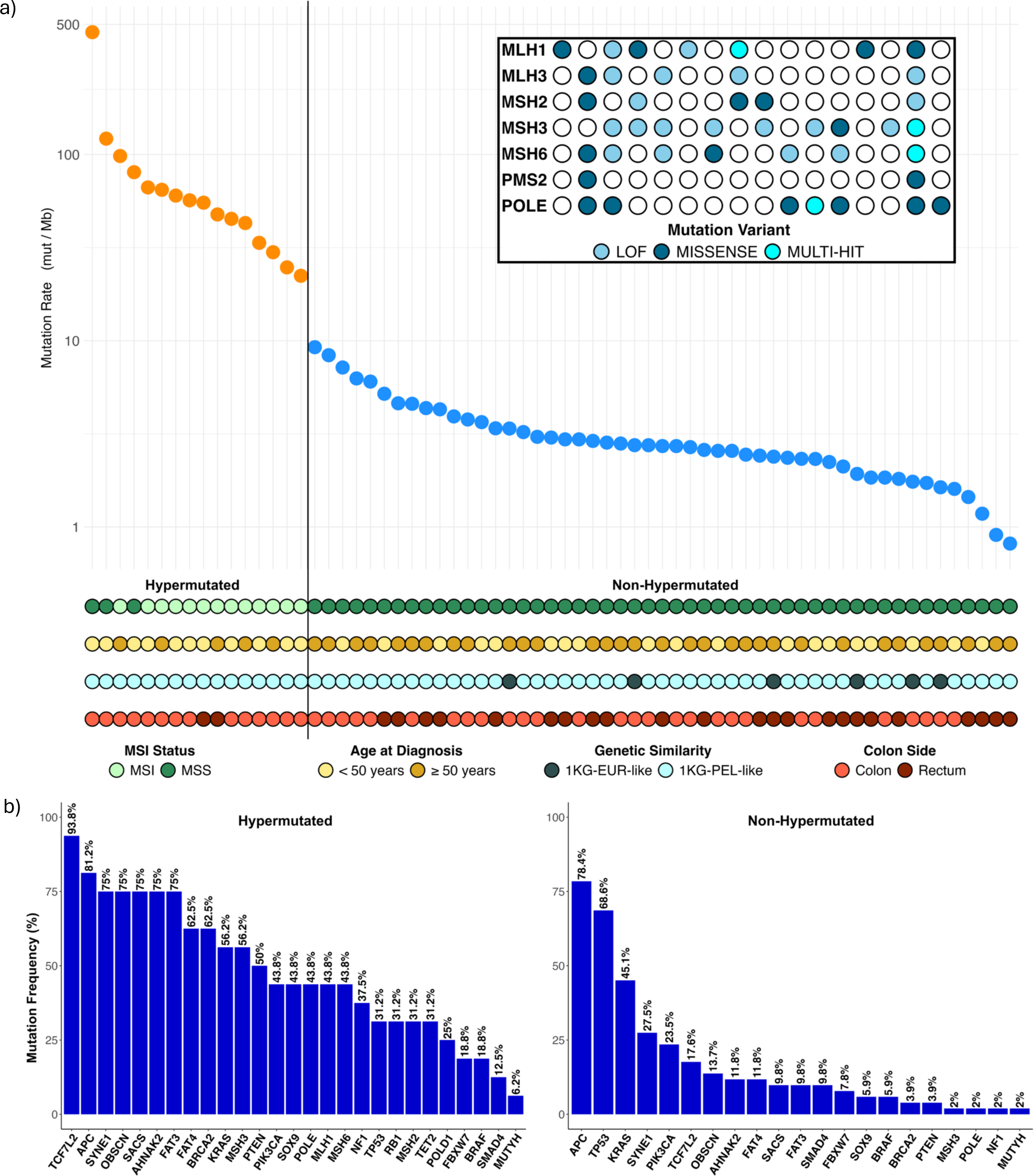
Mutation Frequencies in CRC Tumors from H/L Patients. **a)** This panel shows the mutation frequencies in each of the tumor samples from 67 Hispanic/Latino colorectal cancer patients. The samples are categorized as hypermutated or non-hypermutated. The color codes represent different attributes: light green for MSI, dark green for MSS, light yellow for age at diagnosis < 50 years, brown for age at diagnosis > 50 years, light blue for highest 1KG-PEL-like similarity proportions, dark blue for highest 1KG-EUR-like similarity proportions, light red for colon tumors, and dark red for rectum tumors. The inset highlights mutations in mismatch-repair genes and POLE among the hypermutated samples, with the sample order consistent with the main graph. **b)** This panel depicts the significantly mutated genes in hypermutated and non-hypermutated tumors.

To assess the basis for the considerably different mutation rates —hypermutated (>22.31 per 10^6^) and non-hypermutated (<9.23 per 10^6^)—we evaluated MSI and mutations in the DNA mismatch-repair pathway genes MLH1, MLH3, MSH2, MSH3, MSH6 and PMS2. [18, 19] The following proportions of these 6 genes were observed in our cohort’s 13 hypermutated MSI tumors: 38% MLH1, 31% MLH3, 23% MSH2, 62% MSH3, 46% MSH6, and 8% PMS2. These results agree with results reported by others in reference [13]. Our study also identified similar POLE aberrations in the two samples with highest mutation rates and MSS. **(Fig. 2a)** All hypermutated samples exhibited high 1KG-PEL-like similarity (63% average). The majority were located in the colon (88%); 69% were early onset cases **(Fig. 2a)**.

In both the hypermutated and non-hypermutated cancers (our entire cohort), we identified 27 recurrent somatic mutations in CRC-related genes **(Fig. 2b)**. We observed 27 in hypermutated cancers and 21 in non-hypermutated cancers (**Fig. 2b**, for a complete list see **Supplementary Table 4**). Among the non-hypermutated tumors, the nine most frequently mutated genes were APC, TP53, KRAS, SYNE1, PIK3CA, TCFL2, OBSCN, AHNAK2, and FAT4. As expected, 34% of our samples, which had a higher 1KG-PEL-like (56% average), exhibited mutations in either KRAS (G12D, G12V, G12A, G12C, G13D,) or BRAF (V600E) (**Supplementary Table 5**); these mutations influence diverse mechanisms related to CRC treatment.[20] SACS, FAT3, SMAD4, FBXW7, BRAF and SOX9 were also mutated frequently. Mutations in SOX9, which is crucial for cell differentiation in the intestinal stem cell environment [21,22], led to missense mutations, loss of function (LOF) mutations, multi-hit mutations, and copy number amplifications. The tumor suppressor genes we identified included APC, TP53, BRCA2, PTEN, FBXW7, NF1, and SMAD4. Among these, APC and TP53 showed the highest number of samples with mutations. NF1, PTEN, and SMAD4 were associated with the lowest average age at diagnosis, while PTEN and BRCA2 had the highest average of 1KG-PEL-like proportion (**Supplementary Table 6**). All these genes have been reported in CRC. [13, 23]

In the hypermutated tumors, we frequently observed mutations in TCF7L2, APC, SYNE1, OBSCB, SACS, AHNAK2, FAT3, FAT4, and BRCA2 (**Fig. 2b**). One gene was frequently mutated in non-hypermutated tumors but not in hypermutated tumors: TP53 (66 versus 31%, P<0.05). Twelve genes were more frequently mutated in the hypermutated tumors than in the non-hypermutated tumors: (1) TCF7L2 (94 versus 19% P=7.624e-08), (2) OBSCN (75 versus 13% P=), (3) SACS (75 versus 11% P=2.52e-06), (4) AHNAK2 (75 versus 11% P= 2.52e-06), (5) FAT3 (75 versus 9% P=9.036e-07), (6) FAT4 (63 versus 11% P= 9.104e-05), (7) BRCA2 (63 versus 4% P= 1.278e-06), (8) MSH3 (56 versus 2% P= 1.807e-06), (9) SOX9 (44 versus 6% P= 0.0008421), (10) PTEN (50 versus 4% P= 5.396e-05), (11) POLE (43 versus 2% P= 7.405e-05), and (12) NF1 (38 versus 2% P= 0.000404). Other genes, including MLH1, MSH2, MSH6, POLD1, RB1 and TET2, were mutated recurrently in the hypermutated cancers but not in the non-hypermutated samples. These findings indicate that hypermutated and non-hypermutated tumors progress through different sequences of genetic events. As expected, hypermutated tumors with MSI showed additional differences in the genomic and clinical profiles. All these tumors presented >55% 1KG-PEL-like proportion. Most of them were detected in colon from young patients who were less than 50 years old at diagnosis. (**Fig. 2b**).

### Mutation rate

Colon and rectal tumors are managed differently in the clinic.[24] At the molecular level, we compared 42 colonic and 24 rectal tumors. We performed an initial, integrative analysis of MSI status, somatic copy-number alterations (SCNAs), genetic similarity status, and gene expression profiles. Our approach allowed us to probe potential biological differences between tumors in these two locations. As expected, in non-hypermutated tumors, the overall patterns of copy number changes, 1KG-PEL-like, and mRNA expression were indistinguishable between colon and rectal carcinomas (**Fig. 2a, Supplementary Fig. 1, Supplementary Fig. 2, Supplementary Table 7**). Based on these findings, we combined the two types (colon and rectal) for all subsequent analyses regardless of whether they were hypermutated or non-hypermutated.

### Whole exome sequence comparison with public databases

We compared the frequencies of commonly mutated cancer genes from our CRC cohort to eight publicly available CRC genomic datasets: these included 148 cases from MSK-NatCommun, 1513 cases from MSK-JNCI, 619 cases from DFCI-CellReports, 224 cases from TCGA-Nature, 534 TCGA-PanCancerAtlas, 22 cases from MSK-CancerDiscovery, 471 cases from MSK-Gastroenterology, and 47 cases from MSK-JCO-PrecisOncol (**Table 2**).In nine datasets, including our study (**Supplementary Fig. 3**), the most commonly CRC mutated genes were APC, TP53, and KRAS. In two databases, these three genes consistently ranked among the top four most commonly mutated genes. Five frequently mutated genes in our Hispanic and Latino cohort were relatively distinct from some of the publicly available datasets: SYNE1, TCF7L2, OBSCN, PIK3CA, and SACS. For example, the percentage of SYNE1 mutations in our cohort was 38.8% vs. 27% in DFCI-CellReports, 21% in TCGA-Nature, 27% in TCGA-PanCancerAtlas. Mutations in OBSCN 28.36% were more frequent in the Hispanic and Latino cohort than three databases with ranges from 6.30% to 18.70%. While the percentage of PIK3CA mutations in our cohort (28.36%) was similar to six databases, it differed drastically from one database with a considerably higher percentage (40.90%) and one with a considerably lower percentage (6.10%). The percentage of tumors with mutated SACS in the Hispanic and Latino cohort (26.87%) was higher than three databases (9.80%, 10.80%, and 12.50%).

**Table 2.** Mutation Frequencies Across CRC Studies. This table includes gene names, mutation frequencies from the current Moonshot study (N=67), and frequencies from external studies: MSK-NatCommun (N= 148), MSK-JNCI (N= 1513), DFCI-CellReports (N= 619), TCGA-Nature (N= 224), TCGA-PanCancerAtlas (N= 534), MSK-CancerDiscovery (N= 22), MSK-Gastroenterology (N= 471), MSK-JCO-PrecisOncol (N= 47). The included p-values demonstrate the significance levels when comparing frequencies with our study.

| Moonshot Study (N = 67) |  | MSK-NatCommun (N = 148) |  | MSK-JNCI (N = 1513) |  | DFCI-CellReports (N = 619) |  | TCGA-Nature (N = 224) |  |
| --- | --- | --- | --- | --- | --- | --- | --- | --- | --- |
| Gene | Mutated n (%) | Mutated n (%) | p-value | Mutated n (%) | p-value | Mutated n (%) | p-value | Mutated n (%) | p-value |
| APC | 54 (80.60%) | 35 (23.60%) | **1.333E-14** | 1145 (75.50%) | 0.4227 | 361 (58.30%) | **6.46E-04** | 168 (75.00%) | 0.4346 |
| TP53 | 40 (59.70%) | 131 (88.50%) | **3.05E-06** | 1107 (73.00%) | **0.0245** | 320 (51.70%) | 0.2637 | 121 (54.00%) | 0.4959 |
| KRAS | 34 (50.75%) | 41 (27.70%) | **0.0018** | 666 (43.90%) | 0.3303 | 173 (27.90%) | **1.98E-04** | 94 (42.00%) | 0.2583 |
| SYNE1 | 26 (38.81%) | 0 (NA) | 1 | 0 (NA) | 1 | 167 (27.00%) | 0.0572 | 47 (21.00%) | **0.0052** |
| TCF7L2 | 25 (37.31%) | 0 (NA) | 1 | 196 (14.20%) | **6.725E-07** | 44 (7.10%) | **3.089E-14** | 27 (12.10%) | **5.28E-06** |
| OBSCN | 19 (28.36%) | 0 (NA) | 1 | 0 (NA) | 1 | 116 (18.70%) | 0.0856 | 14 (6.30%) | **1.69E-06** |
| PIK3CA | 19 (28.36%) | 9 (6.10%) | **1.90E-05** | 298 (19.70%) | 0.1128 | 132 (21.30%) | 0.2441 | 45 (20.10%) | 0.2057 |
| SACS | 18 (26.87%) | 0 (NA) | 1 | 0 (NA) | 1 | 67 (10.80%) | **0.0003** | 22 (9.80%) | **8.01E-04** |
| AHNAK2 | 18 (26.87%) | 0 (NA) | 1 | 0 (NA) | 1 | 106 (17.10%) | 0.0717 | 21 (9.40%) | **4.96E-04** |
| FAT3 | 17 (25.37%) | 9 (6.10%) | **1.49E-04** | 0 (NA) | 1 | 90 (14.50%) | **0.0320** | 16 (7.10%) | **9.26E-05** |
| FAT4 | 16 (23.88%) | 0 (NA) | 1 | 0 (NA) | 1 | 109 (17.60%) | 0.2728 | 39 (17.40%) | 0.3130 |
| BRCA2 | 12 (17.91%) | 5 (3.40%) | **7.13E-04** | 83 (5.50%) | **8.45E-05** | 42 (6.80%) | **0.0029** | 10 (4.50%) | **7.01E-04** |
| MSH3 | 10 (14.93%) | 2 (2.20%) | **0.0042** | 28 (4.70%) | **0.0018** | 20 (3.20%) | **3.60E-05** | 15 (6.70%) | **0.0460** |
| SOX9 | 10 (14.93%) | 4 (2.70%) | **0.0016** | 154 (10.20%) | 0.2945 | 62 (10.00%) | 0.3004 | 37 (4.50%) | 0.9032 |
| PTEN | 10 (14.93%) | 4 (2.70%) | **0.0016** | 88 (5.80%) | **0.0056** | 51 (8.20%) | 0.1095 | 8 (3.60%) | **1.96E-03** |
| POLE | 8 (11.94%) | 4 (2.70%) | **0.0101** | 85 (5.60%) | 0.0585 | 47 (7.60%) | 0.3135 | 15 (6.70%) | 0.2552 |
| FBXW7 | 7 (10.45%) | 20 (13.50%) | 0.6847 | 205 (13.50%) | 0.5893 | 86 (13.90%) | 0.5520 | 37 (16.50%) | 0.3066 |
| NF1 | 7 (10.45%) | 9 (6.10%) | 0.3957 | 76 (5.00%) | 0.0943 | 45 (7.30%) | 0.4898 | 8 (3.60%) | 0.0551 |
| MLH1 | 7 (10.45%) | 1 (0.70%) | **0.0014** | 41 (2.70%) | **0.0011** | 24 (3.90%) | **0.0316** | 6 (2.70%) | **0.0181** |
| MSH6 | 7 (10.45%) | 3 (2.00%) | **0.0114** | 59 (3.90%) | **0.0206** | 27 (4.40%) | 0.0596 | 15 (6.70%) | 0.4498 |
| SMAD4 | 7 (10.45%) | 19 (12.80%) | 0.7856 | 238 (15.70%) | 0.3219 | 73 (11.80%) | 0.9001 | 26 (11.60%) | 0.9657 |
| BRAF | 6 (8.96%) | 6 (4.10%) | 0.2588 | 135 (8.90%) | 1 | 127 (20.50%) | **0.0348** | 21 (9.40%) | 1 |
| RB1 | 5 (7.46%) | 1 (0.70%) | **0.0120** | 36 (2.40%) | **0.0298** | 22 (3.60%) | 0.2179 | 5 (2.20%) | 0.0930 |
| MSH2 | 5 (7.46%) | 2 (1.40%) | **0.0316** | 38 (2.50%) | **0.0396** | 12 (1.90%) | **0.0188** | 5 (2.20%) | 0.0930 |
| TET2 | 5 (7.46%) | 3 (2.00%) | 0.1113 | 53 (3.50%) | 0.1742 | 11 (1.80%) | **0.0123** | 4 (1.80%) | **0.0328** |
| POLD1 | 4 (5.97%) | 3 (2.10%) | 0.2098 | 45 (3.30%) | 0.2818 | 34 (5.50%) | 0.7805 | 5 (2.20%) | 0.2188 |
| MUTYH | 2 (2.99%) | 3 (2.00%) | 0.6480 | 16 (1.10%) | 0.1751 | 8 (1.30%) | 0.2545 | 2 (0.90%) | 0.2280 |
| Moonshot Study (N = 67) |  | TCGA-PanCancerAtlas (N = 534) |  | MSK-CancerDiscovery (N = 22) |  | MSK-Gastroenterology (N = 471) |  | MSK-JCO-PrecisOncol (N = 47) |  |
| Gene | Mutated n (%) | Mutated n (%) | p-value | Mutated n (%) | p-value | Mutated n (%) | p-value | Mutated n (%) | p-value |
| APC | 54 (80.60%) | 387 (72.50%) | 0.2035 | 21 (95.50%) | 0.174 | 325 (69.00%) | 0.07137 | 39 (83.00%) | 0.9382 |
| TP53 | 40 (59.70%) | 314 (58.80%) | 0.9925 | 21 (95.50%) | **0.00124** | 345 (73.20%) | **0.03115** | 20 (42.60%) | 0.1064 |
| KRAS | 34 (50.75%) | 218 (40.80%) | 0.1556 | 22 (100.00%) | **6.33E-06** | 238 (50.50%) | 1 | 45 (95.70%) | **8.217E-08** |
| SYNE1 | 26 (38.81%) | 144 (27.00%) | 0.05952 | 0 (NA) | 1 | 0 (NA) | 1 | 0 (NA) | 1 |
| TCF7L2 | 25 (37.31%) | 58 (10.90%) | **1.018E-08** | 11 (50.00%) | 0.1797 | 36 (9.20%) | **1.251E-09** | 17 (37.80%) | 1 |
| OBSCN | 19 (28.36%) | 99 (18.50%) | 0.08115 | 0 (NA) | 1 | 0 (NA) | 1 | 0 (NA) | 1 |
| PIK3CA | 19 (28.36%) | 147 (27.50%) | 1 | 9 (40.90%) | 0.4035 | 88 (18.70%) | 0.09051 | 17 (36.20%) | 0.4974 |
| SACS | 18 (26.87%) | 67 (12.50%) | **0.002841** | 0 (NA) | 1 | 0 (NA) | 1 | 0 (NA) | 1 |
| AHNAK2 | 18 (26.87%) | 65 (12.20%) | **0.001947** | 0 (NA) | 1 | 0 (NA) | 1 | 0 (NA) | 1 |
| FAT3 | 17 (25.37%) | 81 (15.20%) | **0.05048** | 0 (NA) | 1 | 0 (NA) | 1 | 0 (NA) | 1 |
| FAT4 | 16 (23.88%) | 124 (23.20%) | **0.05296** | 0 (NA) | 1 | 0 (NA) | 1 | 0 (NA) | 1 |
| BRCA2 | 12 (17.91%) | 38 (7.10%) | **0.005419** | 3 (13.60%) | 1 | 21 (4.50%) | **5.78E-05** | 7 (14.90%) | 0.8004 |
| MSH3 | 10 (14.93%) | 11 (2.10%) | **2.02E-05** | 3 (13.60%) | 1 | 6 (3.90%) | **0.009045** | 5 (12.50%) | 0.9507 |
| SOX9 | 10 (14.93%) | 64 (12.00%) | 0.6219 | 8 (36.40%) | 0.06202 | 39 (8.30%) | 0.1231 | 11 (23.40%) | 0.3659 |
| PTEN | 10 (14.93%) | 34 (6.40%) | **0.02224** | 4 (18.20%) | 0.7407 | 23 (4.90%) | **0.003355** | 10 (21.30%) | 0.4558 |
| POLE | 8 (11.94%) | 36 (6.70%) | 0.1967 | 7 (31.80%) | 0.06683 | 17 (3.60%) | **0.006509** | 10 (21.30%) | 0.278 |
| FBXW7 | 7 (10.45%) | 90 (16.90%) | 0.2431 | 8 (36.40%) | **0.0128** | 51 (10.80%) | 1 | 13 (27.70%) | **0.03331** |
| NF1 | 7 (10.45%) | 31 (5.80%) | 0.228 | 5 (22.70%) | 0.2698 | 24 (5.10%) | 0.1392 | 6 (12.80%) | 0.933 |
| MLH1 | 7 (10.45%) | 22 (4.10%) | **0.04817** | 10 (45.50%) | **9.28E-04** | 9 (1.90%) | **5.31E-04** | 2 (4.30%) | 0.3032 |
| MSH6 | 7 (10.45%) | 24 (4.50%) | 0.07446 | 8 (36.40%) | **0.0128** | 6 (1.30%) | **3.32E-05** | 8 (17.00%) | 0.4589 |
| SMAD4 | 7 (10.45%) | 68 (12.70%) | 0.7356 | 14 (63.60%) | **1.52E-06** | 69 (14.60%) | 0.4614 | 11 (23.40%) | 0.1081 |
| BRAF | 6 (8.96%) | 62 (11.60%) | 0.6584 | 7 (31.80%) | **0.008478** | 58 (12.30%) | 0.5532 | 4 (8.50%) | 1 |
| RB1 | 5 (7.46%) | 14 (2.60%) | 0.07767 | 8 (36.40%) | **0.000785** | 7 (1.50%) | **0.007873** | 3 (6.40%) | 1 |
| MSH2 | 5 (7.46%) | 21 (3.90%) | 0.3076 | 5 (22.70%) | 0.1146 | 4 (0.80%) | **0.002197** | 3 (6.40%) | 1 |
| TET2 | 5 (7.46%) | 23 (4.30%) | 0.3966 | 12 (54.50%) | **5.07E-06** | 13 (2.80%) | 0.1011 | 3 (6.40%) | 1 |
| POLD1 | 4 (5.97%) | 29 (5.40%) | 0.7777 | 5 (22.70%) | **0.03801** | 7 (1.80%) | 0.06152 | 5 (11.10%) | 0.4804 |
| MUTYH | 2 (2.99%) | 7 (1.30%) | 0.2648 | 3 (13.60%) | 0.09447 | 3 (0.60%) | 0.1192 | 1 (2.10%) | 1 |

Of 19 genes we studied in our cohort, sixteen of them exhibited similar percentages of mutations to at least one of the eight public databases. Three genes had slightly higher frequencies than all the referenced databases (**Table 2**). Two genes exhibited high, statistically significant differences with respect to the largest dataset (MSK-JNCI): TCF7L2 (p-values of 6.725E-07) and BRCA2 (p-value of 8.45E-05). Both of these genes had higher mutational frequencies in our cohort (**Table 2**). Other genes with notable variant differences with respect to the largest dataset include TP53, MSH3, PTEN, MLH1, MSH6, RB1 and MSH2 (**Table 2**). Ethnicity information was reported only in the TCGA-PanCancer Atlas: an extremely small percentage of samples (0.8%) of the samples identified as Hispanic and Latino.

Furthermore, we compared our CRC cohort to two previous studies—Innocenti et al. (2023, *JCO*) and Nunes et al. (2024, *Nature*)—as detailed in Supplementary Table 8. Notably, one of these studies highlighted significant differences in the frequency of APC and KRAS mutations when compared to our cohort, particularly among non-Hispanic White (NHW) individuals. In our Hispanic and Latino cohort, seventeen genes were frequently mutated and showed relative distinction from one or both studies. These genes include: TCF7L2, PIK3CA, FAT3, BRCA2, SOX9, PTEN, POLE, NF1, MLH1, MSH6, SMAD4, BRAF, RB1, MSH2, TET2, POLD1, and MUTYH. This distinct mutational landscape underscores the potential genetic differences in CRC across diverse populations.

We additionally assessed somatic mutations in CRC cases from the AACR Project GENIE dataset (**Table 3**). This dataset includes self-identified race and ethnicity information. We combined cases from our cohort, TCGA-PanCancer and AACR Genie, while stratifying by self-reported race. This combined dataset included a total of 140 (9%) patients who self-identified as Hispanic and Latino. The remaining 1487 (91%) cases self-identified as Non-Hispanic White (**Table 3**). Based upon this large cohort, which was race stratified by self-identification, we identified twelve statistically significant differences in the frequency of commonly mutated genes: SYNE1, TCF7L2, OBSCN, PIK3CA, SACS, AHNAK2, FAT3, FAT4, BRCA2, MSH3, POLE and RB1 (**Table 3**).

**Table 3:**
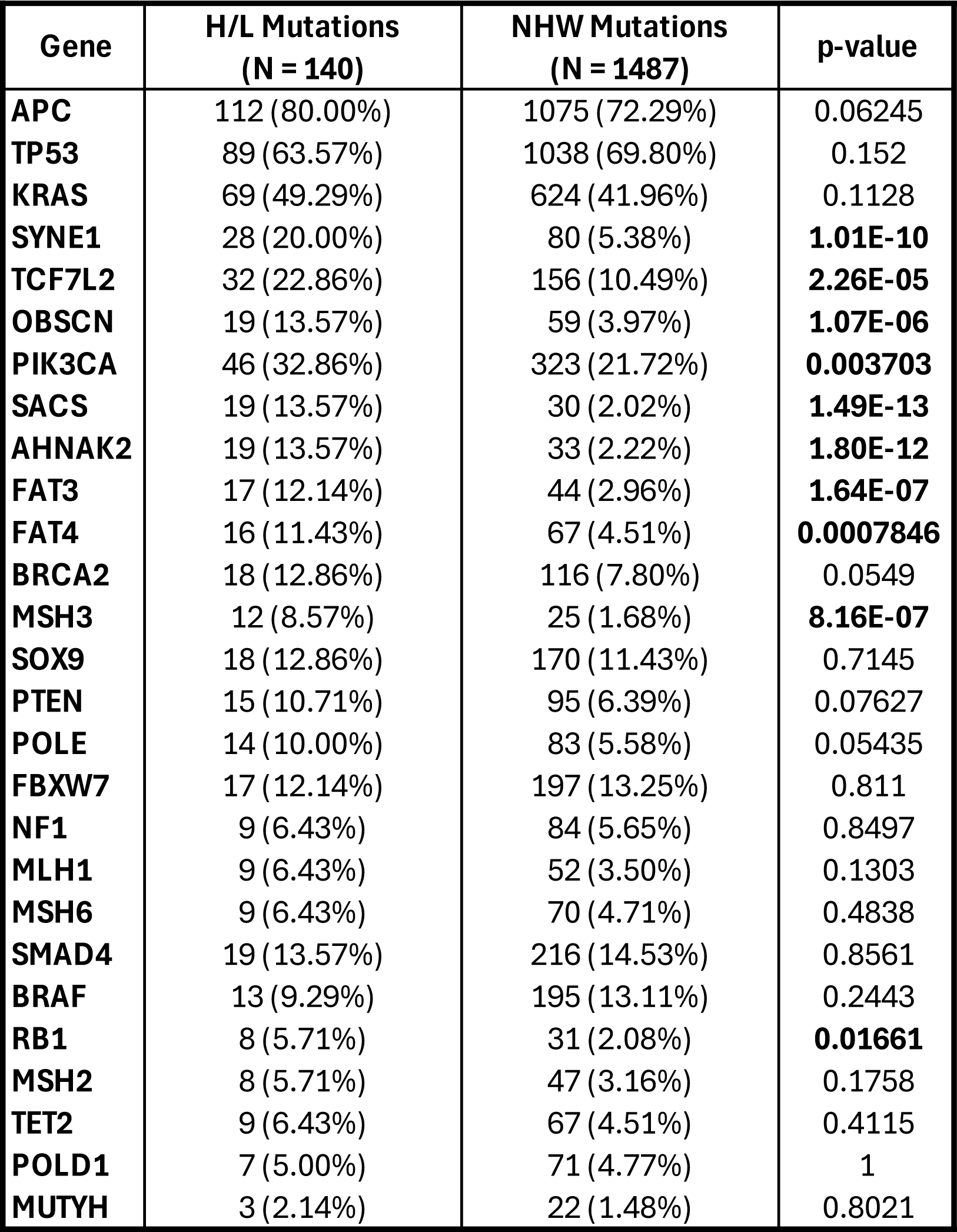
Mutation Frequencies in Hispanic/Latino and Non-Hispanic White Cases. This table compares mutation frequencies in Hispanic/Latino (H/L) and Non-Hispanic White (NHW) cases across three datasets: our Moonshot study, the AACR Project GENIE dataset, and the TCGA-PanCancer project. It lists gene names along with their respective mutation frequencies as percentages for each racial group. The final column presents p-values, indicating the statistical significance of the observed differences in mutation frequencies between H/L and NHW patients.

### Chromosomal and sub-chromosomal changes

In total, 67 tumors were profiled by Whole Exome Sequencing (WES) for Somatic Copy-Number Alterations (SCNAs, **Fig. 3**). As previously reported in CRC studies, the hypermutated tumors had far fewer SCNAs (**Supplementary Fig. 4**) than non-hypermutated tumors. However, hypermutated MSI samples had more SCNAs than MSS samples (**Supplementary Fig. 5**). In addition, samples with ≤55% 1KG-PEL-like proportion had slightly more amplifications than samples with >55 1KG-PEL-like proportion (**Supplementary Fig. 6**). No significant differences were detected between non-hypermutated colon and rectum (**Supplementary Fig. 1**).

**Figure 3:**
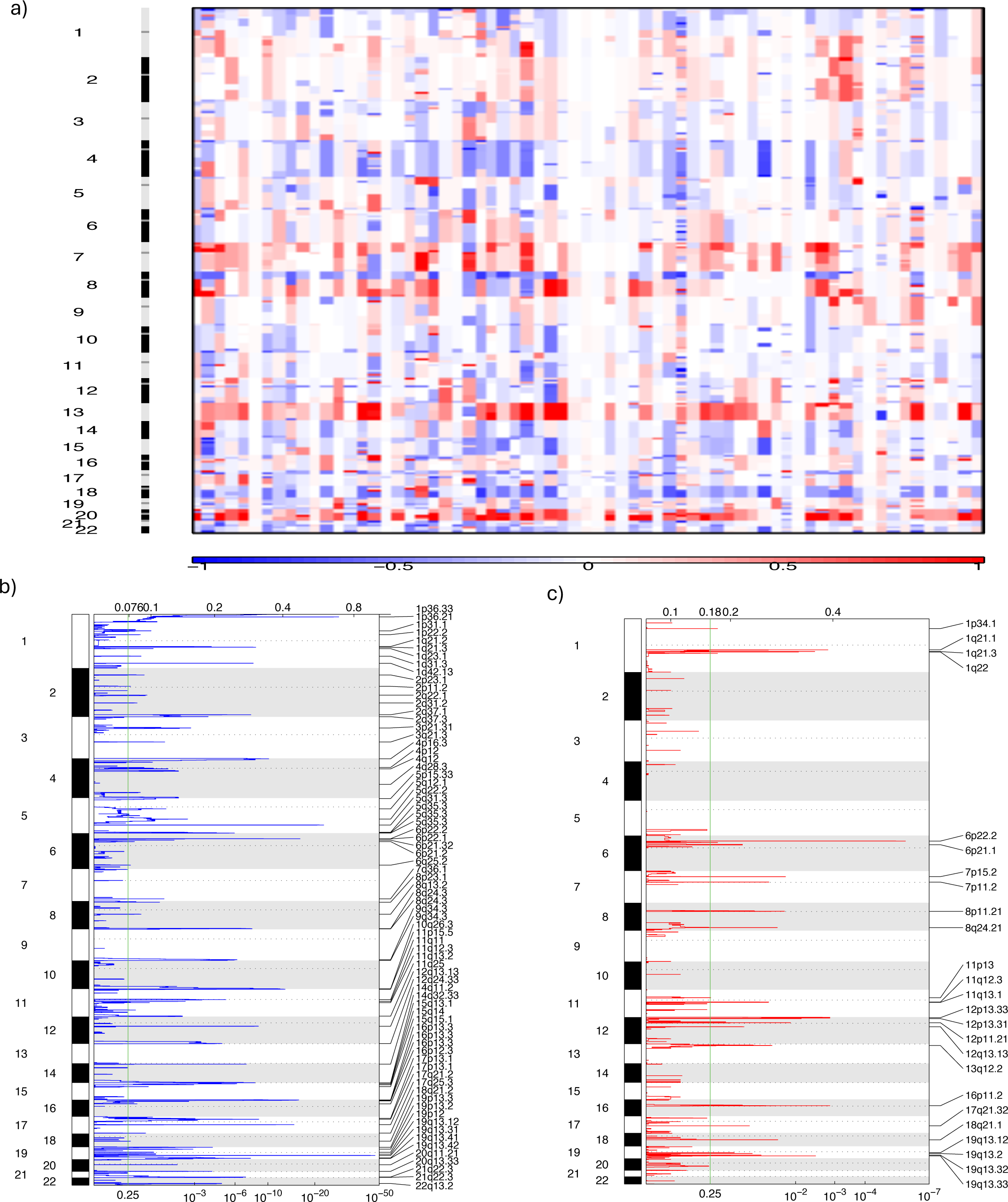
Somatic Copy Number Alterations (SCNAs) Analysis of Genomic Changes in 67 Hispanic/Latino Colorectal Cancer Patients. **a)** Heatmap of SCNAs: This panel displays SCNAs at both chromosomal and sub-chromosomal levels across 67 tumor samples from Hispanic/Latino colorectal cancer patients. Deletions (losses) are indicated in blue, while insertions (gains) are shown in red. **b)** Focal Deletions: This figure highlights the focal deletions identified across all 67 tumor samples. **c)** Focal Amplifications: This figure presents the focal amplifications observed in the same set of tumor samples.

We used the Genomic Identification of Significant Targets in Cancer (GISTIC) algorithm and manually curated data to identify probable gene targets of focal alterations.[25] Most tumors had several, previously well-defined arm-level changes: gains of 1q (93%), 7p (96%) and q (92.5%), 8p (87%) and q (97%), 12q (87%), 13q (96%), 19q (93%), and 20p (97%) and q (100%) (**Supplementary Table 9**). Significantly amplified chromosomes arms were 1q (including OBSCN) in 2% of the samples, 13q (including SACS, BRCA2 and RB1) in 10%, and 14q (including AHNAK2) in 3% (**Supplementary Fig. 8**). CCND1 and OBSCN had also deletions as described below. Other significantly deleted chromosome arms were 2p (80%), 13p (99%), 14p (100%), 15p (100%), 20q (80%), 22p (96%) (**Supplementary Table 9**). Compared to previously reported[13], CRC-related deletion of chromosome arms, our samples exhibited smaller amounts of deletions in 1p (28%), 4q (55%), 5q (30%), 8p (48%), 14q (40%), 15q (48%), 20p (78%) and 22q (49%) (**Supplementary Table 9**). Other significantly deleted chromosomes arms were 1p and q (including MUTYH in 2% and OBSCN in 7% of the samples), 4q (including FAT4 in 2% and FBXW7 in 2%), 5q (including APC in 5% and MSH3 in 2%), 10q (including PTEN in 6%), 11q (including FAT3 in 3%), 12q (including POLE in 3%), 14q (including AHNAK2 in 2%), 18q (including SMAD4 in 12%), and 19q (including POLD1 in 2%) (**Supplementary Fig. 7**).

Our data from the 67 Hispanic and Latino patients had 66 recurrent deletion peaks (**Supplementary Table 10a**). The following four categories of genes were affected by focal deletions in our samples: (1) RBFOX1 and WWOX;[13] (2) the tumor-suppressor genes SMAD4[26] and APC;[27] (3) the mismatch repair genes MSH2 and MSH6;[28] and (4) a ubiquitin ligase called FBXW7, which targets oncoproteins for degradation (alterations can lead to the accumulation of oncogenic proteins).[29] Our data set also had a significant focal deletion of 1q42.13, which spanned two genes. This included OBSCN, which was also frequently mutated in our data set. Other significant focal deletions were 4q28.3, 12q24.33, 4q28.3, and 19q13.41: they involved the genes FAT4, POLE, TET2A and POLD1. Four significant focal deletions 6q25.2, 2p23.1, 2p11.2, and 18q21.2 affect genes detected in our gene fusion analyses: PTPRK, ALK, EML4 and RSPO3. While these appear to be non-significant, we observed 25 regions of significant focal amplification of 6p22.2; this included two genes – HIST1H4H [28] and BTN3A2 [30,31] – which play crucial roles in both histone modification patterns and regulation of immune responses (**Supplementary Table 10b**). Some of these overlapped with extensive gains on chromosome arms and featured notable peaks at 8q24.21 and 7p11.2. Importantly, these peaks encompass both the key oncogene MYC [32] and the well-known, druggable target gene EGFR.[33]

### Transcriptomics Analysis

We performed RNA sequencing on the same 67 tumors to define gene expressions profiles and fusions and pathways.

### Transcriptomic comparisons with public databases: Hispanics and Latinos compared to Non-Hispanic Whites

Compared to colorectal tumors of Non-Hispanic White populations, tumors from our Hispanic and Latino patients had unique gene expression patterns and cellular pathways. Specifically, we conducted a differential gene expression and pathway analysis using RNA sequencing (**Fig. 4, Supplementary Fig. 8**). We compared gene expression profiling in 67 tumors from the Hispanic and Latino patients to 67 tumors from Non-Hispanic White patients. Data from Non-Hispanic White patients were taken from the TCGA-COAD and TCGA-READ projects. We matched tumors from the different populations using parameters shown in Table 1: tumor location, MSI status, age at onset, and TNM classification (**Supplementary Table 11**). A comparison of case matched ratios revealed at least 10 statistically significant CRC related genes at p-value of <0.0001. These genes were as follows: PIK3CA, MUTYH, PMS2P3, POLE, PTEN, SYNE1, POLD1, SMAD4, HFM1, and FAT3. Pathway analysis identified 10 cellular pathways related to colorectal cancer. These pathways were related to dysregulation of GTPases, microRNA (miRNA), non-coding RNAs, post-transcriptional mechanisms, apoptotic signaling, mRNA degradation, ribosome biogenesis, protein-protein interactions, and mitochondrial membrane organization. Notably, activation of GTPase activity, such as RAS and RHO family members, is commonly associated with colorectal cancer.

**Figure 4:**
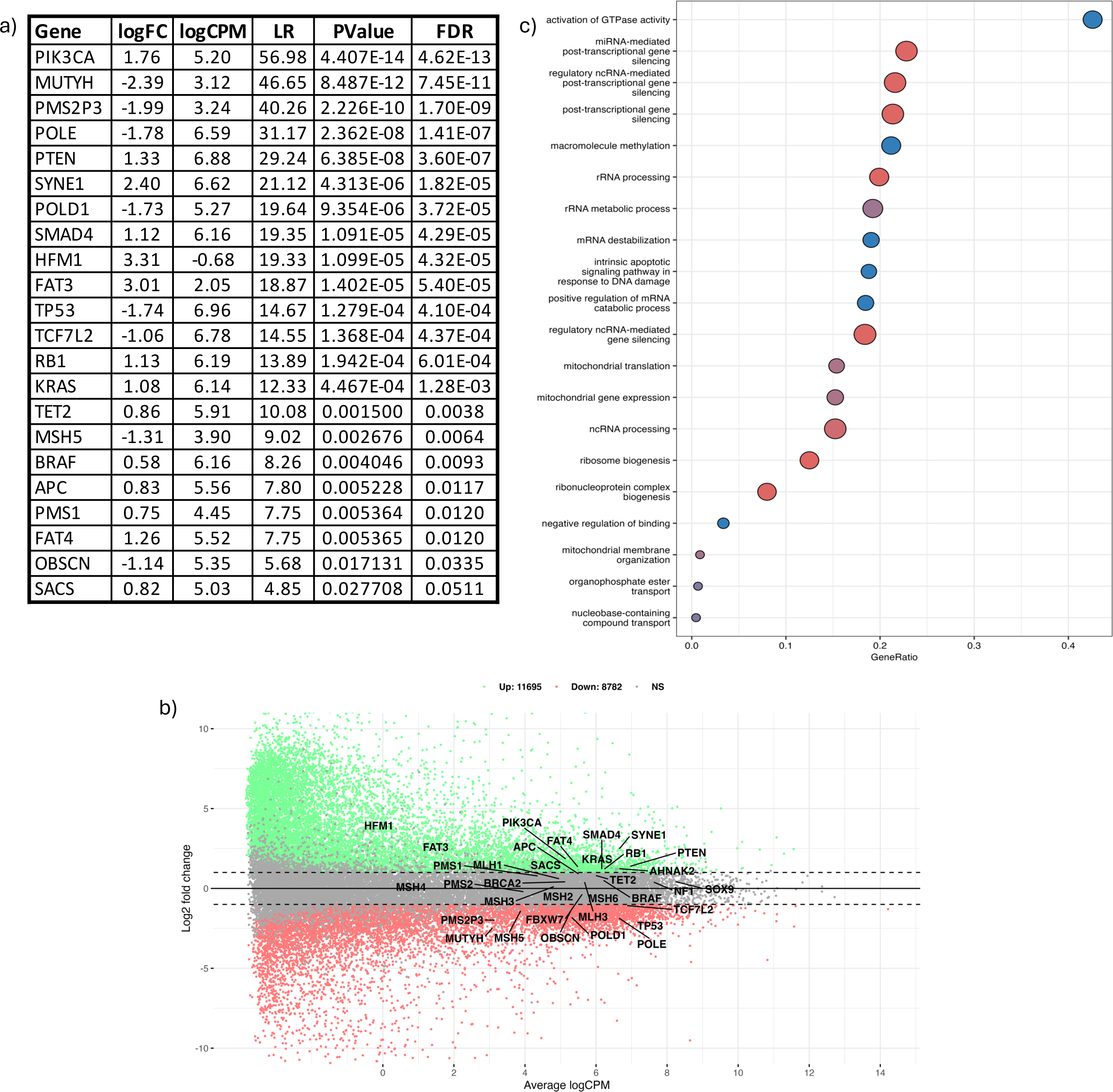
Differential gene expression (DGE) analysis among CRC tumors from 67 Hispanic/Latino patients and 67 Non-Hispanic White patients. DGE analysis identified unique gene expression patterns in our Hispanic/Latino (H/L) CRC cohort compared to Non-Hispanic White (NHW) CRC cases. The NHW cases were individually matched to our CRC cohort using demographic, clinical, and genomic data from two publicly available databases: TCGA-COAD and TCGA-READ. **a)** Differentially expressed CRC-related genes: This table shows the genes that are differentially expressed when comparing H/L and NHW CRC cases. **b)** Mean-Average (MA) plot: This scatter plot shows the upregulated and downregulated CRC-related genes according to the log2 fold change. **c)** Pathway analysis: This graph presents the distinct cellular pathways identified through pathway analysis by comparing H/L and NHW CRC cases. The graph displays the names of the pathways, adjusted p-values, and the number of genes altered in each pathway.

### Hypermutated and non-hypermutated tumors

Using the same approach (*vide supra*), we compared whether hypermutated samples differed from non-hypermutated samples (**Supplementary Fig. 9**). Differential gene expression analysis revealed 9 statistically significant CRC related genes at p-value of <0.05. These genes were MSH4, TET2, FAT4, TCF7L2, HFM1, SACS, OBSCN, SYNE1, and SMAD4 (**Supplementary Table 12**). Among the 20 top statistically significant pathways, we identified at least 6 cellular pathways implicated in CRC. These pathways were involved in dysregulations of protease activity, endopeptidase activity, calcium signaling, hydrolase activity, proteolysis activity, and catalytic activity. Notably these pathways are directly or indirectly relevant to CRC biology and may influence tumor growth, invasion, and metastasis.

### Higher proportions of 1KG-PEL-like

As above (*vide supra*), we compared tumors with > 55% 1KG-PEL-like to those with ≤ 55% (**Fig. 4c, Supplementary Fig. 10**). The differential gene expression revealed two statistically significant CRC related genes at p-value of <0.0001. These genes were TET2 and HFM1 (**Supplementary Table 13**). Among the 20 top statistically significant pathways, we identified at least 6 cellular pathways associated with CRC. These pathways were involved in dysregulated cell growth, cellular pathways, and immune responses. Notably, these pathways are either directly or indirectly relevant to CRC biology and may influence tumor growth, invasion, and metastasis.

### Gene fusions

While gene fusions are atypical in CRC, the emergence of fusion-targeted therapeutics underscores the crucial need for identifying clinically actionable fusions. [34–36] To identify gene fusions, we performed gene fusion analysis using star fusion in all the tumors (**Fig. 5 and Supplementary Table 14**). With this high genome coverage, we detected in average 2 gene fusion events per sample. Among these events, we identified 21 samples with gene fusions. Fusions typically involved one of the following three partners: either Mismatch Repair System Component Pseudogene 9 (PMS2P9), Mismatch Repair System Component Pseudogene 11 (PMS2P11), or Mismatch Repair System Component Pseudogene 9 (PMS2P9). One of these were typically fused with either Coiled-Coil Domain Containing 146 (CCDC146) or AC105052.5.[37, 38] Most of these fusions were more prevalent in samples with MSS (90%) than in MSI (10%). Most also occurred in patients with 1KG-PEL-like > 55%.

**Figure 5:**
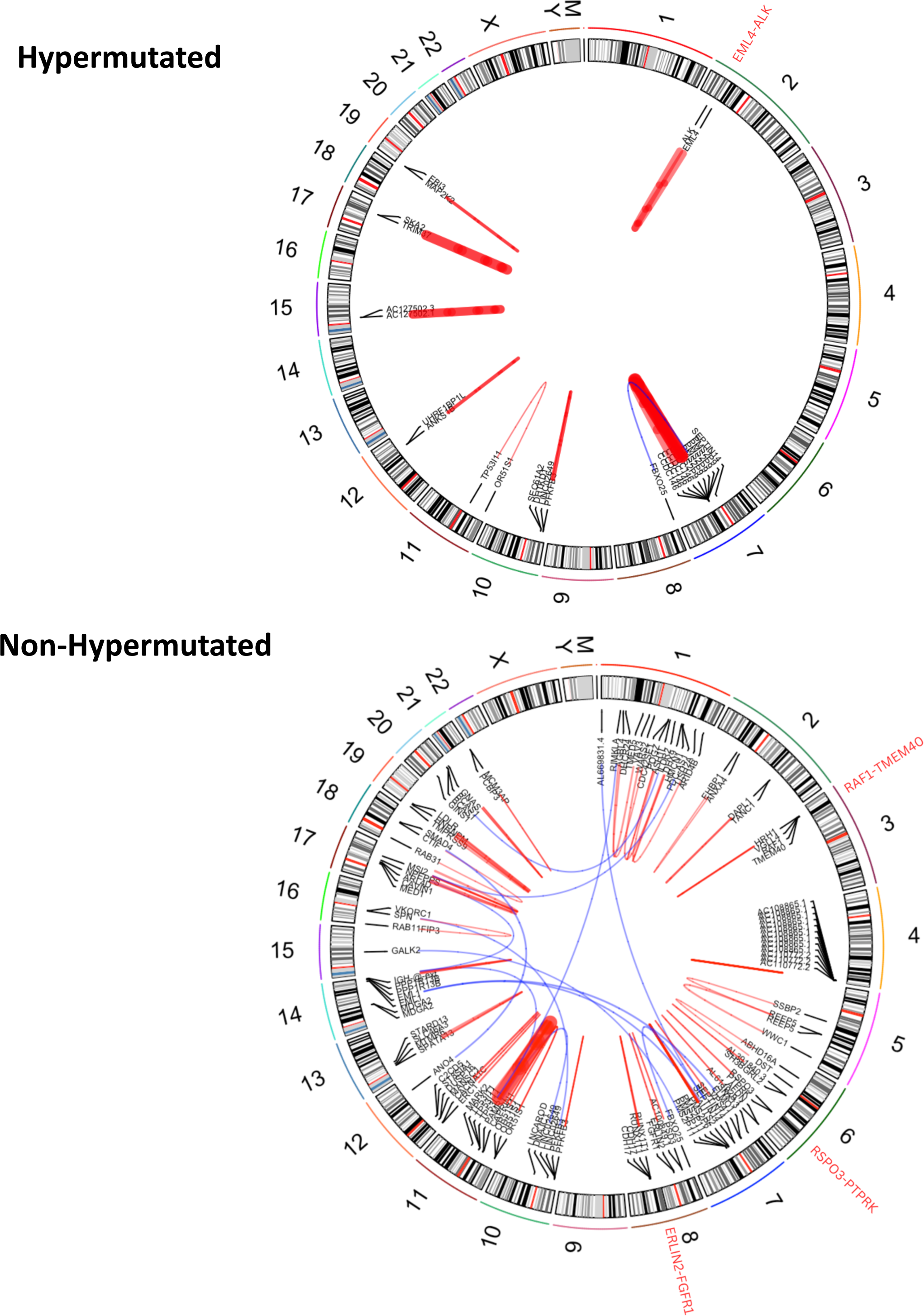
Gene Fusion Analysis stratified by Tumor Mutational Burden (TMB) in CRC Tumors from 67 Hispanic/Latino Patients. This circle plot illustrates the gene fusions detected across all samples from our Hispanic/Latino (H/L) colorectal cancer (CRC) cohort, stratified by hypermutated and non-hypermutated tumors. Clinically actionable gene fusions are highlighted in red, providing insights into potential therapeutic targets for this patient population.

TP53I11 is a gene under the regulatory control of TP53. In four samples, TP53I11 was fused with OR51S1; these samples also had MSS and had 1KG-PEL-like > 55%. Additionally, our analysis revealed two clinically actionable gene fusions in separate samples of CRC: (1) Anaplastic Lymphoma Kinase (ALK) and (2) Fibroblast Growth Factor Receptor (FGFR). The sample with the ALK fusion was MSI; the sample with the FGFR fusion was MSS. Both samples had 1KG-PEL-like >55%. Importantly, CRCs with these fusion have been successfully treated with ALK and FGFR inhibitors.[39–42] Additionally, two patients in our cohort exhibited gene fusions relevant for CRC. (1) One colon tumor sample harbored the PTPRK-RSPO3 gene fusion. This young patient (<50) exhibited MSS and an 1KG-PEL-like >55%. The PTPRK-RSPO3 gene fusion plays a role in both activating the WNT signaling pathway,[43, 44] and promoting CRC. (2) One rectum tumor sample harbored the RAF1-TMEM40 gene fusion. This case was late-onset (≥50yrs), exhibited MSS, and was 1KG-PEL-like of >55%. RAF1 gene fusions, such as RAF1-TMEM40 (seen in our samples), are part of the broader RAF kinase gene family; this includes BRAF, RAF1, and CRAF. These gene fusions are known to activate the MAPK signaling pathway and have been found across various types of cancers. RAF1 fusions can contribute to oncogenic activity in CRC. [45]

### Altered pathways

The 67 samples underwent integrated analysis for mutations, copy number, mRNA expression, and genetic similarity in 67 tumors. Our objective was to understand of how some well-defined pathways were deregulated. We grouped samples by hypermutation status and identified recurrent alterations in WNT, TGFB, TP53, IGF2/PI3K, RTK/RAS, and combined RTK/RAS/PI3K pathways (**Fig. 6**).

**Figure 6:**
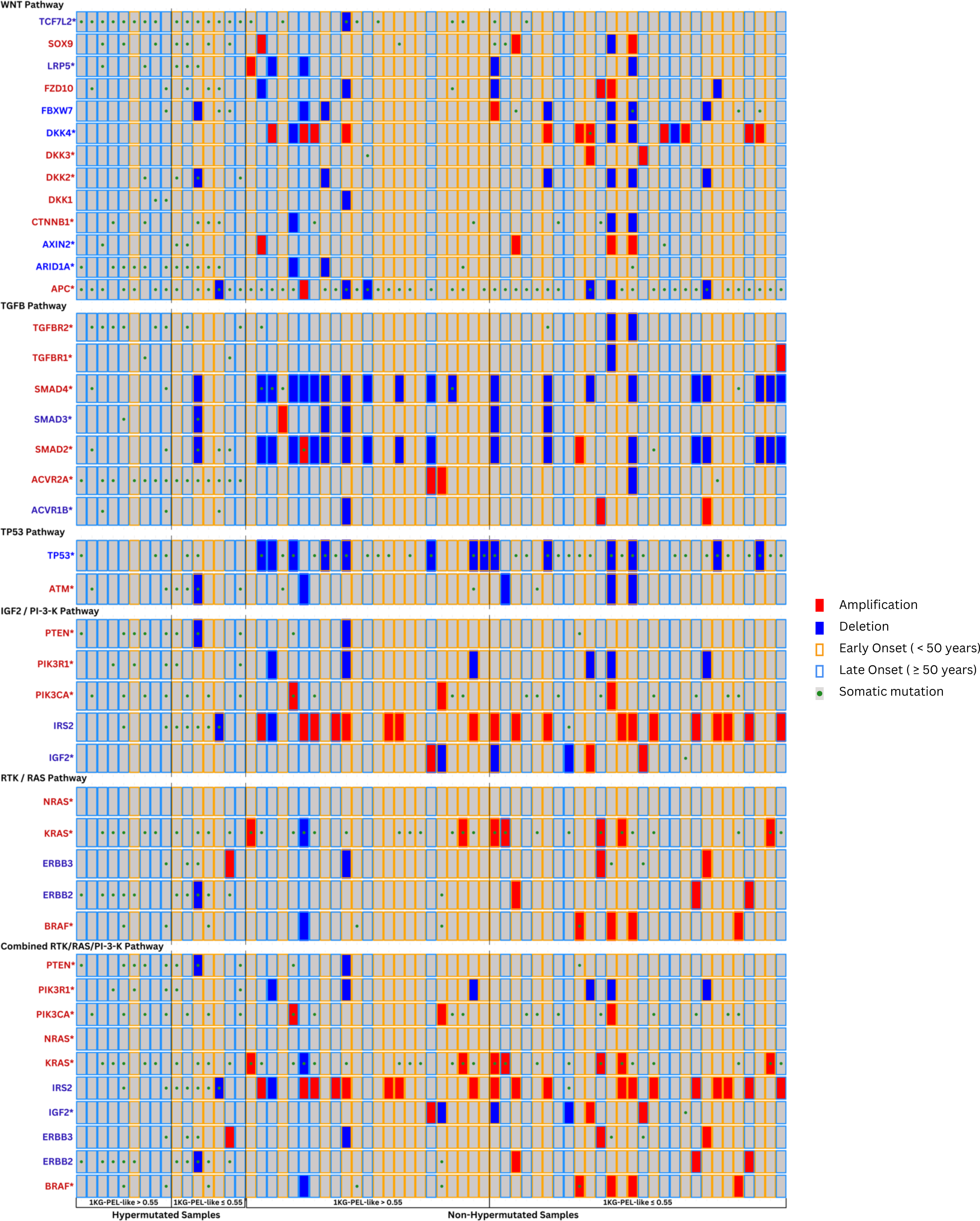
Integrative Genomic Alteration Patterns in Selected Pathways Among CRC Tumors from 67 Hispanic/Latino Patients. This grid represents integrative genomic alteration patterns in the WNT, TGFB, TP53, IGF2/PI3K, RTK/RAS, and combined RTK/RAS/PI3K pathways among colorectal cancer (CRC) tumors from 67 Hispanic/Latino patients. Each column corresponds to an individual case, and each row represents a gene. The grid is divided into hypermutated and non-hypermutated samples, further subdivided by high (>55%) or low (55%) 1KG-PEL-like similarity proportion. The color codes represent different attributes: red for amplifications, blue for deletions, a yellow border for early onset (<50 years), a blue border for late onset (>50 years), and a green dot for somatic mutations.

Ninety-six percent of tumors (64/67) had perturbations in the WNT signaling pathway; accordingly, a previous study (N=165) reported such perturbations in 93% of tumors. In our cohort, 48% of the tumors (31/64) had an 1KG-PEL-like >55%. Fifty-three percent (36/67) were early onset. As expected, we found higher mutations (80%) than SCNA (10%) in APC. CTNNB1 exhibited three times as many mutations (12%) as SCNAs (4%). Both mutations and SCNAs were found in SOX9 and TCF7L2, as well as the DKK family members and AXIN2, FBXW7 and ARID1A [46, 47]. In the RNAseq analysis results we obtained, the typically considered tumor suppressor genes ARID1A and AXIN2 were downregulated (meaning their expression levels were decreased), while the oncogene CTNNB1 was upregulated (meaning its expression level was increased) **Transcriptomics comparison with public databases: Hispanics/Latinos and Non-Hispanic Whites**). Altogether, we observed expression differences between the Hispanic and Latino samples vs. the Non-Hispanic White samples (Fig 4). In the Hispanic and Latino cohort, various genetic alterations were observed, including mutations, deletions, amplifications and dysregulations in WNT-related genes such as TCFL2, SOX9, LRP5, FZD10, FBXW7, DKK4, DKK3, DKK2, DKK1, CTNNB1, AXIN2, and ARID1A, alongside APC mutations in tumors (**Fig. 6**). Thus, this tumor suppressor gene plays a crucial role in disrupting the WNT signaling pathway.

Genetic alterations in both the PI3K and RAS pathways are common in CRC. According to our results, 86% of tumors had perturbations in the combined RTK/RAS/PI3K pathway. From these, 47% had an 1KG-PEL-like of >55% and 55% were early onset. We found higher number of mutations and SCNAs (amplifications and deletions) in IRS2 than in IGF2. PIK3CA exhibited nearly four times more mutations than PIK3R1. PIK3CA also exhibited amplifications but no deletions. Conversely, PIK3R1 had deletions but no amplifications. Compared to tumors with 1KG-PEL-like <55%, slightly more tumors with 1KG-PEL-like >55% presented KRAS mutations and double the number of the BRAF mutations. Interestingly, none of the tumors presented NRAS mutations. PTEN deletions were present in tumors with different 1KG-PEL-like proportions. We also investigated mutations in the erythroblastic leukemia viral oncogene homolog (ERBB) family of receptors due to their significant translational implications. Specifically, ERBB2 and ERBB3 mutations were identified in 16% and 7% of the tumors, respectively, with the majority exhibiting 1KG-PEL-like proportions <55%. Oncogenes PIK3CA, NRAS, KRAS and BRAF were significantly upregulated. Compared to a previous study [13], we identified slight differences in perturbations involving RAS and PI3K pathways. Altogether, common CRC pathways exhibited genomic heterogeneity for our Hispanic and Latino cohort.

The TGF-b signaling pathway is known to be deregulated in CRC and other cancers [48]. In 70% of the 67 samples, we found genomic perturbations in TGFBR1, TGFBR2, ACVR2A, ACVR1B, SMAD2, SMAD3 and SMAD4. In this 70%, almost half of the samples had 1KG-PEL-like >55% and were early onset. Consistent with a previous study [12], we found alterations in TP53 in 63% of the tumors. Additionally, we observed more than double the expected alterations in ATM, a kinase that phosphorylates and activates P53 after DNA damage; 19% of the tumors exhibited these alterations. Interestingly, the tumor mutation gene TP53 was significantly downregulated in the Hispanic and Latino population vs. the Non-Hispanic White population (Fig. 4).

## DISCUSSION

This study provides a comprehensive integrative analysis of 67 colorectal tumor and germline matching samples from Hispanic and Latino individuals. It yielded several key insights into the biology of colorectal cancer (CRC) and has highlighted potential therapeutic targets. For example, we observed interesting differences between the hypermutated (MSI) and non-hypermutated tumors (MSS). For example, the non-hypermutated samples were predominantly located in the left colon. Some of these samples exhibited high 1KG-PEL-like while others had mostly 1KG-EUR-like. In contrast, the hypermutated samples were mainly found in the right colon and exhibited predominantly 1KG-PEL-like. (**Fig. 1**). We do not know why most of the hypermutated samples came from the right colon and exhibited predominantly 1KG-PEL-like. Given that patients with MSI-related cancers, which are hypermutated, tend to have better survival rates, the mutation rate might serve as a more effective prognostic indicator. Additionally, the high prevalence of 1KG-PEL-like in these samples could also serve as another important prognostic factor.

In general, the percentages of genetic similarity populations were not associated with CRC tumor mutations. However, one exception was the SMAD4 gene; tumors with mutated SMAD4 had high (>55%) 1KG-PEL-like. Previous studies have demonstrated associations between genetic similarity differences and various cancers, including breast, melanoma, and prostate cancer.[49–56] Two prior studies examined the association of genetic similarity with CRC in Hispanic and Latino populations – in Puerto Ricans[11] and Colombians[57]. Their results suggest slight differences primarily in the 1KG-AFR-like component. In our study, SMAD4-mutated tumors were significantly more frequent—almost seven times higher—in patients with an 1KG-PEL-like proportion of >55%. However, the exact magnitude of the risk increase remains uncertain due to the wide confidence interval observed in the data. The high prevalence of SMAD4 mutations in our cohort of Hispanic and Latino individuals with CRC in Los Angeles is concerning due to its association with poor prognosis and treatment resistance. SMAD4 is a crucial tumor suppressor gene involved in the TGF-beta signaling pathway, and its loss of function is linked to more aggressive tumor behavior, lower survival rates, and resistance to standard therapies like EGFR inhibitors and anti-VEGF therapies [86–88]. These challenges necessitate personalized treatment approaches for SMAD4-mutated tumors. Moreover, our findings suggest that genetic disparities, such as the predominance of 1KG-PEL-like genetic similarity, may also contribute to CRC disparities in this population, alongside socioeconomic factors [89]. Thus, understanding the interplay between genetic and socioeconomic factors is essential for developing targeted interventions and improving outcomes for Hispanic and Latino CRC patients.

Our study compared the mutation frequencies of commonly mutated cancer genes in our CRC cohort with those from eight publicly available CRC genomic datasets: MSK-NatCommun, MSK-JNCI, DFCI-CellReports, TCGA-Nature, TCGA-PanCancerAtlas, MSK-CancerDiscovery, MSK- Gastroenterology, and MSK-JCO-PrecisOncol **(Table 2 and Fig. 4)**. Across all datasets, APC, TP53, and KRAS were the most frequently mutated genes. These three genes were also the most frequently mutated in our cohort. Interestingly, the mutation frequency of SYNE1 in our Hispanic and Latino cohort was slightly higher than in other databases. TCF7L2 and OBSCN showed comparable or slightly elevated mutation frequencies. Our data revealed that 16 out of 19 genes had similar mutation frequencies to at least one of the public databases, with TCF7L2 and BRCA2 exhibiting significantly higher mutation frequencies compared to the MSK-JNCI dataset. We used the AACR Project GENIE dataset, which is stratified by self-reported race, to compare the mutational frequencies of the Hispanic and Latino cohort to those of the Non-Hispanic White population: we observed significant differences in mutational frequencies of TCF7L2, PIK3CA, SACS, AHNAK2, MSH3, and RB1. Although our comparisons with large-scale analyses revealed notable similarities, they also uncovered distinct molecular characteristics in the Hispanic and Latino population. Altogether, diverse populations should be more comprehensively included in genetic studies to better understand CRC disparities and identify potential targets for precision medicine.

Whole-exome sequencing and integrative analysis of the genomic data provided further insights into the pathways affected in CRC when comparing Hispanic and Latino and Non-Hispanic White populations. For example, 96% of the tumors in our cohort exhibited mutations and SCNAs in the WNT signaling pathway. Compared to gene expression in the Non-Hispanic White cohort, our cohort had unique, deregulated genes in the WNT signaling pathway: two examples are the tumor suppressor genes ARID1A and AXIN2. ARID1A has been shown to suppress MYC transcription and to develop a mechanism of resistance to cetuximab.[58, 90] MYC activation is known to result from activation of the WNT signaling pathway and inactivation of the TGF-β signaling pathway. Accordingly, we found alterations in TGF-β signaling genes in 70% of the tumors, which suggested a significant role of MYC in CRC among Hispanic and Latino patients that merits further study. Additionally, when we used differential gene expression analysis to compare our cohort with the Non-Hispanic White population, we observed that the oncogene CTNNB1 was upregulated.[59,60] CTNNB1 encodes the protein β-catenin, a crucial component of the WNT signaling pathway. Another affected component of WNT signaling was TCF7L2: tumors with 1KG-PEL-like >55% had more than double TCF7L2 mutations than tumors with 1KG-PEL-like <55. Altogether, our integrative analysis underscores the pivotal role of the WNT signaling pathway in the oncogenesis of CRC in this Hispanic and Latino patients.

According to our results, colon and rectum tumors have biological similarities which, to the best of our knowledge, were previously unknown. These similarities are important because colon and rectal tumors do not have the same standard-of-care. Irrespective of their anatomical origin, non-hypermutated tumors from our cohort had similar types of copy numbers and expression profiles. More than 90% had perturbations in one or more members of the WNT signaling pathway; all these samples exhibited predominantly 1KG-PEL-like. Additionally, from a therapeutic perspective, our data from two samples provide clinically actionable gene fusions involving ALK and FGFR. Targeted therapies, including ALK and FGFR inhibitors, have been successfully implemented in the treatment of CRC following the detection of these fusions.[39, 40] Another clinical actionable approach is the WNT-signaling inhibitors and small-molecule b-catenin inhibitors.[61–63] We find that several proteins in the RTK–RAS (drug targetable fusions) and PI3K pathways, including IGF2, ERBB2, and ERBB3 could be targets for inhibition.

## CONCLUSION

The data presented in this study provide a valuable resource for understanding the molecular characteristics of CRC in the context of 1KG-PEL-like proportions and potential therapeutic targets in an underrepresented minority group. This is the first integrative report to examine the multi-omics landscape and the associations of genetic 1KG-PEL-like with CRC in Hispanic and Latino populations, compared to multiple studies in Non-Hispanic White. Our findings suggest potential differences in the disease’s biology, which could influence screening and treatment options for patients. Further studies with larger sample sizes are needed to elucidate the genomic heterogeneity and the role of 1KG-PEL-like in the development of CRC tumors in Hispanic and Latino populations.

## MATERIALS AND METHODS

### Study population

Participants in this study were recruited through the NIH NCI grant project number U2CCA252971, a Center for Optimizing Engagement of Hispanic Colorectal Cancer Patients in Cancer Genomic Characterization Studies in the University of Southern California, Los Angeles area, that started in 2021. The University of Southern California COPECC center (https://usccopecc.org) is a member of the Participant Engagement and Cancer Genome Sequencing (PE-CGS) Network (https://pe-cgs.org) part of the Cancer Moonshot Initiative^SM^ (https://www.cancer.gov/research/key-initiatives/moonshot-cancer-initiative) that is intended to accelerate cancer research.

### Data collection

All participants completed questionnaires as part of the USC COPECC center Patient Engagement Unit (https://usccopecc.org/patient-engagement-unit/) which the main goal is to develop a patient engagement framework uniquely tailored for Hispanic cancer patients.

### Clinical data

Clinical data is obtained for all cases through USC COPECC center. Data on tumor location, tumor stage (TNM stage), MSI, primary site of diagnosis, colon side, pathological group stage at diagnosis, and medications used were collected.

### DNA and RNA extraction and quality control

Tumor tissue and blood samples were collected from study participants according to standard operating procedures through the USC COPECC center Genome Characterization Unit (https://usccopecc.org/genome-characterization-unit/).

### DNA Isolation

Genomic DNA and RNA was extracted from tissue using the Covaris truXTRAC© total NA Column kit (Woburn, MA) based on the manufacturer’s recommendations. Briefly, tissue samples were placed into Covaris Adaptive Focused Acoustics (AFA) and suspended in Lysis buffer followed by Proteinase K digestion. Tissue mixtures were then equally split for separate DNA and RNA extraction. Tissue mixtures were then ultrasonically emulsed using the Covaris E220 System. DNA or RNA sample emulsions were then separately chemically de-crosslinked and pipetted onto appropriate spin columns. DNA and RNA were then collected using provided elution buffers. DNA and RNA were quality assessed and quantified using both the (Thermo Fisher Scientific, Inc. Waltham, MA) and the Genomic DNA Screen Tape Assay utilizing the Agilent 4200 TapeStation System (Santa Clara, CA). DNA and RNA samples were stored at −80°C.

### Whole exome and RNA sequencing

For Whole exome sequencing we utilized a custom expanded exome bait set (Agilent Technologies, Inc.). Briefly, components of the expanded exome included the following probe groups: original baits from SureSelect Human All Exon V6, (Agilent Technologies, Inc.) and custom baits for select genomic regions.[64] Genomic DNA in the amount of 50-200 nanograms (ng) from colorectal tumor tissue and paired uninvolved (normal) colorectal tissue from each case was sheared in 50 microliters (μl) of TE low EDTA buffer employing the Covaris E220 system (Covaris, Inc., Woburn, MA) to target fragment sizes of 150 – 200 bp. Fragmented DNA was then converted to an adapter-ligated whole genome library using the Kapa Hyper Prep Library Prep kit (Kapa Biosciences, Inc., Wilmington, MA) according to the manufacturer’s protocol. SureSelect XT Adaptor Oligo Mix was utilized in the ligation step (Agilent Technologies, Inc.). Individual tumor adapter-ligated libraries were enriched into the exome capture reaction, and for germline each adapter-ligated library was pooled before proceeding to capture using Agilent’s SureSelect Human All Exon V6 + custom probes capture library kit. Samples that had successful libraries created were then sequenced on Illumina MiSeq technology for quality control to assess the ability of the libraries to be sequenced. Subsequently, each library was pooled and sequenced on Illumina’s NovaSeq 6000 (Illumina, San Diego, CA) using 300 cycle kit. Raw FASTQs were generated using the industry standard BCL2FASTQ v1.8.4. Mean target coverage was 105X for tumor samples and 55X for uninvolved control samples.

RNA was extracted from cells from colorectal cancer tissues samples and analyzed by RNA-seq. Sequencing libraries prepared with the TruSeq Stranded Total RNA kit (Illumina Inc), from 1 μg total RNA.

### Whole exome sequencing analysis

All sequencing reads were converted to industry standard FASTQ files using the Bcl Conversion and Demultiplexing tool (Illumina, Inc). Sequencing reads were aligned to the GRCh38 reference genome using the MEM module of BWA v0.7.17[65] and SAMTOOLS v1.9 [117] to produce BAM files. After alignment, the base quality scores were recalibrated and joint indel realignment was performed on the BAM files using GATK v4.0.10.1.[66] Duplicate read pairs were marked using PICARD v2.18.22.[67] Final BAM files were then used to identify germline and somatic events. Germline SNP and INDELS were identified using GATK haplotype caller in the constitutional sample.

### Gene mutations analysis

Somatic variant callers identified single nucleotide variants (SNVs) and small insertions and deletions (indels). Somatic mutation analysis was performed by identifying overlapping variant calls generated by Strelka[68] and Mutect.[69] Due to the diffuse nature of these tumor samples, we retained somatic mutations that had a minor allele frequency of 4%, and manually reviewed variants in the KRAS region using IGV.[70] We compared these mutations against germline results to confirm and verify the somatic mutations. After filtering and manually reviewing somatic variants, VCFs were then annotated with Ensembl’s VEP[71] tool and converted to the MAF file format for visualization purposes. MAFtools[72] was utilized in the generation of oncoplots, annotated with demographic information associated with each sample.

### Somatic Copy-Number Alterations (SCNAs) analysis

For chromosomal and sub-chromosomal changes analysis we used GISTIC2.0 [25] best practices to identify genes targeted by somatic copy-number alterations (SCNAs) and SCNA profiles into underlying arm-level and focal alterations.

### Genetic Similarity analysis

We used GATK HaplotypeCaller v4.0.10.1 to generate germline VCF files for genetic similarity analysis. Known population genotype data were downloaded from the 1000 Genomes Project phase 3 and grouped by super population for genetic similarity analysis.[73] VCF subsetting and merging were performed using VCFtools v0.1.17 and SnpSift v4.3t.[74,75] VCF genotype allele coding (0/1) was converted to numeric (012) using VCFtools and PLINK v1.90b6.7.[76] All genetic similarity analyses were performed on autosomal chromosomes. The population admixture was estimated using STRUCTURE v2.3.4.[77] We subset and merged all VCF files by a list of 1766 genetic similarity-informative markers. The result was converted to Structure format by PLINK. To run STRUCTURE, we set up population numbers k=5, NUMREPS=2000, and BURNIN=50000. Principal component analysis (PCA) was performed on the same dataset by R functions[78]. All similarity analysis results were visualized using R v3.6.0 ggplot2 v3.4.1[79] package. For initial analysis, genotypes for a total of 1,766 genetic similarity-informative markers were extracted and used for estimating genetic similarity based upon five genetic similarity super continental populations including 1000 Genomes Project African-like (1KG-AFR-like), 1000 Genomes Project Peruvian-in-Lima-like (1KG-PEL-like), 1000 Genomes Project European-like (1KG-EUR-like), 1000 Genomes Project East-Asian-like (1KG-EAS-like), and 1000 Genomes Project South-Asian-like (1KG-SAS-like) using STRUCTURE (**Fig. 1**).

### RNA sequencing analysis

Alignment of raw sequence reads to the human transcriptome (hg38) was performed via Rsubread[80] and transcript abundance estimates were normalized and differentially expressed genes (DEG) identified using a standard edgeR pipeline. Functional annotation of gene sets: Pathway enrichment analysis and gene set enrichment analysis (GSEA) were performed using gene sets from the Molecular signatures database (MSigDB). In addition, R package gage (Generally Applicable Gene-set Enrichment) where used to identify enriched pathway and pathview to visualize these pathways. For transcript-aware analyses, the FASTQ files were aligned with salmon[81] and differentially enriched transcripts were identified using DRIMSeq[82] in a similar workflow to edgeR[83].

### Gene fusions analysis

For gene fusion we used STAR-Fusion that is built on top of the STAR aligner according with its best practices.[84] We ran STAR version 2.5.3a, STAR index was created, and reads were aligned to the same human reference genome than used for the WES analysis.

### Statistical analysis

In this study, we included 67 subjects recruited from USC COPECC, of which all passed the genomic sequencing quality controls (**Supplemental Table 1**). Clinical and demographic data were evaluated from each participant. The demographic variables evaluated in the study included gender, age, race and ethnicity. In addition, the following clinical data, including tumor characteristics were evaluated: tumor location, tumor stage (TNM stage), MSI, primary site of diagnosis, colon side, pathological group stage at diagnosis, mutation status of CRC related genes and medications used. Genetic similarity was modeled as a categorical variable (dichotomous for each genetic similairty population: less or equal than median genetic similarity levels).

Demographic, clinical and genomic characteristics among CRC cases were evaluated according to CRC status using Pearson’s chi-square or Fisher’s exact test, as appropriate for categorical variables. Overall, since a previous efforts [11–12] show no different of unadjusted and adjusted ORs, we present unadjusted ORs and its 95% confidence interval (CI) for the association of similarity and each tumor characteristic.

### WES Comparison with Public Databases

cBioPortal [85] for Cancer Genomics that is highly regarded resource for interactive exploration of multidimensional cancer genomics datasets was used to extract genomic data from publicly available datasets and comparisons were evaluated using Pearson’s chi-square or Fisher’s exact test, as appropriate.

### Transcriptomics Comparison with Public Databases

Transcriptomic data of 67 Non-Hispanic White was extracted from public databases projects TCGA-COAD and TCGA-READ projects. All individual demographic and clinical data from the 67 CRC tumors from Non-Hispanic White were matched to the same data from our 67 Hispanic and Latino CRC tumor samples. Differential Gene expression analyses were performed as mentioned before in the RNA sequencing analysis section.

## Data Availability

All data produced in the present study are available upon reasonable request to the authors

## Abbreviations

CRC: Colorectal Cancer
Hispanic and Latino: Hispanic/Latino
Non-Hispanic White: Non-Hispanic White
MSI: Microsatellite instability
DGE: Differential Gene Expression
SCNA: Somatic Copy Number Alteration
SNP: Single nucleotide polymorphism
US: United States.

## Acknowledgements

The authors would like to thank the Department of Integrative Translational Sciences at City of Hope (COH), the Cancer Control and Population Sciences program (P30CA033572) within the City of Hope Comprehensive Cancer Center, as well as, the NIH NCI grant project number U2CCA252971, the Genomic Characterization Unit and the Center for Optimizing Engagement of Hispanic Colorectal Cancer Patients in Cancer Genomic Characterization (COPECC) studies in the University of Southern California, Los Angeles. In addition, the authors would like to thank the patients, families, advocates, recruiters and all the personnel involved in the study for their contribution in this study.

