## Supplemental Figures and Tables for "Integrative multi-omics profiling of colorectal cancer from a Hispanic/Latino cohort of patients"

**Supplementary Index**


| | |
|---|---|
| Supplementary Figure 1 | 8 |
| Supplementary Figure 2 | 9 |
| Supplementary Figure 3 | 12 |
| Supplementary Figure 4 | 13 |
| Supplementary Figure 5 | 14 |
| Supplementary Figure 6 | 15 |
| Supplementary Figure 7 | 17 |
| Supplementary Figure 8 | 19 |
| Supplementary Figure 9 | 21 |
| Supplementary Figure 10 | 23 |
| Supplementary Table 1 | 2 |
| Supplementary Table 2 | 3 |
| Supplementary Table 3 | 4 |
| Supplementary Table 4 | 5 |
| Supplementary Table 5 | 6 |
| Supplementary Table 6 | 7 |
| Supplementary Table 7 | 10 |
| Supplementary Table 8 | 11 |
| Supplementary Table 9 | 16 |
| Supplementary Table 10 | 18 |
| Supplementary Table 11 | 20 |
| Supplementary Table 12 | 22 |
| Supplementary Table 13 | 24 |
| Supplementary Table 14 | 25 |

**Supplementary Table 1: Comprehensive Summary of Sequencing Data Quality Metrics and Statistics**

This table provides a detailed summary of the quality metrics and statistics for our sequencing data, including **a)** germline Whole Exome Sequencing (WES) data, **b)** tumor WES data, and **c)** tumor RNA sequencing data. (**Reference Supplementary Table 1 Excel File for complete table)**

**a)**

| Sample | raw_total_sequences | filtered_sequences | sequences | is_sorted | 1st_fragments | last_fragments | reads_mapped | reads_mapped_and_paired | reads_unmapped | reads_properly_paired | reads_paired |
|---|---|---|---|---|---|---|---|---|---|---|---|
| 1 | 223581859 | 26619434 | 196962425 | 1 | 98451471 | 98510954 | 196962425 | 196936317 | 0 | 195123028 | 196962425 |
| 2 | 192693725 | 18985685 | 173708040 | 1 | 86817051 | 86890989 | 173708040 | 173649124 | 0 | 172426956 | 173708040 |
| 3 | 142544847 | 11806495 | 130738352 | 1 | 65391005 | 65347347 | 130738352 | 130695313 | 0 | 129602674 | 130738352 |
| 4 | 207518783 | 21223495 | 186295288 | 1 | 93155376 | 93139912 | 186295288 | 186257389 | 0 | 185184553 | 186295288 |
| 5 | 125381279 | 10371047 | 115010232 | 1 | 57494939 | 57515293 | 115010232 | 114955901 | 0 | 114545455 | 115010232 |
| 6 | 157941897 | 15080639 | 142861258 | 1 | 71436966 | 71424292 | 142861258 | 142807076 | 0 | 141750020 | 142861258 |
| 7 | 192898477 | 23208823 | 169689654 | 1 | 84821621 | 84868033 | 169689654 | 169639756 | 0 | 168428970 | 169689654 |
| 8 | 211708999 | 21206536 | 190502463 | 1 | 95296257 | 95206206 | 190502463 | 190444708 | 0 | 189371016 | 190502463 |
| 9 | 213628241 | 36645564 | 176982677 | 1 | 88423310 | 88559367 | 176982677 | 176955302 | 0 | 175848552 | 176982677 |
| 10 | 192140118 | 31819026 | 160321092 | 1 | 80117883 | 80203209 | 160321092 | 160295043 | 0 | 159427357 | 160321092 |
| 11 | 224668342 | 42643556 | 182024786 | 1 | 91027979 | 90996807 | 182024786 | 181990365 | 0 | 180611894 | 182024786 |
| 12 | 146117872 | 12636594 | 133481278 | 1 | 66694021 | 66787257 | 133481278 | 133428852 | 0 | 132625182 | 133481278 |
| 13 | 128955329 | 10244358 | 118710971 | 1 | 59390290 | 59320681 | 118710971 | 118645566 | 0 | 117949583 | 118710971 |
| 14 | 235625023 | 20635772 | 214989251 | 1 | 107495050 | 107494201 | 214989251 | 214950393 | 0 | 213551117 | 214989251 |
| 15 | 167966036 | 15435658 | 152530378 | 1 | 76261583 | 76268795 | 152530378 | 152492105 | 0 | 151374571 | 152530378 |
| 16 | 182917123 | 18109226 | 164807897 | 1 | 82371143 | 82436754 | 164807897 | 164766797 | 0 | 164049935 | 164807897 |
| 17 | 209465924 | 24104850 | 185361074 | 1 | 92674657 | 92686417 | 185361074 | 185309559 | 0 | 184362421 | 185361074 |
| 18 | 163056795 | 14761446 | 148295349 | 1 | 74086173 | 74209176 | 148295349 | 148245377 | 0 | 147458479 | 148295349 |
| 19 | 239803978 | 26405699 | 213398279 | 1 | 106543625 | 106854654 | 213398279 | 213350284 | 0 | 212338275 | 213398279 |
| 20 | 148571449 | 12514014 | 136057435 | 1 | 68024843 | 68032592 | 136057435 | 135996546 | 0 | 135198589 | 136057435 |
| 21 | 203333707 | 17883660 | 185450047 | 1 | 92749478 | 92700569 | 185450047 | 185396529 | 0 | 184577484 | 185450047 |
| 22 | 145475944 | 11767672 | 133708272 | 1 | 66880117 | 66828155 | 133708272 | 133654310 | 0 | 133064307 | 133708272 |
| 23 | 145498276 | 14350787 | 131147489 | 1 | 65535802 | 65611687 | 131147489 | 131112752 | 0 | 130437032 | 131147489 |
| 24 | 236251933 | 27366198 | 208885735 | 1 | 104306011 | 104579724 | 208885735 | 208852371 | 0 | 207853086 | 208885735 |
| 25 | 157701548 | 14373041 | 143328507 | 1 | 71579718 | 71748789 | 143328507 | 143274545 | 0 | 142468987 | 143328507 |
| 26 | 129215468 | 11592339 | 117623129 | 1 | 58791295 | 58831834 | 117623129 | 117570609 | 0 | 116918356 | 117623129 |
| 27 | 180127252 | 18817570 | 161309682 | 1 | 80553779 | 80755903 | 161309682 | 161272423 | 0 | 160375145 | 161309682 |
| 28 | 171679972 | 16915672 | 154764300 | 1 | 77197189 | 77567111 | 154764300 | 154719155 | 0 | 153626792 | 154764300 |
| 29 | 149303066 | 14027235 | 135275831 | 1 | 67571055 | 67704776 | 135275831 | 135231883 | 0 | 134374302 | 135275831 |
| 30 | 219930935 | 23111764 | 196819171 | 1 | 98279173 | 98539998 | 196819171 | 196783372 | 0 | 195529001 | 196819171 |
| 31 | 177613122 | 15252137 | 162360985 | 1 | 81124542 | 81236443 | 162360985 | 162327257 | 0 | 161297656 | 162360985 |
| 32 | 233879816 | 24838913 | 209040903 | 1 | 104372469 | 104668434 | 209040903 | 209008198 | 0 | 207527508 | 209040903 |
| 33 | 148616035 | 13412129 | 135203906 | 1 | 67602895 | 67601011 | 135203906 | 135144679 | 0 | 134385362 | 135203906 |
| 34 | 178235686 | 19435910 | 158799776 | 1 | 79229314 | 79570462 | 158799776 | 158765763 | 0 | 157551695 | 158799776 |
| 35 | 177962172 | 16416124 | 161546048 | 1 | 80880256 | 80665792 | 161546048 | 161460043 | 0 | 160464844 | 161546048 |
| 36 | 183157086 | 21212016 | 161945070 | 1 | 80907998 | 81037072 | 161945070 | 161912020 | 0 | 160800858 | 161945070 |
| [illegible] | [illegible] | [illegible] | [illegible] | [illegible] | [illegible] | [illegible] | [illegible] | [illegible] | [illegible] | [illegible] | [illegible] |

**b)**

| |
|---|
| [illegible] |

**c)**

| |
|---|
| [illegible] |

**Supplementary Table 2:** Distribution of tumor characteristics by genetic similarity proportions relative to the 1000 Genomes Project reference populations among CRC tumors from 67 Hispanic/Latino patients.

| | 1000 Genomes Project Peruvian-in-Lima-like Similarity (n = 67) | | | 1000 Genomes Project European-like Similarity (n = 67) | | |
|---|---|---|---|---|---|---|
| | 1KG-PEL-like ≤ 0.55 n (%) | 1KG-PEL-like > 0.55 n (%) | p-value | 1KG-EUR-like ≤ 0.38 n (%) | 1KG-EUR-like > 0.38 n (%) | p-value |
| **Tumor Location** | | | | | | |
| Colon | 22 (32.84%) | 20 (29.85%) | 1 | 21 (31.34%) | 21 (31.34%) | 1 |
| Rectum | 13 (19.40%) | 12 (17.91%) | | 13 (19.40%) | 12 (17.91%) | |
| **Tumor Stage** | | | | | | |
| Early (Stage I or II) | 9 (13.43%) | 10 (14.93%) | 0.7090 | 11 (16.42%) | 8 (11.94%) | 0.9019 |
| Late (Stage III or IV) | 16 (23.88%) | 13 (19.40%) | | 14 (20.90%) | 15 (22.39%) | |
| No TNM Applicable/Unknown | 10 (14.93%) | 9 (13.43%) | | 9 (13.43%) | 10 (14.93%) | |
| **Microsatellite Status** | | | | | | |
| MSI | 6 (8.96%) | 7 (10.45%) | 0.8571 | 5 (7.46%) | 8 (11.94%) | 0.4978 |
| MSS | 29 (43.28%) | 25 (37.31%) | | 29 (43.28%) | 25 (37.31%) | |
| **Tumor Mutational Burden** | | | | | | |
| Hypermutated | 7 (10.45%) | 9 (13.43%) | 0.6225 | 7 (10.45%) | 9 (13.43%) | 0.7226 |
| Non-Hypermutated | 28 (41.79%) | 23 (34.33%) | | 27 (40.30%) | 24 (35.82%) | |
| **APC Mutation Status** | | | | | | |
| Mutated | 28 (41.79%) | 25 (37.31%) | 0.6363 | 27 (40.30%) | 26 (38.81%) | 1 |
| Wild type | 7 (10.45%) | 7 (10.45%) | | 7 (10.45%) | 7 (10.45%) | |
| **TP53 Mutation Status** | | | | | | |
| Mutated | 23 (34.33%) | 17 (25.37%) | 0.4237 | 17 (25.37%) | 23 (34.33%) | 0.1633 |
| Wild type | 12 (17.91%) | 15 (22.39%) | | 17 (25.37%) | 10 (14.93%) | |
| **KRAS Mutation Status** | | | | | | |
| Mutated | 15 (22.39%) | 17 (25.37%) | 0.5514 | 19 (28.36%) | 13 (19.40%) | 0.2686 |
| Wild type | 20 (29.85%) | 15 (22.39%) | | 15 (22.39%) | 20 (29.85%) | |
| **SYNE1 Mutation Status** | | | | | | |
| Mutated | 13 (19.40%) | 13 (19.40%) | 0.7414 | 10 (14.93%) | 16 (23.88%) | 0.1767 |
| Wild type | 22 (32.84%) | 19 (28.36%) | | 24 (35.82%) | 17 (25.37%) | |
| **TCF7L2 Mutation Status** | | | | | | |
| Mutated | 9 (13.43%) | 15 (22.39%) | 0.1213 | 13 (19.40%) | 11 (16.42%) | 0.8701 |
| Wild type | 26 (38.81%) | 17 (25.37%) | | 21 (31.34%) | 22 (32.84%) | |
| **OBSCN Mutation Status** | | | | | | |
| Mutated | 11 (16.42%) | 8 (11.94%) | 0.7552 | 7 (10.45%) | 12 (17.91%) | 0.2456 |
| Wild type | 24 (35.82%) | 24 (35.82%) | | 27 (40.30%) | 21 (31.34%) | |
| **PIK3CA Mutation Status** | | | | | | |
| Mutated | 11 (16.42%) | 8 (11.94%) | 0.7552 | 9 (13.43%) | 10 (14.93%) | 0.9387 |
| Wild type | 24 (35.82%) | 24 (35.82%) | | 25 (37.31%) | 23 (34.33%) | |
| **SACS Mutation Status** | | | | | | |
| Mutated | 10 (14.93%) | 7 (10.45%) | 0.7277 | 6 (8.96%) | 11 (16.42%) | 0.2323 |
| Wild type | 25 (37.31%) | 25 (37.31%) | | 28 (41.79%) | 22 (32.84%) | |
| **AHNAK2 Mutation Status** | | | | | | |
| Mutated | 10 (14.93%) | 8 (11.94%) | 0.9573 | 7 (10.45%) | 11 (16.42%) | 0.3676 |
| Wild type | 25 (37.31%) | 24 (35.82%) | | 27 (40.30%) | 22 (32.84%) | |
| **FAT3 Mutation Status** | | | | | | |
| Mutated | 10 (14.93%) | 7 (10.45%) | 0.7277 | 8 (11.94%) | 9 (13.43%) | 0.9432 |
| Wild type | 25 (37.31%) | 25 (37.31%) | | 26 (38.81%) | 24 (35.82%) | |
| **FAT4 Mutation Status** | | | | | | |
| Mutated | 8 (11.94%) | 8 (11.94%) | 1 | 8 (11.94%) | 8 (11.94%) | 1 |
| Wild type | 27 (40.30%) | 24 (35.82%) | | 26 (38.81%) | 25 (37.31%) | |
| **BRCA2 Mutation Status** | | | | | | |
| Mutated | 5 (7.46%) | 7 (10.45%) | 0.6239 | 6 (8.96%) | 6 (8.96%) | 1 |
| Wild type | 30 (44.78%) | 25 (37.31%) | | 28 (41.79%) | 27 (40.30%) | |
| **MSH3 Mutation Status** | | | | | | |
| Mutated | 5 (7.46%) | 5 (7.46%) | 1 | 4 (5.97%) | 6 (8.96%) | 0.5118 |
| Wild type | 30 (44.78%) | 27 (40.30%) | | 30 (44.78%) | 27 (40.30%) | |
| **SOX9 Mutation Status** | | | | | | |
| Mutated | 6 (8.96%) | 4 (5.97%) | 0.7364 | 4 (5.97%) | 6 (8.96%) | 0.5118 |
| Wild type | 29 (43.28%) | 28 (41.79%) | | 30 (44.78%) | 27 (40.30%) | |
| **PTEN Mutation Status** | | | | | | |
| Mutated | 4 (5.97%) | 6 (8.96%) | 0.501 | 4 (5.97%) | 6 (8.96%) | 0.5118 |
| Wild type | 31 (46.27%) | 26 (38.81%) | | 30 (44.78%) | 27 (40.30%) | |
| **POLE Mutation Status** | | | | | | |
| Mutated | 5 (7.46%) | 3 (4.48%) | 0.7113 | 3 (4.48%) | 5 (7.46%) | 0.4673 |
| Wild type | 30 (44.78%) | 29 (43.28%) | | 31 (46.27%) | 28 (41.79%) | |
| **FBXW7 Mutation Status** | | | | | | |
| Mutated | 6 (8.96%) | 1 (1.49%) | 0.1078 | 2 (2.99%) | 5 (7.46%) | 0.2585 |
| Wild type | 29 (43.28%) | 31 (46.27%) | | 32 (47.76%) | 28 (41.79%) | |
| **NF1 Mutation Status** | | | | | | |
| Mutated | 4 (5.97%) | 3 (4.48%) | 1 | 3 (4.48%) | 4 (5.97%) | 0.709 |
| Wild type | 31 (46.27%) | 29 (43.28%) | | 31 (46.27%) | 29 (43.28%) | |
| **MLH1 Mutation Status** | | | | | | |
| Mutated | 3 (4.48%) | 4 (5.97%) | 0.7014 | 3 (4.48%) | 4 (5.97%) | 0.709 |
| Wild type | 32 (47.76%) | 28 (41.79%) | | 31 (46.27%) | 29 (43.28%) | |
| **MSH6 Mutation Status** | | | | | | |
| Mutated | 2 (2.99%) | 5 (7.46%) | 0.2456 | 5 (7.46%) | 2 (2.99%) | 0.4275 |
| Wild type | 33 (49.25%) | 27 (40.30%) | | 29 (43.28%) | 31 (46.27%) | |
| **SMAD4 Mutation Status** | | | | | | |
| Mutated | 1 (1.49%) | 6 (8.96%) | **0.04807** | 7 (10.45%) | 0 (0.00%) | **0.0111** |
| Wild type | 34 (50.75%) | 26 (38.81%) | | 27 (40.30%) | 33 (49.25%) | |
| **BRAF Mutation Status** | | | | | | |
| Mutated | 2 (2.99%) | 4 (5.97%) | 0.4145 | 4 (5.97%) | 2 (2.99%) | 0.6728 |
| Wild type | 33 (49.25%) | 28 (41.79%) | | 30 (44.78%) | 31 (46.27%) | |
| **RB1 Mutation Status** | | | | | | |
| Mutated | 3 (4.48%) | 2 (2.99%) | 1 | 1 (1.49%) | 4 (5.97%) | 0.1974 |
| Wild type | 32 (47.76%) | 30 (44.78%) | | 33 (49.25%) | 29 (43.28%) | |
| **MSH2 Mutation Status** | | | | | | |
| Mutated | 1 (1.49%) | 4 (5.97%) | 0.1848 | 3 (4.48%) | 2 (2.99%) | 1 |
| Wild type | 34 (50.75%) | 28 (41.79%) | | 31 (46.27%) | 31 (46.27%) | |
| **TET2 Mutation Status** | | | | | | |
| Mutated | 2 (2.99%) | 3 (4.48%) | 0.6639 | 2 (2.99%) | 3 (4.48%) | 0.6728 |
| Wild type | 33 (49.25%) | 29 (43.28%) | | 32 (47.76%) | 30 (44.78%) | |
| **POLD1 Mutation Status** | | | | | | |
| Mutated | 2 (2.99%) | 2 (2.99%) | 1 | 1 (1.49%) | 3 (4.48%) | 0.3559 |
| Wild type | 33 (49.25%) | 30 (44.78%) | | 33 (49.25%) | 30 (44.78%) | |
| **MUTYH Mutation Status** | | | | | | |
| Mutated | 1 (1.49%) | 1 (1.49%) | 1 | 1 (1.49%) | 1 (1.49%) | 1 |
| Wild type | 34 (50.75%) | 31 (46.27%) | | 33 (49.25%) | 32 (47.76%) | |

**Supplementary Table 3:** Association of tumors characteristics with genetic similarity proportions relative to the 1000 Genomes Project reference populations among CRC tumors from 67 Hispanic/Latino patients.

| | Odds Ratio (95% CI) |
|---|---|
| **Tumor Location** | |
| 1KG-PEL-like ≤ 0.55 | 1 |
| 1KG-PEL-like > 0.55 | 1.02 (0.37-2.78) |
| **Tumor Stage** | |
| 1KG-PEL-like ≤ 0.55 | 1 |
| 1KG-PEL-like > 0.55 | 0.74 (0.22-2.40) |
| **Microsatellite Status** | |
| 1KG-PEL-like ≤ 0.55 | 1 |
| 1KG-PEL-like > 0.55 | 9.06 (3.68-25.15) |
| **Tumor Mutational Burden** | |
| 1KG-PEL-like ≤ 0.55 | 1 |
| 1KG-PEL-like > 0.55 | 1.55 (0.49-5.05) |
| **APC Mutation Status** | |
| 1KG-PEL-like ≤ 0.55 | 1 |
| 1KG-PEL-like > 0.55 | 0.72 (0.28-1.84) |
| **TP53 Mutation Status** | |
| 1KG-PEL-like ≤ 0.55 | 1 |
| 1KG-PEL-like > 0.55 | 0.60 (0.22-1.61) |
| **KRAS Mutation Status** | |
| 1KG-PEL-like ≤ 0.55 | 1 |
| 1KG-PEL-like > 0.55 | 1.50 (0.57-4.02) |
| **SYNE1 Mutation Status** | |
| 1KG-PEL-like ≤ 0.55 | 1 |
| 1KG-PEL-like > 0.55 | 0.76 (0.29-1.99) |
| **TCF7L2 Mutation Status** | |
| 1KG-PEL-like ≤ 0.55 | 1 |
| 1KG-PEL-like > 0.55 | 2.50 (0.90-0.07) |
| **OBSCN Mutation Status** | |
| 1KG-PEL-like ≤ 0.55 | 1 |
| 1KG-PEL-like > 0.55 | 0.73 (0.24-2.16) |
| **PIK3CA Mutation Status** | |
| 1KG-PEL-like ≤ 0.55 | 1 |
| 1KG-PEL-like > 0.55 | 0.73 (0.24-2.16) |
| **SACS Mutation Status** | |
| 1KG-PEL-like ≤ 0.55 | 1 |
| 1KG-PEL-like > 0.55 | 0.71 (0.22-2.17) |
| **AHNAK2 Mutation Status** | |
| 1KG-PEL-like ≤ 0.55 | 1 |
| 1KG-PEL-like > 0.55 | 0.84 (0.27-2.52) |
| **FAT3 Mutation Status** | |
| 1KG-PEL-like ≤ 0.55 | 1 |
| 1KG-PEL-like > 0.55 | 0.71 (0.22-2.17) |
| **FAT4 Mutation Status** | |
| 1KG-PEL-like ≤ 0.55 | 1 |
| 1KG-PEL-like > 0.55 | 1.12 (0.35-3.57) |
| **BRCA2 Mutation Status** | |
| 1KG-PEL-like ≤ 0.55 | 1 |
| 1KG-PEL-like > 0.55 | 1.65 (0.46-6.40) |
| **MSH3 Mutation Status** | |
| 1KG-PEL-like ≤ 0.55 | 1 |
| 1KG-PEL-like > 0.55 | 1.11 (0.27-4.55) |
| **SOX9 Mutation Status** | |
| 1KG-PEL-like ≤ 0.55 | 1 |
| 1KG-PEL-like > 0.55 | 0.70 (0.16-2.81) |
| **PTEN Mutation Status** | |
| 1KG-PEL-like ≤ 0.55 | 1 |
| 1KG-PEL-like > 0.55 | 1.75 (0.44-7.82) |
| **POLE Mutation Status** | |
| 1KG-PEL-like ≤ 0.55 | 1 |
| 1KG-PEL-like > 0.55 | 0.64 (0.11-2.95) |
| **FBXW7 Mutation Status** | |
| 1KG-PEL-like ≤ 0.55 | 1 |
| 1KG-PEL-like > 0.55 | 0.18 (0.01-1.17) |
| **NF1 Mutation Status** | |
| 1KG-PEL-like ≤ 0.55 | 1 |
| 1KG-PEL-like > 0.55 | 0.81 (0.14-4.21) |
| **MLH1 Mutation Status** | |
| 1KG-PEL-like ≤ 0.55 | 1 |
| 1KG-PEL-like > 0.55 | 11.43 (3.56-53.54) |
| **MSH6 Mutation Status** | |
| 1KG-PEL-like ≤ 0.55 | 1 |
| 1KG-PEL-like > 0.55 | 2.89 (0.55-23.88) |
| **SMAD4 Mutation Status** | |
| 1KG-PEL-like ≤ 0.55 | 1 |
| 1KG-PEL-like > 0.55 | 6.89 (1.04-186.52) |
| **BRAF Mutation Status** | |
| 1KG-PEL-like ≤ 0.55 | 1 |
| 1KG-PEL-like > 0.55 | 2.25 (0.38-19.32) |
| **RB1 Mutation Status** | |
| 1KG-PEL-like ≤ 0.55 | 1 |
| 1KG-PEL-like > 0.55 | 0.73 (0.08-5.11) |
| **MSH2 Mutation Status** | |
| 1KG-PEL-like ≤ 0.55 | 1 |
| 1KG-PEL-like > 0.55 | 4.34 (0.56-123.39) |
| **TET2 Mutation Status** | |
| 1KG-PEL-like ≤ 0.55 | 1 |
| 1KG-PEL-like > 0.55 | 1.66 (0.24-15.08) |
| **POLD1 Mutation Status** | |
| 1KG-PEL-like ≤ 0.55 | 1 |
| 1KG-PEL-like > 0.55 | 0.10 (0.11-11.10) |
| **MUTYH Mutation Status** | |
| 1KG-PEL-like ≤ 0.55 | 1 |
| 1KG-PEL-like > 0.55 | 1.10 (0.03-44.00) |

| | Odds Ratio (95% CI) |
|---|---|
| **Tumor Location** | |
| 1KG-EUR-like ≤ 0.38 | 1 |
| 1KG-EUR-like > 0.38 | 1.08 (0.40-2.97) |
| **Tumor Stage** | |
| 1KG-EUR-like ≤ 0.38 | 1 |
| 1KG-EUR-like > 0.38 | 1.46 (0.45-4.88) |
| **Microsatellite Status** | |
| 1KG-EUR-like ≤ 0.38 | 1 |
| 1KG-EUR-like > 0.38 | 1.82 (0.53-6.91) |
| **Tumor Mutational Burden** | |
| 1KG-EUR-like ≤ 0.38 | 1 |
| 1KG-EUR-like > 0.38 | 1.43 (0.46-4.67) |
| **APC Mutation Status** | |
| 1KG-EUR-like ≤ 0.38 | 1 |
| 1KG-EUR-like > 0.38 | 0.96 (0.29-3.26) |
| **TP53 Mutation Status** | |
| 1KG-EUR-like ≤ 0.38 | 1 |
| 1KG-EUR-like > 0.38 | 2.26 (0.83-6.40) |
| **KRAS Mutation Status** | |
| 1KG-EUR-like ≤ 0.38 | 1 |
| 1KG-EUR-like > 0.38 | 0.52 (0.19-1.38) |
| **SYNE1 Mutation Status** | |
| 1KG-EUR-like ≤ 0.38 | 1 |
| 1KG-EUR-like > 0.38 | 2.22 (0.81-6.30) |
| **TCF7L2 Mutation Status** | |
| 1KG-EUR-like ≤ 0.38 | 1 |
| 1KG-EUR-like > 0.38 | 0.81 (0.29-2.23) |
| **OBSCN Mutation Status** | |
| 1KG-EUR-like ≤ 0.38 | 1 |
| 1KG-EUR-like > 0.38 | 2.16 (0.73-6.85) |
| **PIK3CA Mutation Status** | |
| 1KG-EUR-like ≤ 0.38 | 1 |
| 1KG-EUR-like > 0.38 | 1.20 (0.41-3.60) |
| **SACS Mutation Status** | |
| 1KG-EUR-like ≤ 0.38 | 1 |
| 1KG-EUR-like > 0.38 | 2.28 (0.74-7.72) |
| **AHNAK2 Mutation Status** | |
| 1KG-EUR-like ≤ 0.38 | 1 |
| 1KG-EUR-like > 0.38 | 1.90 (0.63-6.06) |
| **FAT3 Mutation Status** | |
| 1KG-EUR-like ≤ 0.38 | 1 |
| 1KG-EUR-like > 0.38 | 1.21 (0.39-3.79) |
| **FAT4 Mutation Status** | |
| 1KG-EUR-like ≤ 0.38 | 1 |
| 1KG-EUR-like > 0.38 | 1.04 (0.33-3.30) |
| **BRCA2 Mutation Status** | |
| 1KG-EUR-like ≤ 0.38 | 1 |
| 1KG-EUR-like > 0.38 | 1.04 (0.28-3.80) |
| **MSH3 Mutation Status** | |
| 1KG-EUR-like ≤ 0.38 | 1 |
| 1KG-EUR-like > 0.38 | 1.64 (0.41-7.30) |
| **SOX9 Mutation Status** | |
| 1KG-EUR-like ≤ 0.38 | 1 |
| 1KG-EUR-like > 0.38 | 1.64 (0.41-7.30) |
| **PTEN Mutation Status** | |
| 1KG-EUR-like ≤ 0.38 | 1 |
| 1KG-EUR-like > 0.38 | 1.64 (0.41-7.30) |
| **POLE Mutation Status** | |
| 1KG-EUR-like ≤ 0.38 | 1 |
| 1KG-EUR-like > 0.38 | 1.80 (0.39-10.04) |
| **FBXW7 Mutation Status** | |
| 1KG-EUR-like ≤ 0.38 | 1 |
| 1KG-EUR-like > 0.38 | 2.71 (0.51-22.35) |
| **NF1 Mutation Status** | |
| 1KG-EUR-like ≤ 0.38 | 1 |
| 1KG-EUR-like > 0.38 | 1.40 (0.27-8.18) |
| **MLH1 Mutation Status** | |
| 1KG-EUR-like ≤ 0.38 | 1 |
| 1KG-EUR-like > 0.38 | 1.40 (0.27-8.18) |
| **MSH6 Mutation Status** | |
| 1KG-EUR-like ≤ 0.38 | 1 |
| 1KG-EUR-like > 0.38 | 0.39 (0.05-2.08) |
| **SMAD4 Mutation Status** | |
| 1KG-EUR-like ≤ 0.38 | NA |
| 1KG-EUR-like > 0.38 | NA |
| **BRAF Mutation Status** | |
| 1KG-EUR-like ≤ 0.38 | 1 |
| 1KG-EUR-like > 0.38 | 0.51 (0.06-2.96) |
| **RB1 Mutation Status** | |
| 1KG-EUR-like ≤ 0.38 | 1 |
| 1KG-EUR-like > 0.38 | 4.07 (0.53-115.74) |
| **MSH2 Mutation Status** | |
| 1KG-EUR-like ≤ 0.38 | 1 |
| 1KG-EUR-like > 0.38 | 0.69 (0.08-4.80) |
| **TET2 Mutation Status** | |
| 1KG-EUR-like ≤ 0.38 | 1 |
| 1KG-EUR-like > 0.38 | 1.56 (0.22-14.15) |
| **POLD1 Mutation Status** | |
| 1KG-EUR-like ≤ 0.38 | 1 |
| 1KG-EUR-like > 0.38 | 3.00 (0.33-89.32) |
| **MUTYH Mutation Status** | |
| 1KG-EUR-like ≤ 0.38 | 1 |
| 1KG-EUR-like > 0.38 | 1.03 (0.03-41.38) |

**Supplementary Table 4: CRC-Related Genes with mutations from Hypermutated and Non-Hypermutated Tumors in 67 Hispanic/Latino Patients.** This table details the CRC-related genes with mutations, categorized by hypermutated and non-hypermutated tumors, in a cohort of 67 Hispanic/Latino patients.

| | | Genes | | | | | | | | | | | | | | | | | | | | | | | | | | |
|---|---|---|---|---|---|---|---|---|---|---|---|---|---|---|---|---|---|---|---|---|---|---|---|---|---|---|---|---|
| | | APC | TP53 | KRAS | SYNE1 | TCF7L2 | OBSCN | PIK3CA | SACS | AHNAK2 | FAT3 | FAT4 | BRCA2 | MSH3 | SOX9 | PTEN | POLE | FBXW7 | NF1 | MLH1 | MSH6 | SMAD4 | BRAF | RB1 | MSH2 | TET2 | POLD1 | MUTYH |
| Hypermutated Samples | 1 | | | | | | | | | | | | | | | | | | | | | | | | | | | |
| | 2 | | | | | | | | | | | | | | | | | | | | | | | | | | | |
| | 3 | | | | | | | | | | | | | | | | | | | | | | | | | | | |
| | 4 | | | | | | | | | | | | | | | | | | | | | | | | | | | |
| | 5 | | | | | | | | | | | | | | | | | | | | | | | | | | | |
| | 6 | | | | | | | | | | | | | | | | | | | | | | | | | | | |
| | 7 | | | | | | | | | | | | | | | | | | | | | | | | | | | |
| | 8 | | | | | | | | | | | | | | | | | | | | | | | | | | | |
| | 9 | | | | | | | | | | | | | | | | | | | | | | | | | | | |
| | 10 | | | | | | | | | | | | | | | | | | | | | | | | | | | |
| | 11 | | | | | | | | | | | | | | | | | | | | | | | | | | | |
| | 12 | | | | | | | | | | | | | | | | | | | | | | | | | | | |
| | 13 | | | | | | | | | | | | | | | | | | | | | | | | | | | |
| | 14 | | | | | | | | | | | | | | | | | | | | | | | | | | | |
| | 15 | | | | | | | | | | | | | | | | | | | | | | | | | | | |
| | 16 | | | | | | | | | | | | | | | | | | | | | | | | | | | |
| | SUM | 13 | 5 | 9 | 12 | 15 | 12 | 7 | 12 | 12 | 12 | 10 | 10 | 9 | 7 | 8 | 7 | 3 | 6 | 7 | 7 | 2 | 3 | 5 | 5 | 5 | 4 | 1 |
| Non-hypermutated Samples | 1 | | | | | | | | | | | | | | | | | | | | | | | | | | | |
| | 2 | | | | | | | | | | | | | | | | | | | | | | | | | | | |
| | 3 | | | | | | | | | | | | | | | | | | | | | | | | | | | |
| | 4 | | | | | | | | | | | | | | | | | | | | | | | | | | | |
| | 5 | | | | | | | | | | | | | | | | | | | | | | | | | | | |
| | 6 | | | | | | | | | | | | | | | | | | | | | | | | | | | |
| | 7 | | | | | | | | | | | | | | | | | | | | | | | | | | | |
| | 8 | | | | | | | | | | | | | | | | | | | | | | | | | | | |
| | 9 | | | | | | | | | | | | | | | | | | | | | | | | | | | |
| | 10 | | | | | | | | | | | | | | | | | | | | | | | | | | | |
| | 11 | | | | | | | | | | | | | | | | | | | | | | | | | | | |
| | 12 | | | | | | | | | | | | | | | | | | | | | | | | | | | |
| | 13 | | | | | | | | | | | | | | | | | | | | | | | | | | | |
| | 14 | | | | | | | | | | | | | | | | | | | | | | | | | | | |
| | 15 | | | | | | | | | | | | | | | | | | | | | | | | | | | |
| | 16 | | | | | | | | | | | | | | | | | | | | | | | | | | | |
| | 17 | | | | | | | | | | | | | | | | | | | | | | | | | | | |
| | 18 | | | | | | | | | | | | | | | | | | | | | | | | | | | |
| | 19 | | | | | | | | | | | | | | | | | | | | | | | | | | | |
| | 20 | | | | | | | | | | | | | | | | | | | | | | | | | | | |
| | 21 | | | | | | | | | | | | | | | | | | | | | | | | | | | |
| | 22 | | | | | | | | | | | | | | | | | | | | | | | | | | | |
| | 23 | | | | | | | | | | | | | | | | | | | | | | | | | | | |
| | 24 | | | | | | | | | | | | | | | | | | | | | | | | | | | |
| | 25 | | | | | | | | | | | | | | | | | | | | | | | | | | | |
| | 26 | | | | | | | | | | | | | | | | | | | | | | | | | | | |
| | 27 | | | | | | | | | | | | | | | | | | | | | | | | | | | |
| | 28 | | | | | | | | | | | | | | | | | | | | | | | | | | | |
| | 29 | | | | | | | | | | | | | | | | | | | | | | | | | | | |
| | 30 | | | | | | | | | | | | | | | | | | | | | | | | | | | |
| | 31 | | | | | | | | | | | | | | | | | | | | | | | | | | | |
| | 32 | | | | | | | | | | | | | | | | | | | | | | | | | | | |
| | 33 | | | | | | | | | | | | | | | | | | | | | | | | | | | |
| | 34 | | | | | | | | | | | | | | | | | | | | | | | | | | | |
| | 35 | | | | | | | | | | | | | | | | | | | | | | | | | | | |
| | 36 | | | | | | | | | | | | | | | | | | | | | | | | | | | |
| | 37 | | | | | | | | | | | | | | | | | | | | | | | | | | | |
| | 38 | | | | | | | | | | | | | | | | | | | | | | | | | | | |
| | 39 | | | | | | | | | | | | | | | | | | | | | | | | | | | |
| | 40 | | | | | | | | | | | | | | | | | | | | | | | | | | | |
| | 41 | | | | | | | | | | | | | | | | | | | | | | | | | | | |
| | 42 | | | | | | | | | | | | | | | | | | | | | | | | | | | |
| | 43 | | | | | | | | | | | | | | | | | | | | | | | | | | | |
| | 44 | | | | | | | | | | | | | | | | | | | | | | | | | | | |
| | 45 | | | | | | | | | | | | | | | | | | | | | | | | | | | |
| | 46 | | | | | | | | | | | | | | | | | | | | | | | | | | | |
| | 47 | | | | | | | | | | | | | | | | | | | | | | | | | | | |
| | 48 | | | | | | | | | | | | | | | | | | | | | | | | | | | |
| | 49 | | | | | | | | | | | | | | | | | | | | | | | | | | | |
| | 50 | | | | | | | | | | | | | | | | | | | | | | | | | | | |
| | 51 | | | | | | | | | | | | | | | | | | | | | | | | | | | |
| | SUM | 40 | 35 | 23 | 14 | 9 | 7 | 12 | 5 | 6 | 5 | 6 | 2 | 1 | 3 | 2 | 1 | 4 | 1 | 0 | 0 | 5 | 3 | 0 | 0 | 0 | 0 | 1 |

**Supplementary Table 5: Number of Tumor Samples with CRC-Related Mutations, Average 1KG-PEL-like Proportion, and Age at Diagnosis in 67 Hispanic/Latino CRC Patients.** This table presents the number of tumor samples observed with CRC-related mutations, alongside the average proportion of 1000 Genomes Project Peruvian-in-Lima-like (1KG-PEL-like) and the age at diagnosis.

| Mutation Type | Number of Samples | Average 1KG-PEL-like | Average Age at Dx |
|---|---|---|---|
| **BRAF V600E** | 1 | 0.457 | 60.1 |
| **KRAS G12A** | 2 | 0.646 | 44.55 |
| **KRAS G12C** | 1 | 0.548 | 52.6 |
| **KRAS G12D** | 6 | 0.4875 | 50.25 |
| **KRAS G12V** | 11 | 0.6016 | 57.95 |
| **KRAS G13D** | 7 | 0.6569 | 49.39 |

**Supplementary Table 6: Number of tumor samples with mutations in tumor suppressor genes and associated demographic data in 67 Hispanic/Latino CRC patients.** This table presents the number of tumor samples with mutations in key tumor suppressor genes (APC, TP53, BRCA2, PTEN, FBXW7, NF1, and SMAD4) in a cohort of 67 Hispanic/Latino colorectal cancer (CRC) patients. Alongside the mutation data, the table includes the average proportion of 1000 Genomes Project Peruvian-in-Lima-like (1KG-PEL-like) and the age at diagnosis.

| Gene | Number of Samples | Average 1KG-PEL-like | Average Age at Dx |
|---|---|---|---|
| **APC** | 53 | 0.580 | 52.8 |
| **TP53** | 40 | 0.555 | 51.3 |
| **BRCA2** | 12 | 0.624 | 50.2 |
| **PTEN** | 10 | 0.620 | 46.5 |
| **FBXW7** | 7 | 0.570 | 58.0 |
| **NF1** | 7 | 0.633 | 42.6 |
| **SMAD4** | 7 | 0.653 | 48.0 |

### Non-Hypermutated Colon Samples

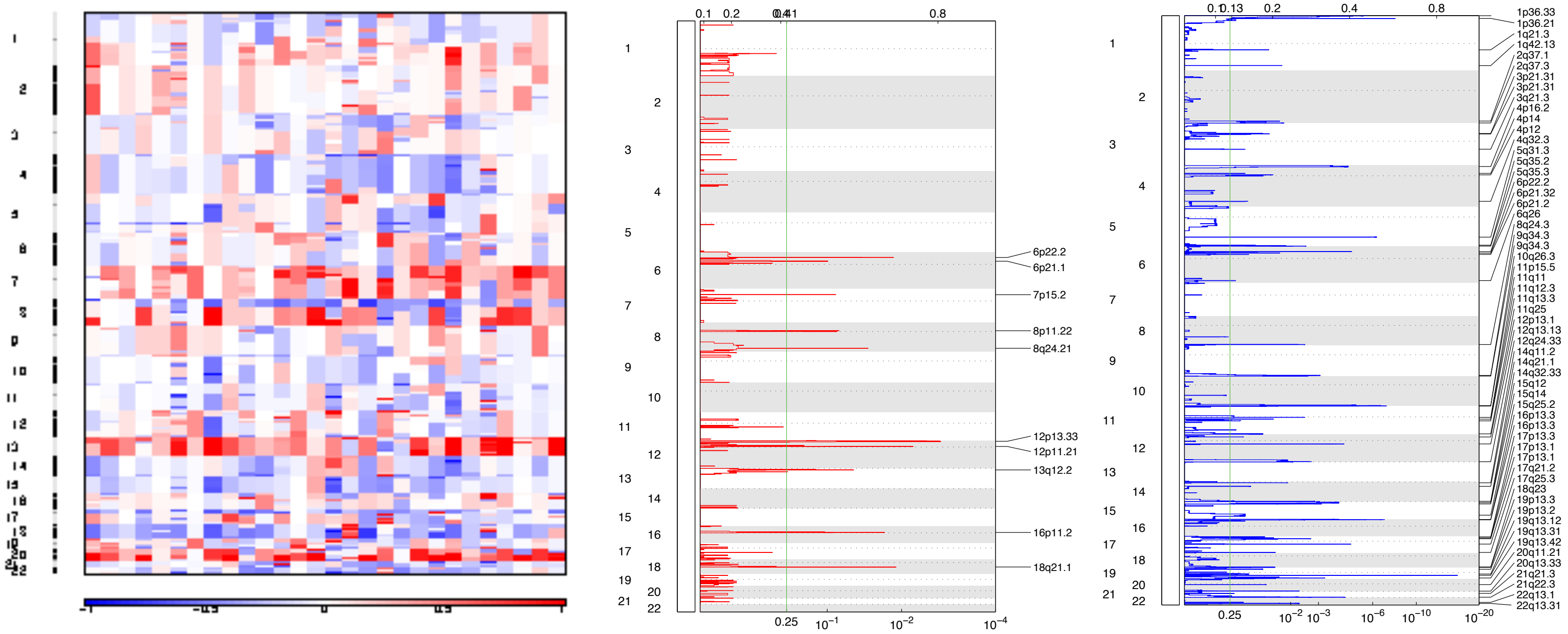


### Non-Hypermutated Rectum Samples

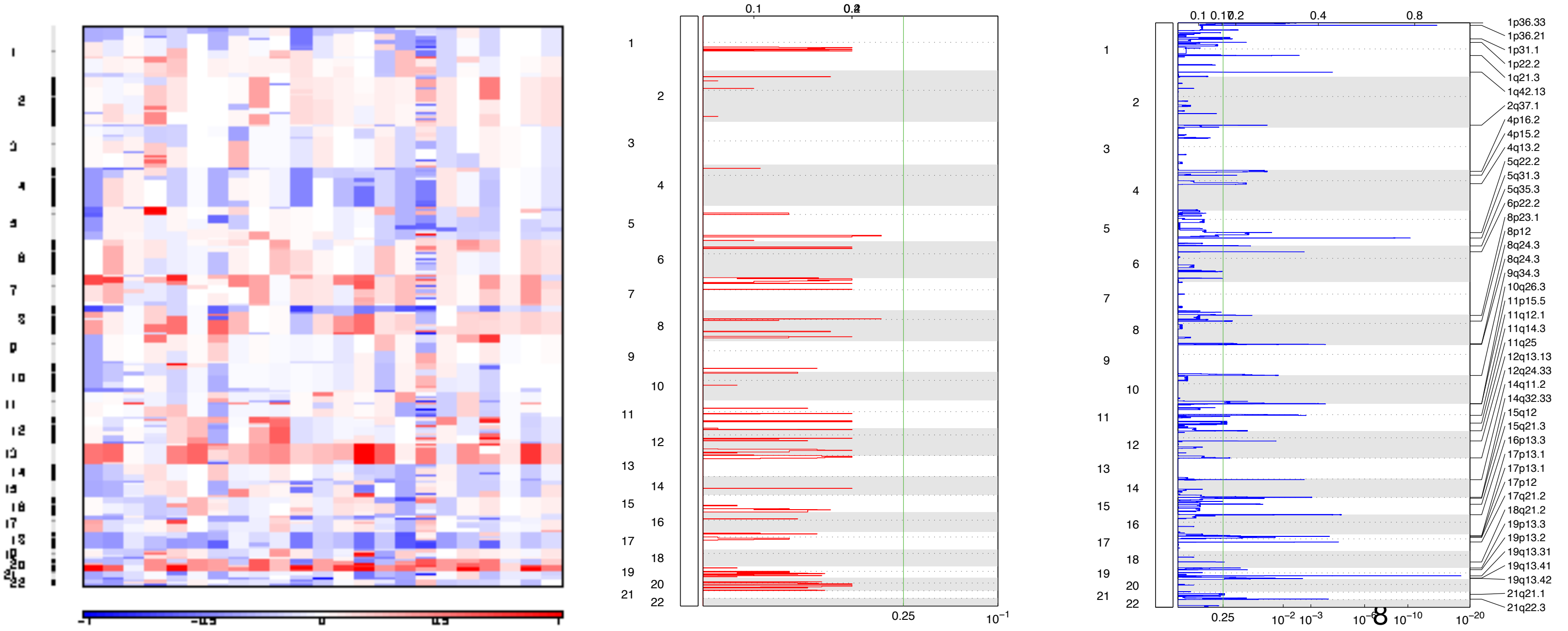

**a)**

**Supplementary Figure 1: Somatic Copy Number Alterations (SCNAs) Analysis in Non-Hypermutated Colon and Rectum Samples from Hispanic/Latino Colorectal Cancer Patients.**

This figure presents the SCNAs analysis of genomic changes in our Hispanic/Latino colorectal cancer (CRC) cohort, categorized into non-hypermutated colon and non-hypermutated rectum samples. The left panels display heatmaps of SCNAs at both chromosomal and sub-chromosomal levels, with deletions (losses) indicated in blue and insertions (gains) shown in red. The middle panels highlight focal deletions in red, while the right panels illustrate focal amplifications in blue.

**b)**

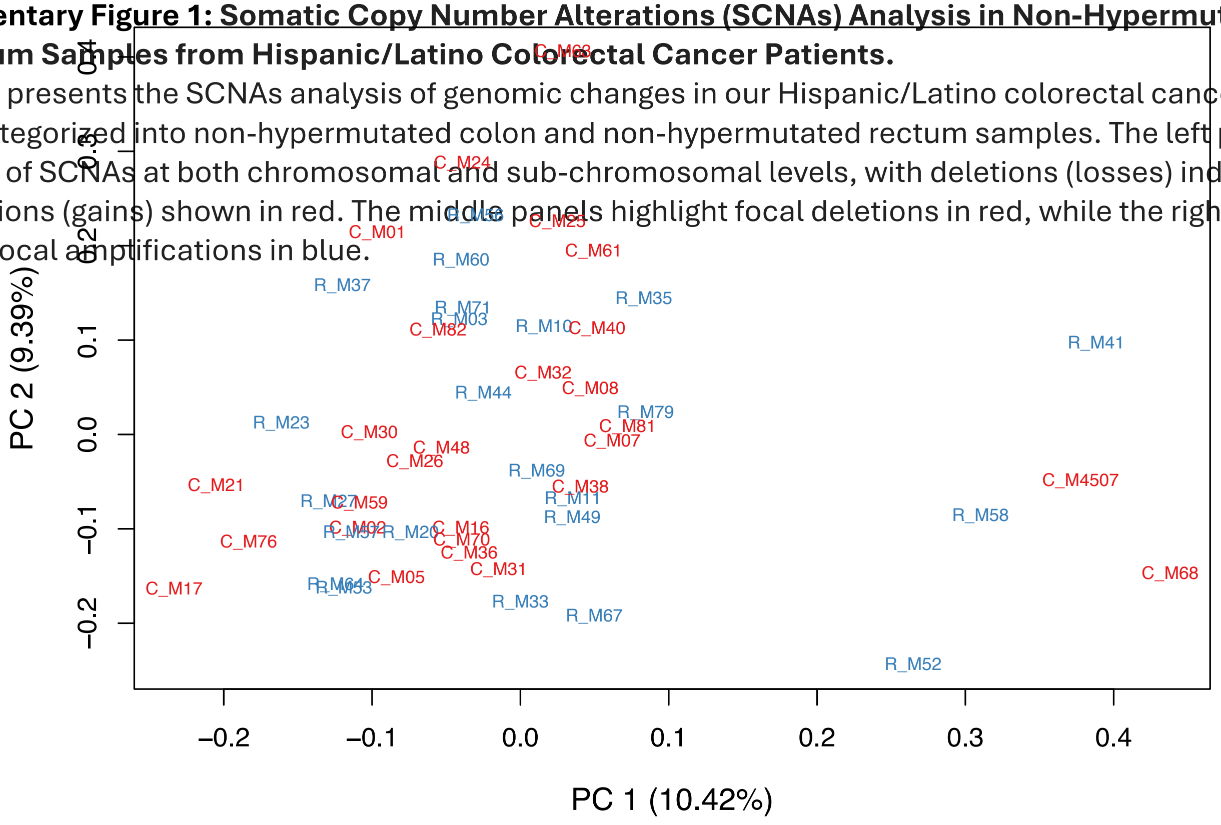


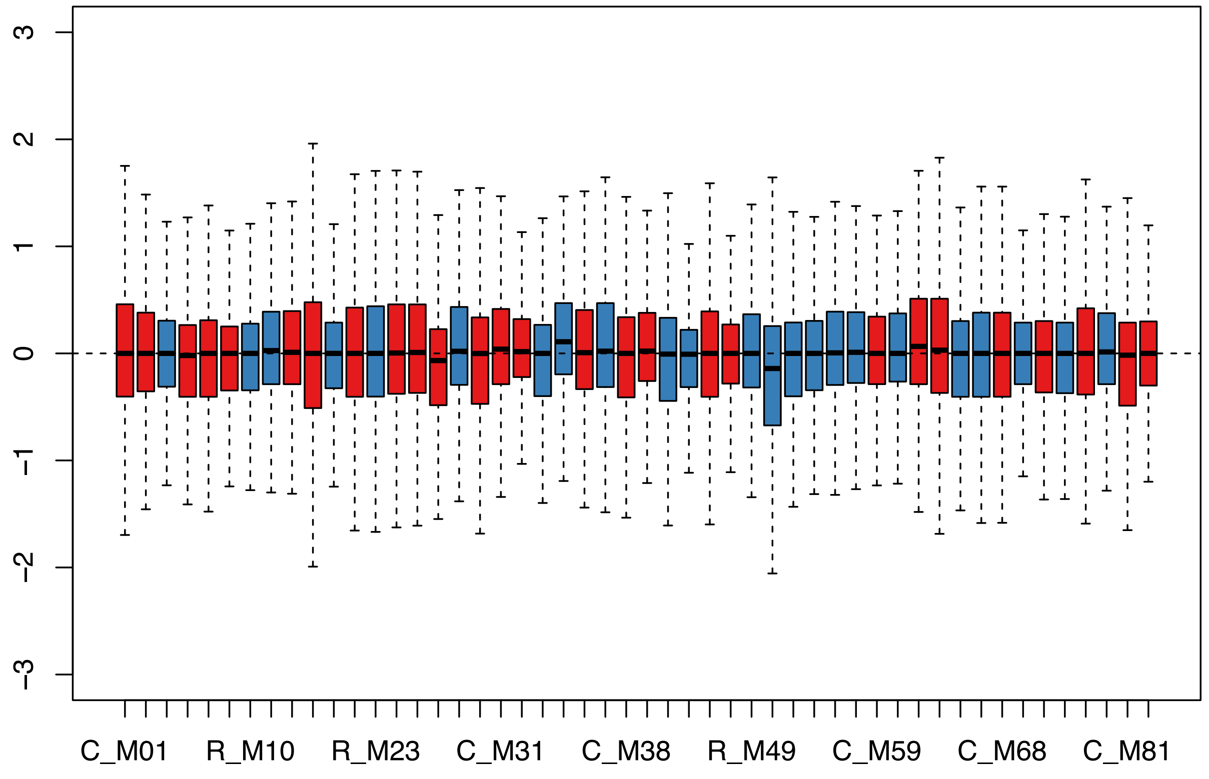

**Supplementary Table 7: Differentially Expressed CRC-Related Genes in Non-Hypermutated Colon and Non-Hypermutated Rectum Tumors Among Hispanic/Latino Patients.** This table highlights the specific colorectal cancer (CRC)-related genes that show significant differences in expression levels between non-hypermutated colon and rectum tumors in Hispanic/Latino patients.

| Gene | logFC | logCPM | LR | p-value |
|---|---|---|---|---|
| SMAD4 | -0.27 | 6.43 | 3.40 | 0.065367 |
| POLE | 0.30 | 6.74 | 3.23 | 0.072237 |
| KRAS | -0.32 | 6.18 | 2.33 | 0.127193 |
| BRAF | -0.20 | 6.89 | 2.31 | 0.128846 |
| PIK3CA | -0.19 | 5.62 | 1.98 | 0.159811 |
| MLH1 | 0.13 | 4.92 | 1.74 | 0.187624 |
| MSH6 | 0.16 | 6.03 | 1.59 | 0.207689 |
| SOX9 | -0.24 | 7.65 | 1.59 | 0.207998 |
| RB1 | -0.17 | 6.04 | 1.21 | 0.271250 |
| MUTYH | -0.15 | 4.06 | 1.21 | 0.271699 |

**Supplementary Figure 2: Differential Gene Expression (DGE) Analysis in Non-Hypermutated Colon and Non-Hypermutated Rectum Tumors Among Hispanic/Latino Patients.** This figure presents a comprehensive differential gene expression analysis of colorectal cancer (CRC) tumors from Hispanic/Latino patients, categorized into non-hypermutated colon and rectum groups. **a) Principal Component Analysis (PCA) Plot:** This plot displays the distribution of samples along two principal components, PC1 and PC2, capturing the majority of the variance in the genetic data. Colon samples are represented in red, while rectum samples are shown in blue. **b) Relative Log Expression (RLE) Plot:** This diagnostic plot assesses the quality and normalization of gene expression data, ensuring consistent expression levels across non-hypermutated colon (red) and non-hypermutated rectum (blue) tumor samples.

**Supplementary Table 8. Mutation frequencies across CRC projects.** This table includes gene names, mutation frequencies from the current Moonshot study (N=67), and frequencies from external studies: Innocenti et al. 2023-JCO (N= 540), Innocenti et al. 2023-JCO NHW (N= 463), Nunes et al. 2024-Nature (N= 1063). The included p-values demonstrate the significance levels when comparing frequencies with our study.

| | Moonshot Study (N = 67) | Innocenti et al. 2023-JCO (N = 540) | | Innocenti et al. 2023-JCO (N = 463), NHW | | Nunes et al. 2024-Nature (N =1063) | |
|---|---|---|---|---|---|---|---|
| **Gene** | **Mutated *n* (%)** | **Mutated *n* (%)** | **p-value** | **Mutated *n* (%)** | **p-value** | **Mutated *n* (%)** | **p-value** |
| **APC** | 54 (80.60%) | 355 (65.70%) | **0.0142** | 300 (64.58%) | **0.0103** | 785 (73.85%) | 0.2204 |
| **TP53** | 40 (59.70%) | 368 (68.10%) | 0.1673 | 312 (67.39%) | 0.2132 | 639 (60.11%) | 0.9468 |
| **KRAS** | 34 (50.75%) | 159 (29.40%) | **0.0004** | 125 (27.00%) | **7.347E-05** | 469 (44.12%) | 0.2899 |
| **SYNE1** | 26 (38.81%) | 0 (NA) | 1 | 0 (NA) | 1 | 0 (NA) | 1 |
| **TCF7L2** | 25 (37.31%) | 0 (NA) | 1 | 0 (NA) | 1 | 158 (14.86%) | **1.312E-06** |
| **OBSCN** | 19 (28.36%) | 0 (NA) | 1 | 0 (NA) | 1 | 0 (NA) | 1 |
| **PIK3CA** | 19 (28.36%) | 91 (16.90%) | **0.0217** | 83 (17.93%) | **0.0429** | 250 (23.52%) | 0.3670 |
| **SACS** | 18 (26.87%) | 0 (NA) | 1 | 0 (NA) | 1 | 0 (NA) | 1 |
| **AHNAK2** | 18 (26.87%) | 0 (NA) | 1 | 0 (NA) | 1 | 0 (NA) | 1 |
| **FAT3** | 17 (25.37%) | 37 (6.90%) | **5.793E-07** | 33 (7.13%) | **1.792E-06** | 0 (NA) | 1 |
| **FAT4** | 16 (23.88%) | 0 (NA) | 1 | 0 (NA) | 1 | 0 (NA) | 1 |
| **BRCA2** | 12 (17.91%) | 14 (2.60%) | **5.436E-09** | 11 (2.38%) | **5.441E-09** | 0 (NA) | 1 |
| **MSH3** | 10 (14.93%) | 0 (NA) | 1 | 0 (NA) | 1 | 0 (NA) | 1 |
| **SOX9** | 10 (14.93%) | 39 (7.20%) | **0.0283** | 31(6.70%) | **0.0184** | 160 (15.05%) | 0.9776 |
| **PTEN** | 10 (14.93%) | 20 (3.70%) | **0.0001** | 18 (3.89%) | **0.0002** | 93 (8.75%) | 0.0884 |
| **POLE** | 8 (11.94%) | 15 (2.80%) | **0.0002** | 12 (2.59%) | **0.0002** | 0 (NA) | 1 |
| **FBXW7** | 7 (10.45%) | 54 (10.00%) | 0.908 | 52 (11.23%) | 0.8489 | 191 (17.97%) | 0.1163 |
| **NF1** | 7 (10.45%) | 11 (2.00%) | **0.0001** | 9 (1.94%) | **0.0001** | 0 (NA) | 1 |
| **MLH1** | 7 (10.45%) | 4 (0.70%) | **1.203E-08** | 4 (0.86%) | **2.702E-07** | 0 (NA) | 1 |
| **MSH6** | 7 (10.45%) | 11 (2.00%) | **0.0001** | 10 (2.16%) | **0.0003** | 0 (NA) | 1 |
| **SMAD4** | 7 (10.45%) | 70 (13.00%) | 0.5546 | 17 (12.31%) | **0.0127** | 131 (12.32%) | 0.6492 |
| **BRAF** | 6 (8.96%) | 84 (15.60%) | **0.1496** | 74 (15.98%) | 0.1331 | 245 (23.05%) | **0.0071** |
| **RB1** | 5 (7.46%) | 7 (1.30%) | **0.0006** | 7 (1.51%) | **0.0022** | 0 (NA) | 1 |
| **MSH2** | 5 (7.46%) | 1 (0.20%) | **1.816E-08** | 1 (0.22%) | **1.602E-07** | 0 (NA) | 1 |
| **TET2** | 5 (7.46%) | 12 (2.20%) | **0.0135** | 10 (2.16%) | **0.0144** | 0 (NA) | 1 |
| **POLD1** | 4 (5.97%) | 6 (1.10%) | **0.0031** | 6 (1.13%) | **0.0086** | 0 (NA) | 1 |
| **MUTYH** | 2 (2.99%) | 2 (0.40%) | **0.0154** | 2 (0.043%) | **0.0240** | 0 (NA) | 1 |

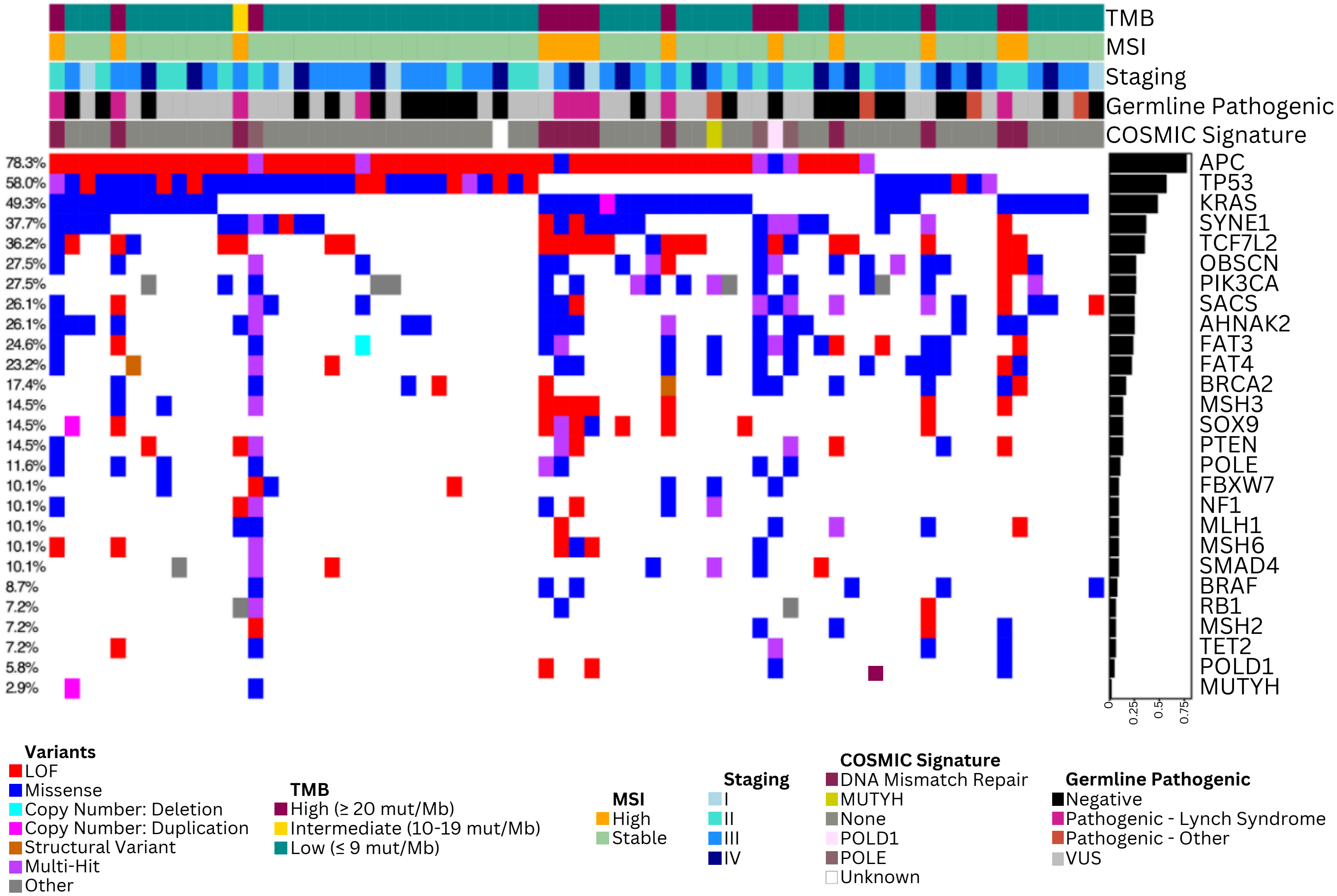


**Supplementary Figure 3: Oncoplot of Our 67 CRC H/L Patient Cohort.** This oncoplot identifies CRC-related genes and somatic mutations within our cohort of 67 Hispanic/Latino colorectal cancer (CRC) patients. The top panel displays the total mutational burden (TMB), microsatellite stability (MSI), cancer staging, germline pathogenic mutations, and COSMIC signatures calculated for each sample. The middle panel highlights CRC-related genes, categorized by the type of mutation observed. The bottom panel provides legends for the middle and upper panels, detailing the color codes used.

Hypermutated Samples

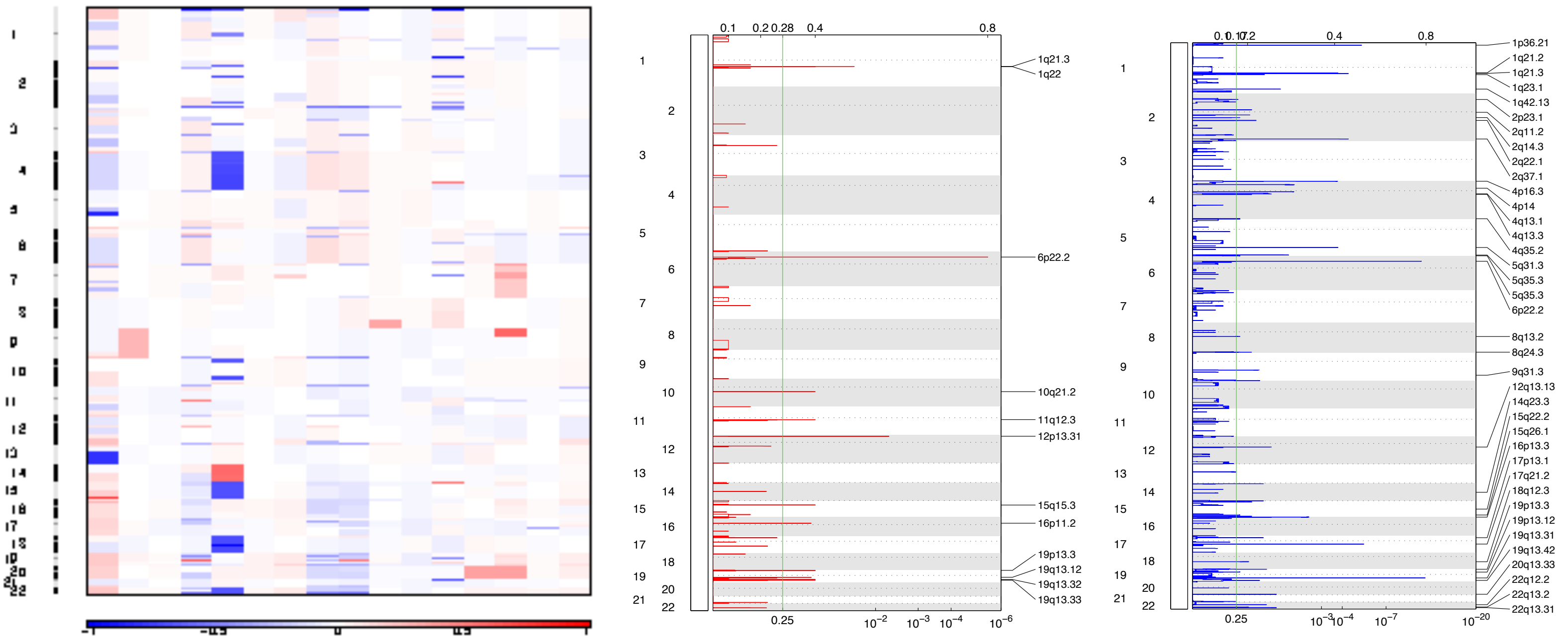


Non-Hypermutated Samples

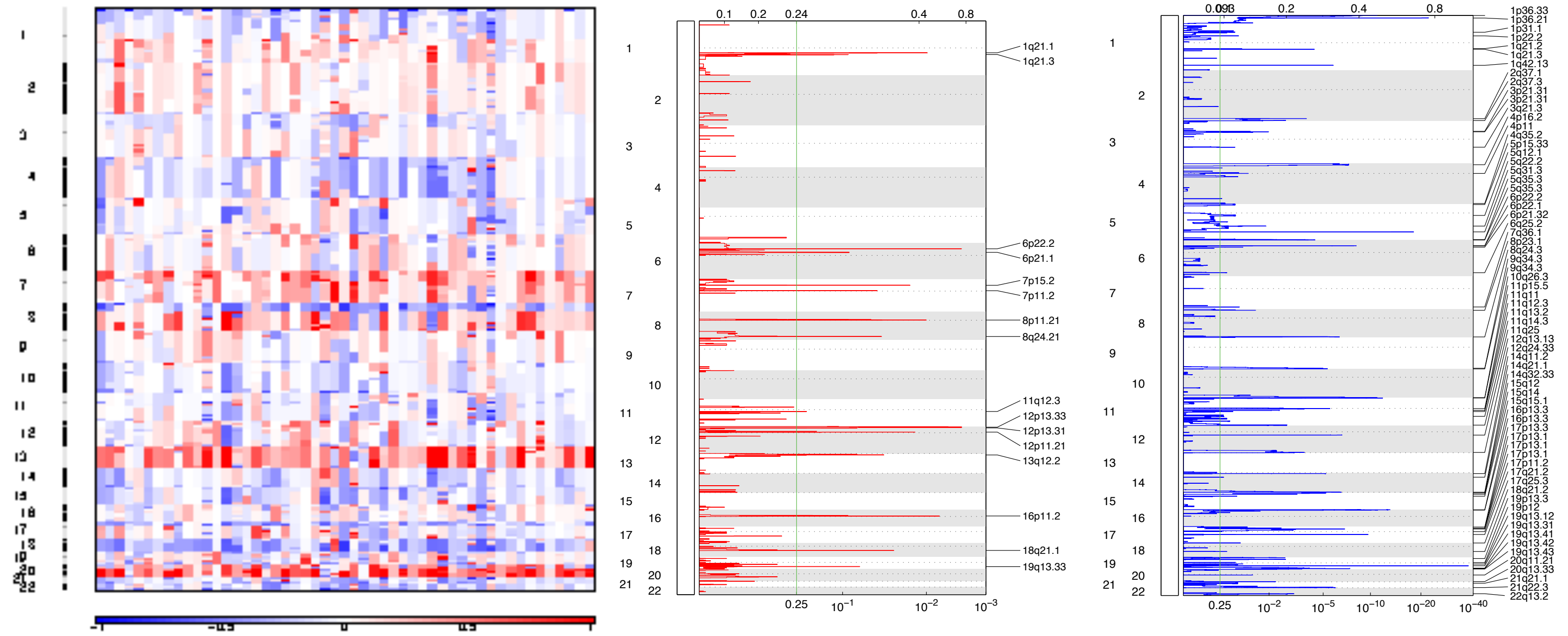


**Supplementary Figure 4: Somatic Copy Number Alterations (SCNAs) Analysis of the hypermutated and non-hypermutated samples from the 67 Hispanic/Latino Colorectal Cancer Patients.** This figure presents the SCNAs analysis of genomic changes in 67 Hispanic/Latino colorectal cancer (CRC) patients, categorized into hypermutated and non-hypermutated samples. The left panels display heatmaps of SCNAs at both chromosomal and sub-chromosomal levels, with deletions (losses) indicated in blue and insertions (gains) shown in red. The middle panels show focal deletions in red, while the right panels display focal amplifications in blue.

Hypermutated MSI Samples

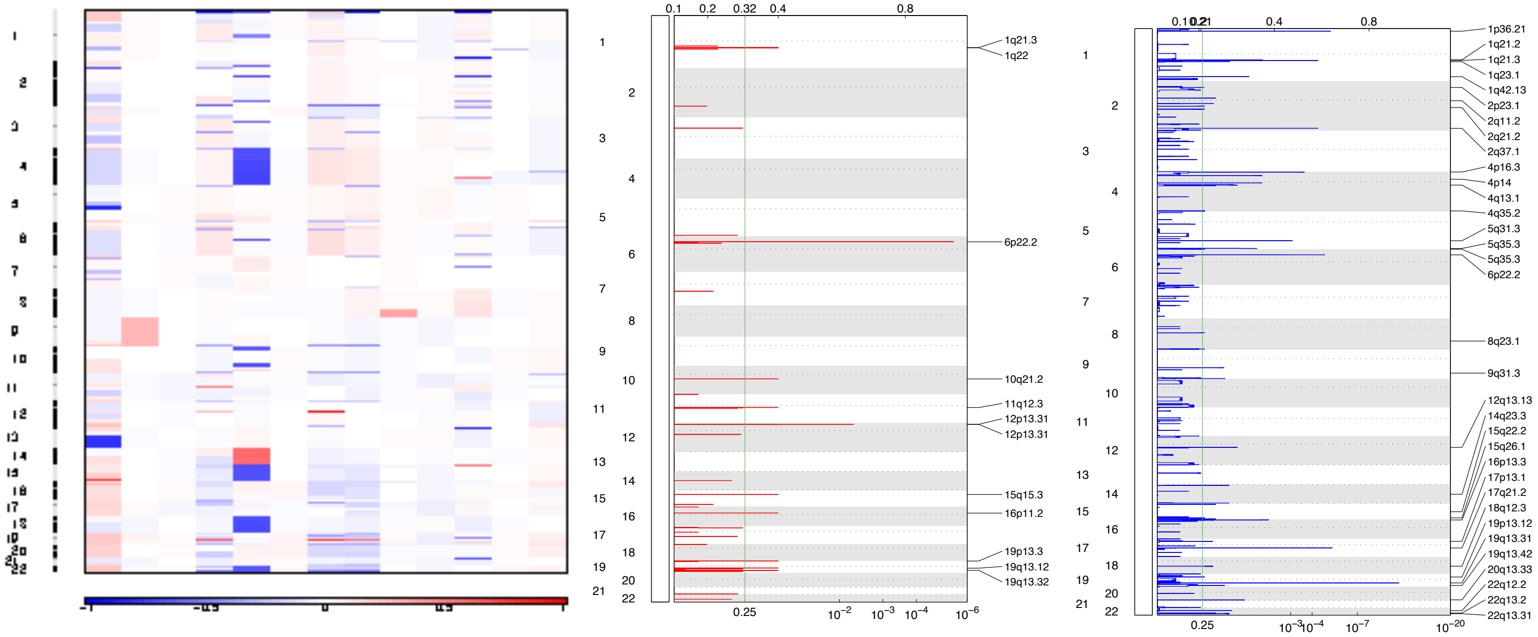


Hypermutated MSS Samples

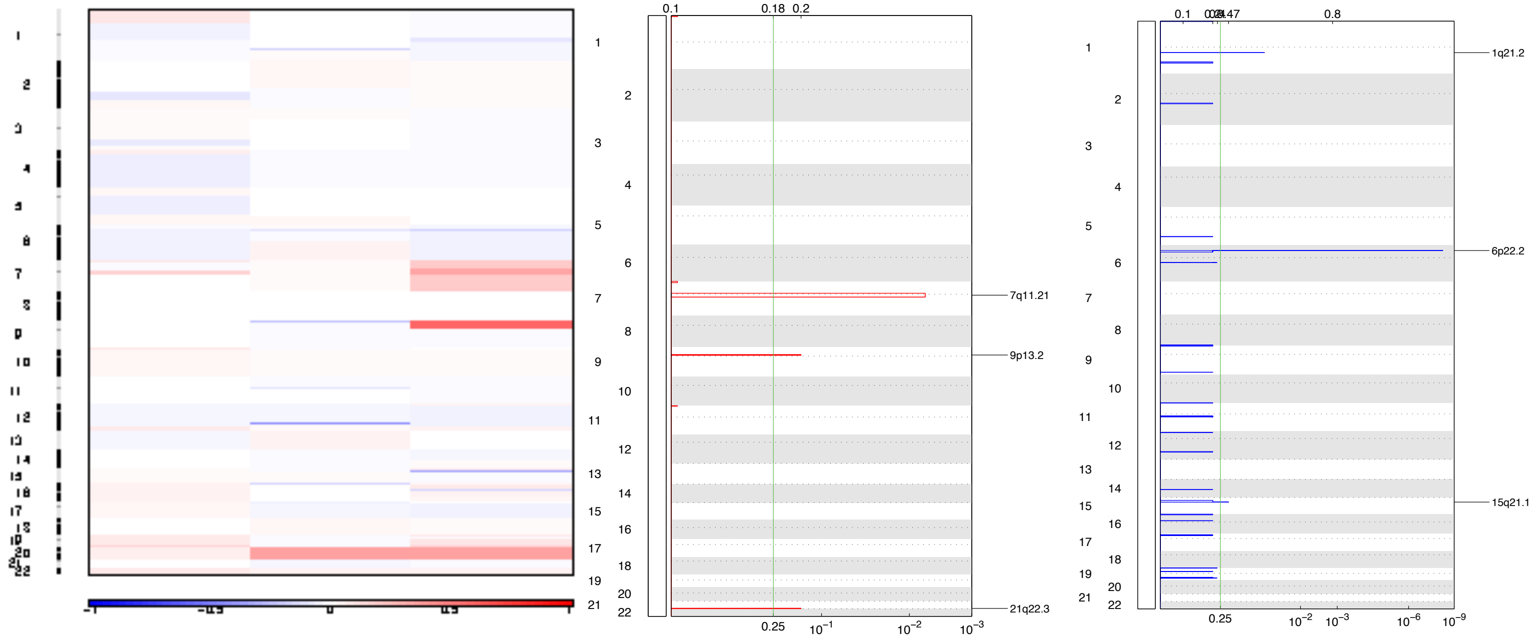


**Supplementary Figure 5: Somatic Copy Number Alterations (SCNAs) Analysis in Hypermutated MSI and Hypermutated MSS Samples from Hispanic/Latino Colorectal Cancer Patients.** This figure presents the SCNAs analysis of genomic changes in our Hispanic/Latino colorectal cancer (CRC) cohort, categorized into hypermutated MSI (microsatellite instability) and hypermutated MSS (microsatellite stable) samples. The left panels display heatmaps of SCNAs at both chromosomal and sub-chromosomal levels, with deletions (losses) indicated in blue and insertions (gains) shown in red. The middle panels highlight focal deletions in red, while the right panels illustrate focal amplifications in blue.

≤55% 1KG-PEL-like Samples

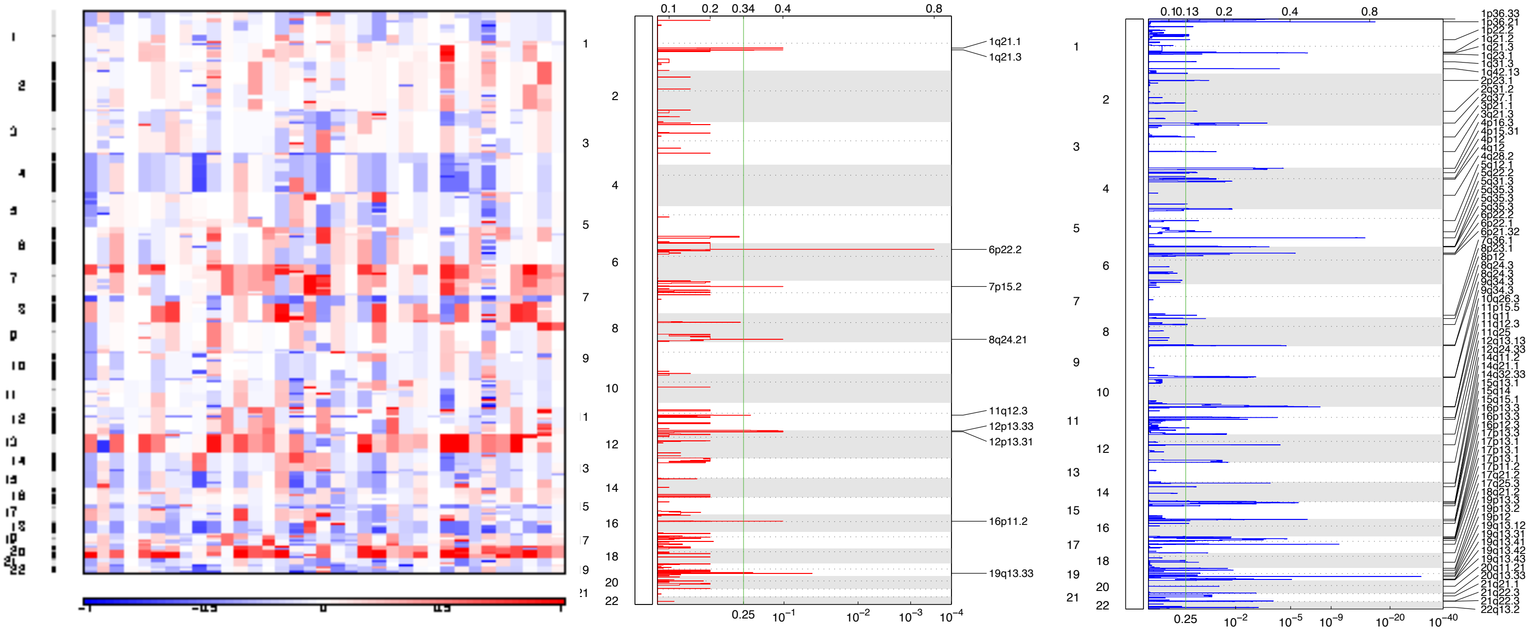


>55% 1KG-PEL-like Samples

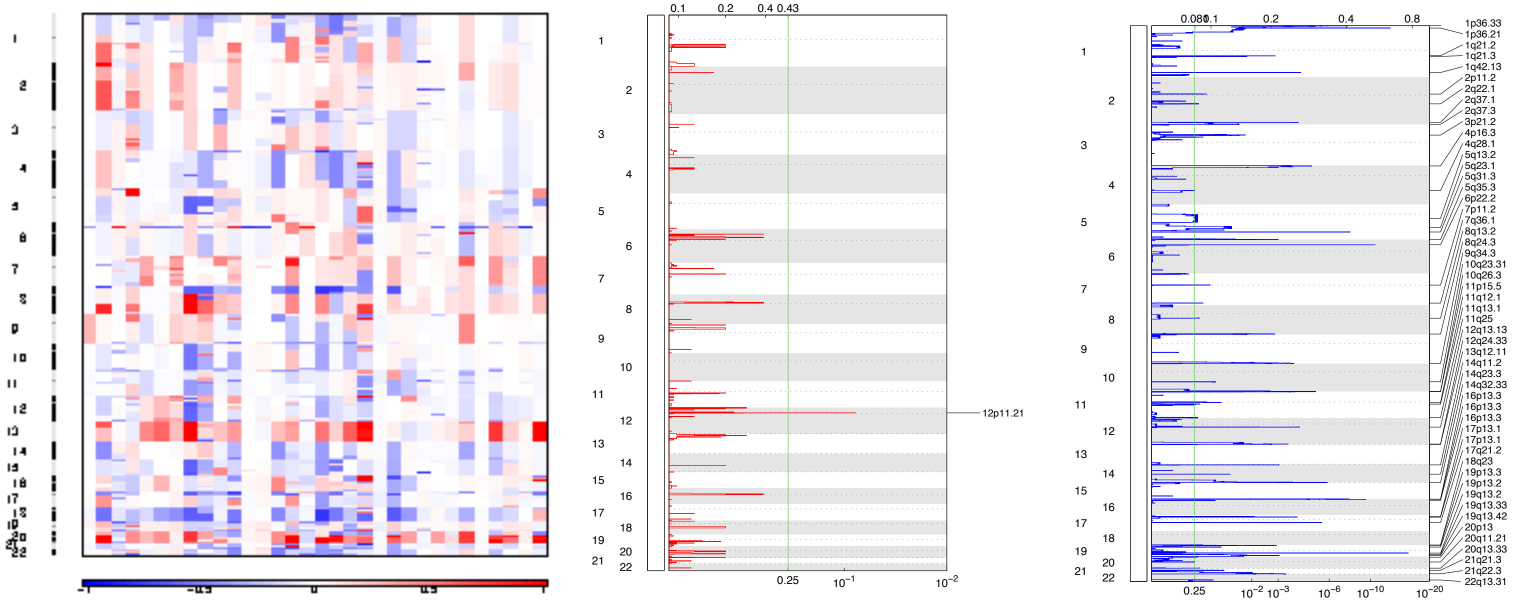


**Supplementary Figure 6: Somatic Copy Number Alterations (SCNAs) Analysis Based on 1KG-PEL-like Proportion in Hispanic/Latino Colorectal Cancer Patients.** This figure presents the SCNAs analysis of genomic changes in our Hispanic/Latino colorectal cancer (CRC) cohort, categorized into samples with ≤ 55% 1000 Genomes Project Peruvian-in-Lima-like (1KG-PEL-like) proportion and samples with >55% 1KG-PEL-like proportion. The left panels display heatmaps of SCNAs at both chromosomal and sub-chromosomal levels, with deletions (losses) indicated in blue and insertions (gains) shown in red. The middle panels highlight focal deletions in red, while the right panels illustrate focal amplifications in blue.

**Supplementary Table 9: Chromosomal and sub-chromosomal aberrations in 67 Hispanic/Latino CRC Patients.** This table presents the results of a visual inspection of chromosomal and sub-chromosomal aberrations in our H/L CRC cohort. Amplifications are indicated in blue, while deletions are shown in orange.

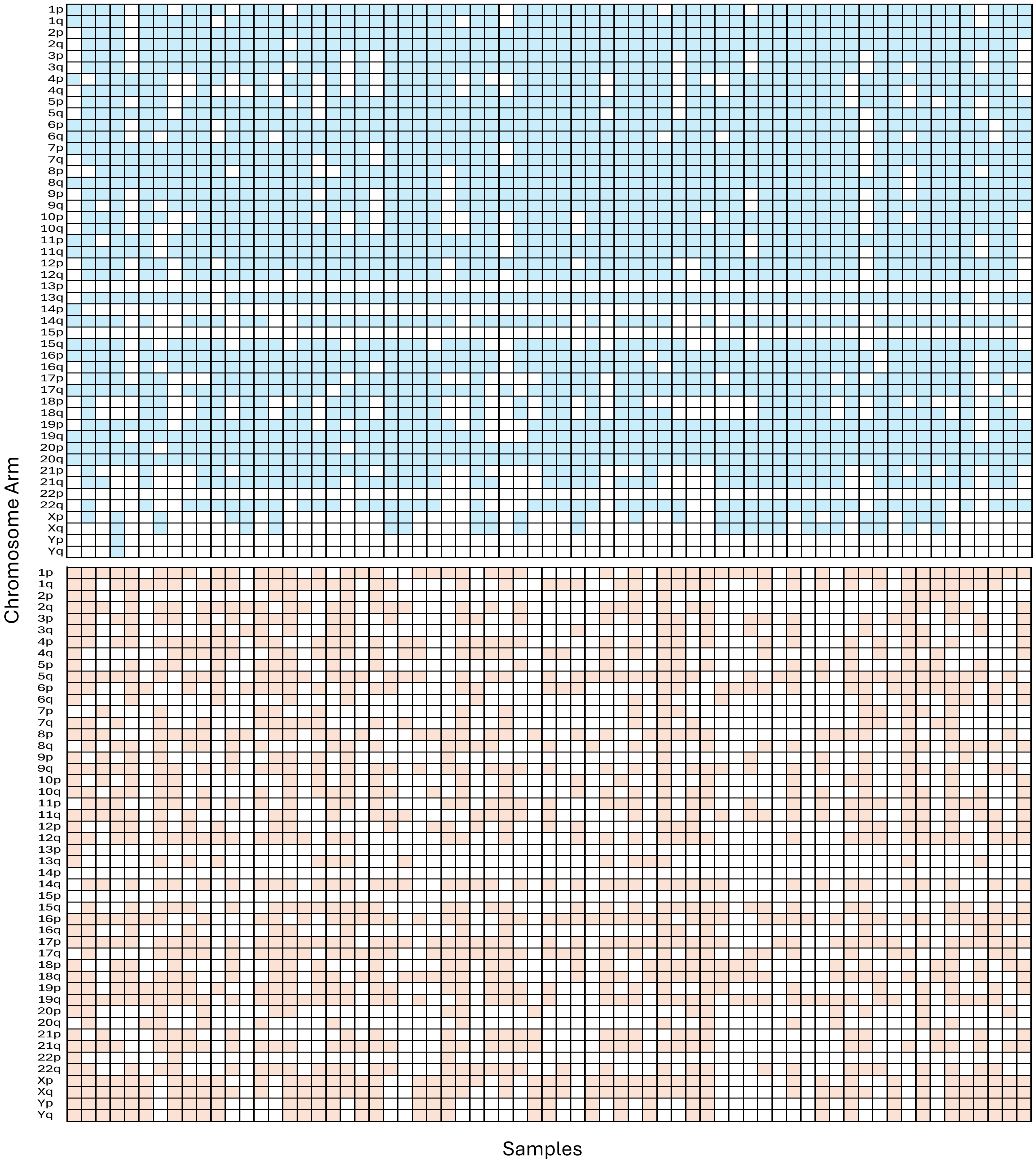

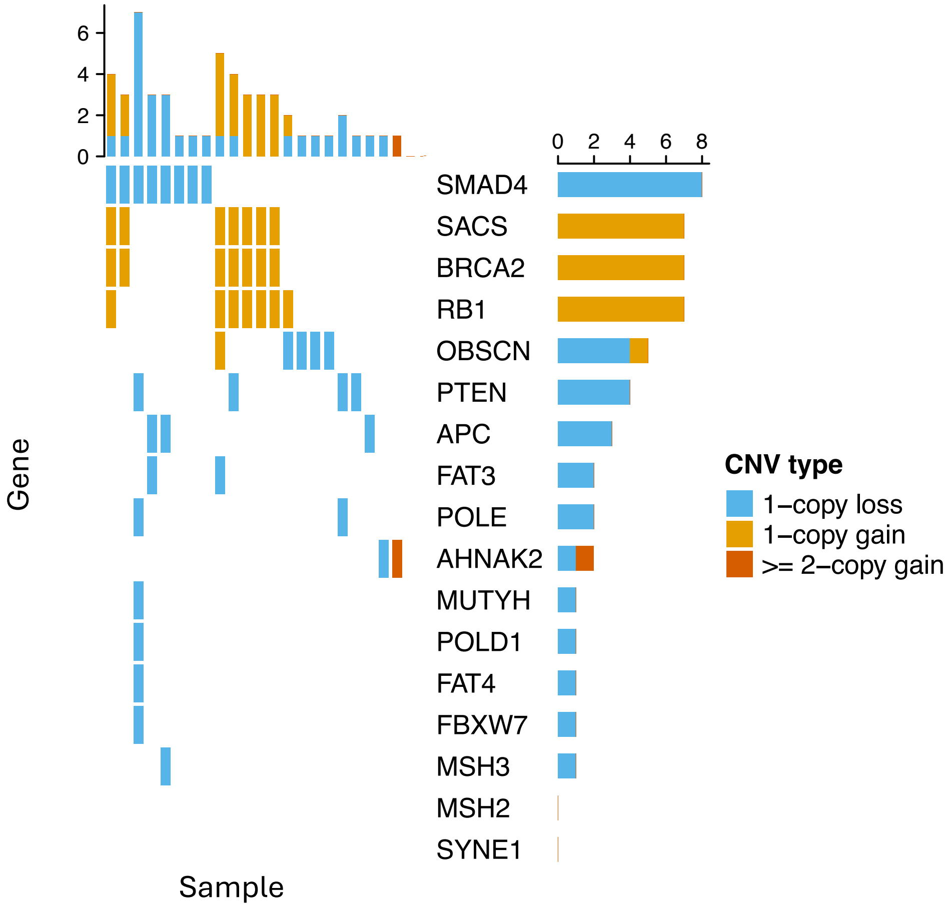


**Supplementary Figure 7: SCNAs in Selected CRC-Related Genes in 67 Hispanic/Latino CRC Patients.** This figure illustrates somatic copy number alterations (SCNAs) in selected colorectal cancer (CRC)-related genes in 67 Hispanic/Latino CRC patients. The right panel features a bar graph showing the genes and the number of samples affected.

**Supplementary Table 10: Focal Deletions and Amplifications Identified from SCNAs in Selected CRC-Related Genes in 67 Hispanic/Latino CRC Patients.** This table presents recurrent **a)** deletion peaks and **b)** amplification peaks identified from somatic copy number alterations (SCNAs) in selected colorectal cancer (CRC)-related genes in 67 Hispanic/Latino CRC patients. The data includes the cytoband location, residual q-values, genes within the peak, CRC related genes, wide peak boundaries, and genes in the peak. (**Reference Supplementary Table 10 Excel File for complete table)**

**a) Focal Deletions**

| Cytoband | Residual q Value | Genes in Peak | CRC Related | Wide Peak Boundaries | Genes in Peak |
|---|---|---|---|---|---|
| 19q13.31 | 6.25E-48 | 3 | None | chr19:42932592-43199470 | PSG2, PSG5, PSG11 |
| 1p36.21 | 1.77E-27 | 14 | None | chr1:12820027-13125204 | PRAMEF2, HNRNPCL1, PRAMEF10, PRAMEF4, PRAMEF11, PRAMEF6, HNRNPCL2, PRAMEF7, PRAMEF25, PRAMEF26, |
| 5q31.3 | 1.62E-23 | 4 | None | chr5:141005324-141113859 | PCDHB1, PCDHB3, PCDHB2, LOC101926905 |
| 6p22.2 | 1.58E-16 | 8 | None | chr6:26098570-26198085 | HIST1H1E, HIST1H1T, HIST1H2BD, HIST1H2AC, HIST1H2BE, HIST1H2BC, HIST1H4D, HIST1H4C |
| 17q21.2 | 1.65E-15 | 25 | None | chr17:41034219-41308737 | KRTAP1-1, KRTAP9-9, KRTAP4-6, KRTAP2-1, KRTAP4-12, KRTAP9-2, KRTAP9-3, KRTAP9-8, KRTAP4-4, KRTAP9-4, KRTAP4 |
| 11p15.5 | 1.12E-13 | 1 | None | chr11:1033176-1077667 | LOC101927503 |
| 10q26.3 | 9.81E-13 | 11 | None | chr10:132784817-133263055 | UTF1, VENTX, CFAP46, ADGRA1, KNDC1, LINC01168, MIR202, ADGRA1-AS1, LINC01166, MIR202HG, LINC01167 |
| 16p13.3 | 4.03E-11 | 20 | None | chr16:731803-1356583 | SSTR5, TPSAB1, UBE2I, CACNA1H, BAIAP3, MSLN, TPSD1, TPSG1, SOX8, GNG13, CHTF18, TPSB2, LMF1, RPUSD1, TSF |
| 4p16.3 | 9.26E-11 | 5 | None | chr4:1794144-2043171 | LETM1, WHSC1, NELFA, SCARNA22, MIR943 |
| 1p36.33 | 5.49E-10 | 75 | None | chr1:1-1638262 | DVL1, SCNN1D, TNFRSF4, MMP23B, MMP23A, TNFRSF18, ISG15, NOC2L, SSU72, SDF4, MXRA8, CPSF3L, C1orf159, |
| 17p13.1 | 1.87E-09 | 2 | None | chr17:10472011-10534871 | MYH1 |
| 12q13.13 | 2.29E-09 | 15 | None | chr12:52474096-52897327 | KRT1, KRT2, KRT3, KRT4, KRT5, KRT6A, KRT76, KRT71, KRT74, KRT72, KRT78, KRT73, KRT79, KRT77, KRT73-AS1 |
| 1q21.3 | 5.92E-09 | 32 | None | chr1:152604080-153223734 | IVL, SMCP, SPRR1A, SPRR1B, SPRR2A, SPRR2B, SPRR2C, SPRR2D, SPRR2E, SPRR2F, SPRR2G, SPRR3, LCE2B, LELP1, |
| 14q32.33 | 6.47E-09 | 15 | None | chr14:105490707-107043718 | ELK2AP, ADAM6, FAM30A, TMEM121, C14orf80, LINC00226, LINC00221, MIR4507, MIR4538, MIR4539, MIR4537, MIF |
| 1q42.13 | 1.04E-08 | 2 | OBSCN | chr1:228175077-228394604 | OBSCN, C1orf145 |
| 8q24.3 | 6.77E-08 | 28 | None | chr8:141405130-143268781 | ADGRB1, CYP11B1, CYP11B2, GML, LY6E, LY6H, PSCA, LY6D, JRK, PTP4A3, ARC, THEM6, LY6K, SLURP1, LYNX1, LYPD |
| 14q11.2 | 6.77E-08 | 3 | None | chr14:23383421-23434386 | MIR208A, MIR208B, MHRT |
| 2q37.1 | 2.13E-07 | 5 | None | chr2:232379416-232526366 | ALPI, ALPPL2, ECEL1, ECEL1P2, PRSS56 |
| 19q13.42 | 8.40E-07 | 21 | None | chr19:54503836-54874189 | KIR2DL1, KIR2DL2, KIR2DL3, KIR2DL4, KIR2DS1, KIR2DS3, KIR2DS4, KIR2DS5, KIR3DL1, KIR3DL2, KIR3DS1, LILRB1, L |
| 11q11 | 9.62E-06 | 7 | None | chr11:55545865-55783070 | OR4P4, OR4C15, OR4C16, OR4C11, OR4S2, OR4C6, OR5D13 |
| 15q13.1 | 9.62E-06 | 15 | None | chr15:27524123-29123362 | APBA2, OCA2, HERC2, GOLGA8G, HERC2P9, WHAMMP2, PDCD6IPP2, GOLGA8M, GOLGA6L7P, GOLGA8F, LOC1002 |
| 12q24.33 | 1.38E-05 | 30 | POLE | chr12:131846069-133275309 | GOLGA3, POLE, PXMP2, ZNF10, ZNF26, ZNF84, ZNF140, ULK1, ZNF268, P2RX2, ANKLE2, GALNT9, CHFR, EP400, FBRS |
| 22q13.2 | 1.38E-05 | 23 | None | chr22:43275668-45292584 | MPPED1, NUP50, KIAA0930, ARHGAP8, SAMM50, SULT4A1, PARVB, PRR5, PARVG, EFCAB6, PNPLA3, LDOC1L, KIAA16 |
| 16p13.3 | 3.85E-05 | 12 | None | chr16:2036977-2168522 | NTHL1, PKD1, TSC2, RAB26, SNORD60, MIR1225, MIR3180-5, SNHG19, MIR4516, MIR6511B1, MIR6511B2, LOC105 |
| 5q35.3 | 7.99E-05 | 3 | None | chr5:180590355-180830768 | FLT4, MGAT1, OR2Y1 |
| 19p13.3 | 7.99E-05 | 3 | None | chr19:1250297-1286407 | CIRBP, C19orf24, CIRBP-AS1 |
| 20q13.33 | 7.99E-05 | 7 | None | chr20:63265854-63520994 | CHRNA4, EEF1A2, KCNQ2, ARFGAP1, COL20A1, LOC100130587, MIR4326 |
| 1q21.2 | 0.00011897 | 12 | None | chr1:149701747-149919753 | FCGR1A, HIST2H2AA3, HIST2H2AC, HIST2H2BE, HIST2H4A, SV2A, BOLA1, HIST2H2AB, HIST2H3A, HIST2H2BC, HIST2 |
| 9q34.3 | 0.00012629 | 42 | None | chr9:134433454-136429550 | COL5A1, FCN1, FCN2, LCN1, PAEP, SNAPC4, LHX3, PPP1R26, UBAC1, OLFM1, SDCCAG3, PMPCA, GPSM1, DKFZP43 |
| 21q22.3 | 0.0002128 | 10 | None | chr21:44568106-44663749 | KRTAP10-10, KRTAP10-4, KRTAP10-6, KRTAP10-7, KRTAP10-9, KRTAP10-11, KRTAP10-5, KRTAP10-8, KRTAP12-3, KRTAF |
| 21q22.3 | 0.00062412 | 13 | None | chr21:45341257-46236977 | COL6A1, COL6A2, LSS, SLC19A1, FTCD, PCBP3, COL18A1, SPATC1L, COL18A1-AS1, LOC100129027, COL18A1-AS2, L |
| 17p13.1 | 0.0009865 | 11 | None | chr17:7218382-7314033 | ACADVL, CLDN7, DVL2, EIF5A, SLC2A4, GABARAP, CTDNEP1, ELP5, YBX2, PHF23, MIR324 |
| 3p21.31 | 0.0014881 | 2 | None | chr3:50348625-50561846 | CACNA2D2, CYB561D2, TMEM115 |
| 19p13.2 | 0.0014881 | 8 | None | chr19:8821081-9193547 | ZNF317, MBD3L1, MUC16, OR7G1, OR1M1, OR7D2, OR7G2, OR7G3 |
| 9q34.3 | 0.0019733 | 8 | None | chr9:137815180-138394717 | CACNA1B, TUBBP5, MIR602, FAM157B, LOC100133077, LOC101928786, LOC101928932, LOC105376331 |
| 5q22.2 | 0.0022709 | 77 | APC | chr5:103520650-122066782 | hsa-mir-1244-2, APC, CAMK4, CDO1, AP3S1, DMXL1, EFNA5, FER, HSD17B4, KCNN2, MAN2A1, MCC, PGGT1B, SRP1 |
| 5q35.3 | 0.0041653 | 7 | None | chr5:178817885-179559864 | GRM6, ADAMTS2, ZNF354C, ZFP2, ZNF354B, ZNF454, ZNF879 |
| 11q25 | 0.0041653 | 23 | None | chr11:130846540-135086622 | OPCML, IGSF9B, NCAPD3, ACAD8, B3GAT1, THYN1, NTM, JAM3, GLB1L2, VPS26B, GLB1L3, SPATA19, MIR4697HG, LC |
| 11q12.3 | 0.0044341 | 8 | None | chr11:62885709-63509090 | CHRM1, SLC22A6, SLC22A8, SLC22A9, HRASLS5, SLC22A24, SLC22A25, SLC22A10 |
| 4q28.3 | 0.0061173 | 662 | FAT4, FBXW7, | chr4:60904894-190214555 | hsa-mir-1979, ADH1A, ADH1B, ADH1C, ADH4, ADH5, ADH6, ADH7, AFM, AFP, AGA, ALB, AMBN, ANK2, SLC25A4, ANX |
| 20q11.21 | 0.0067542 | 10 | None | chr20:32985779-33408649 | BPIFA1, CDK5RAP1, BPIFB2, BPIFB1, BPIFB6, BPIFA3, BPIFA2, BPIFB4, BPIFA4P, BPIFB3 |
| 5q35.3 | 0.0095241 | 6 | None | chr5:176996302-177321169 | FGFR4, ZNF346, PRELID1, RAB24, NSD1, MXD3 |
| 2q37.3 | 0.011757 | 12 | None | chr2:240588288-241233832 | AGXT, KIF1A, GPR35, PPP1R7, PASK, SNED1, ANO7, C2orf54, MTERF4, LOC200772, AQP12A, AQP12B |
| 6p21.32 | 0.017651 | 5 | None | chr6:33053142-33196333 | COL11A2, HLA-DPA1, HLA-DPB1, HLA-DPB2, HCG24 |
| 5q12.1 | 0.019813 | 217 | MSH3 | chr5:51354294-81972486 | TRIM23, ARSB, BHMT, BTF3, CCNB1, CDK7, CKMT2, ERCC8, CRHBP, DHFR, F2R, F2RL1, F2RL2, FOXD1, GTF2H2, GZ |
| 16p13.3 | 0.019813 | 839 | None | chr16:2088711-81235628 | hsa-mir-1826, AARS, ABAT, ABCA3, ADCY7, ADCY9, AP1G1, AGRP, ALDOA, AMFR, AQP8, ABCC6, ZFHX3, ATP2A1, ATP |
| 3q21.3 | 0.023821 | 7 | None | chr3:126955371-127602491 | PLXNA1, TPRA1, C3orf56, LOC101927123, LINC01471, MIR7976, MIR6825 |
| 6p22.1 | 0.023821 | 20 | None | chr6:27400948-27921140 | HIST1H1B, ZNF184, HIST1H2AI, HIST1H2AK, HIST1H2AJ, HIST1H2AL, HIST1H2AM, HIST1H2BL, HIST1H2BN, HIST1H2 |
| 7q36.1 | 0.023821 | 2 | None | chr7:149762591-149849047 | SSPO, ZNF467 |
| 19q13.41 | 0.023821 | 195 | None | chr19:50353541-54109275 | hsa-mir-643, KLK3, CD33, SIGLEC6, ETFB, FPR1, FPR2, FPR3, GPR32, HAS1, KLK1, KLK2, LIM2, MYBPC2, NDUFA3, NK |
| 8p23.1 | 0.033703 | 2 | None | chr8:2724913-6416553 | CSMD1, LOC100287015 |
| 5p15.33 | 0.045767 | 608 | APC, MSH3 | chr5:1-118572512 | hsa-mir-1274a, ADCY2, APC, TRIM23, ARSB, BHMT, BTF3, C6, C7, C9, CAMK4, CAST, CCNB1, CCNH, CDH6, CDH9, |
| 4q12 | 0.045767 | 5 | None | chr4:54100246-55384926 | KDR, KIT, PDGFRA, SRD5A3, LOC339978 |
| 19q13.12 | 0.045767 | 63 | None | chr19:36123090-38625129 | CAPNS1, COX7A1, PSMD8, RYR1, ZNF146, DPF1, KCNK6, SPINT2, MAP4K1, ZFP30, SIPA1L3, ZNF345, ZNF571, ZFP14, |
| 1q23.1 | 0.051806 | 3 | None | chr1:158588115-158706297 | SPTA1, OR6K2, OR10Z1 |
| 19p12 | 0.051806 | 20 | None | chr19:21763201-23415031 | ZNF43, ZNF91, ZNF99, ZNF208, ZNF492, ZNF257, ZNF98, ZNF676, LOC374890, ZNF728, GOLGA2P9, ZNF724, LINC |
| 15q14 | 0.081577 | 6 | None | chr15:33145048-34086793 | CHRM5, RYR3, AVEN, FMN1, TMCO5B, LOC101928134 |
| 2q22.1 | 0.089974 | 4 | None | chr2:138671766-143134649 | KYNU, LRP1B, YY1P2, MIR7157 |
| 17q25.3 | 0.091655 | 8 | None | chr17:81089299-81234147 | AATK, CEP131, AATK-AS1, MIR338, MIR657, MIR1250, MIR3065, LOC105371925 |
| 2p23.1 | 0.10211 | 13 | None | chr2:29162283-32020209 | ALK, SRD5A2, XDH, EHD3, MEMO1, YPEL5, GALNT14, LBH, CAPN13, LCLAT1, LOC285043, CAPN14, LOC105374389 |
| 1p31.1 | 0.10764 | 258 | None | chr1:46658274-85254559 | ACADM, AK4, C8A, C8B, CDKN2C, CPT2, CRYZ, CTBS, CTH, CYP2J2, CYP4A11, CYP4B1, DAB1, GADD45A, DHCR24, |
| 11q13.2 | 0.10764 | 3 | None | chr11:68033384-68053887 | TCIRG1, MIR4691, MIR6753 |
| 1q31.3 | 0.12622 | 11 | None | chr1:193230957-197159497 | F13B, CFH, CFHR1, CFHR2, CFHR4, CFHR3, CFHR5, ASPM, KCNT2, MIR4735, LINC01031 |
| 8q13.2 | 0.12622 | 3 | None | chr8:67726272-68920862 | PREX2, C8orf34, C8orf34-AS1 |
| 4p12 | 0.13934 | 3 | None | chr4:44673423-46244093 | GABRG1, GUF1, GNPDA2 |
| 8q24.3 | 0.13934 | 11 | None | chr8:143691498-143831749 | PUF60, SCRIB, MAPK15, BREA2, FAM83H, MIR937, FAM83H-AS1, CCDC166, MIR4664, LOC101928160, LOC10537 |
| 2q31.2 | 0.15721 | 4 | None | chr2:178501277-179109798 | TTN, CCDC141, TTN-AS1, LOC101927055 |
| 15q15.1 | 0.15721 | 51 | None | chr15:41480426-43786301 | CAPN3, CKMT1B, EPB42, GANC, PDIA3, ITPKA, LTK, MAP1A, TP53BP1, TYRO3, JMJD7-PLA2G4B, SNAP23, TGM5, PPIP5 |
| 16p12.3 | 0.15721 | 7 | None | chr16:20308155-20736779 | GP2, UMOD, ACSM5, ACSM1, ACSM2A, PDILT, ACSM2B |
| 6p21.2 | 0.17291 | 47 | None | chr6:37168784-41741180 | DNAH8, GLO1, GLP1R, MDFI, MOCS1, NFYA, PIM1, TFEB, KCNK5, RNF8, NCR2, APOBEC2, CMTR1, DAAM2, TREM2, T |
| 2p11.2 | 0.22813 | 1740 | MSH6, MSH2 | chr2:1-242193529 | hsa-mir-1244-1, hsa-mir-3130-3, hsa-mir-1245, AAMP, ACADL, ACP1, ACTG2, ACVR1, ACVR2A, ACYP2, ADCY3, ADD |
| 18q21.2 | 0.22813 | 410 | SMAD4 | chr18:1-80373285 | ADCYAP1, AQP4, ATP5A1, BCL2, LDLRAD4, CDH2, CDH7, CETN1, CIDEA, CYB5A, DCC, DSC1, DSC2, DSC3, DSG1, D |
| 6q25.2 | 0.22813 | 1400 | SYNE1 | chr6:1-170805979 | hsa-mir-2113, ABCF1, ACAT2, CRISP1, AGER, AIF1, AIM1, AMD1, ARG1, ATP6V1G2, ADGRB3, BAK1, BCKDHB, CFB, PI |
| 1p22.2 | 0.22813 | 2699 | None | chr1:1-248956422 | hsa-mir-664, hsa-mir-3119-1, hsa-mir-551a, ABCA4, ABL2, ACADM, ACTA1, ACTN2, ADAR, ADORA1, ADORA3, PARP1 |

**b) Focal Amplifications**

| Cytoband | Residual q Value | Genes in Peak | CRC Related | Wide Peak Boundaries | Genes in Peak |
|---|---|---|---|---|---|
| 6p22.2 | 1.8978E-06 | 2 | None | chr6:26285065-26377724 | HIST1H4H, BTN3A2 |
| 12p13.33 | 0.0013843 | 24 | None | chr12:1018844-2898391 | CACNA1C, FKBP4, FOXM1, TULP3, ERC1, ITFG2, ADIPOR2, WNT5B, RHNO1, NRIP2, CACNA2D4, FBXL14, DCP1B, LO |
| 16p11.2 | 0.0013843 | 26 | None | chr16:30081807-30479268 | SEPT1, ITGAL, PPP4C, MAPK3, SULT1A3, TBX6, CD2BP2, CORO1A, SEPHS2, TBC1D10B, MYLPF, ZNF771, SLX1B, DCTF |
| 1q21.1 | 0.0023775 | 9 | None | chr1:145909864-146019833 | ITGA10, PEX11B, RBM8A, TXNIP, POLR3GL, GNRHR2, LIX1L, HFE2, ANKRD34A |
| 19q13.33 | 0.0081553 | 6 | None | chr19:48614669-48670645 | CA11, DBP, RPL18, SPHK2, NTN5, SEC1P |
| 12p13.31 | 0.0096137 | 19 | None | chr12:6865885-6978788 | ATN1, ENO2, PTPN6, TPI1, USP5, LPCAT3, LRRC23, EMG1, PHB2, SPSB2, C12orf57, DSTNP2, RPL13P5, MIR141, MIR2 |
| 12p11.21 | 0.012916 | 6 | None | chr12:29774019-31001663 | IPO8, CAPRIN2, TMTC1, TSPAN11, LOC645485, LINC00941 |
| 7p15.2 | 0.01638 | 16 | None | chr7:27129126-27244617 | EVX1, HOXA4, HOXA5, HOXA6, HOXA7, HOXA9, HOXA10, HOXA11, HOXA13, HOXA11-AS, MIR196B, HOXA-AS3, HOTT |
| 8p11.21 | 0.017191 | 13 | None | chr8:41748910-42707147 | ANK1, CHRNB3, IKBKB, PLAT, POLB, SLC20A2, VDAC3, KAT6A, AP3M2, DKK4, SMIM19, LOC101929897, LOC105379 |
| 18q21.1 | 0.023373 | 4 | None | chr18:48753413-49266691 | SMAD7, CTIF, DYM, MIR4744 |
| 8q24.21 | 0.023807 | 24 | None | chr8:126547551-129758304 | MYC, POU5F1B, PVT1, GSDMC, CCDC26, FAM84B, TMEM75, CASC8, LINC00977, CASC11, MIR1205, MIR1206, MIR |
| 13q12.2 | 0.029928 | 12 | None | chr13:27271385-27986142 | CDX2, GTF3A, PDX1, POLR1D, MTIF3, GSX1, LNX2, RASL11A, ATP5EP2, URAD, LINC00543, PDX1-AS1 |
| 7p11.2 | 0.03441 | 12 | None | chr7:53035573-55375760 | EGFR, HPVC1, SEC61G, LANCL2, VSTM2A, POM121L12, VSTM2A-OT1, LINC01446, EGFR-AS1, LOC100996654, LINC |
| 11q12.3 | 0.03441 | 11 | None | chr11:62744667-62842408 | POLR2G, STX5, NXF1, TAF6L, WDR74, TMEM223, ZBTB3, TMEM179B, MIR6514, MIR6748, LOC105369332 |
| 1q21.3 | 0.035089 | 15 | None | chr1:153449472-153688794 | ILF2, NPR1, S100A1, S100A2, S100A3, S100A4, S100A5, S100A6, S100A7, S100A13, SNAPIN, CHTOP, S100A14, S100A |
| 11q13.1 | 0.035089 | 8 | None | chr11:64225400-64263481 | FKBP2, DNAJC4, PLCB3, VEGFB, PPP1R14B, TRPT1, NUDT22, LOC105369340 |
| 1q22 | 0.061091 | 7 | None | chr1:155078843-155187852 | EFNA1, EFNA3, MUC1, DPM3, SLC50A1, TRIM46, KRTCAP2 |
| 17q21.32 | 0.072747 | 18 | None | chr17:48521027-48778545 | HOXB1, HOXB2, HOXB3, HOXB4, HOXB5, HOXB6, HOXB7, HOXB8, HOXB9, HOXB13, PRAC1, TTLL6, PRAC2, HOXB-A |
| 12q13.13 | 0.088166 | 3 | None | chr12:52901060-53024355 | EIF4B, KRT8, KRT18 |
| 6p21.1 | 0.093298 | 71 | None | chr6:41333272-44228159 | BYSL, CCND3, SLC29A1, GUCA1A, GUCA1B, MDFI, MEA1, PEX6, PGC, POLH, PPP2R5D, PTK7, PRPH2, SRF, TBCC, VE |
| 19q13.32 | 0.093298 | 18 | None | chr19:45388678-45707586 | ERCC1, FOSB, GIPR, GPR4, RTN2, SNRPD2, VASP, PPP1R13L, CD3EAP, EML2, QPCTL, OPA3, PPM1N, MIR330, MIR64 |
| 19q13.2 | 0.10832 | 9 | None | chr19:38632550-38877459 | ACTN4, ECH1, HNRNPL, LGALS4, LGALS7, EIF3K, RINL, CAPN12, LGALS7B |
| 1p34.1 | 0.20333 | 11 | None | chr1:44757315-44831590 | PLK3, RPS8, PTCH2, KIF2C, SNORD55, SNORD46, SNORD38A, SNORD38B, BTBD19, BEST4, TCTEX1D4 |
| 19q13.12 | 0.22933 | 11 | None | chr19:35056890-35271240 | HPN, FXYD1, FXYD3, USF2, LSR, FXYD7, FXYD5, FAM187B, LGI4, HPN-AS1, MIR6887 |
| 11p13 | 0.24675 | 24 | None | chr11:34501751-40124261 | CD44, ELF5, RAG1, RAG2, SLC1A2, TRAF6, PDHX, FJX1, PAMR1, EHF, COMMD9, APIP, TRIM44, LRRC4C, PRR5L, C11o |

**a)**

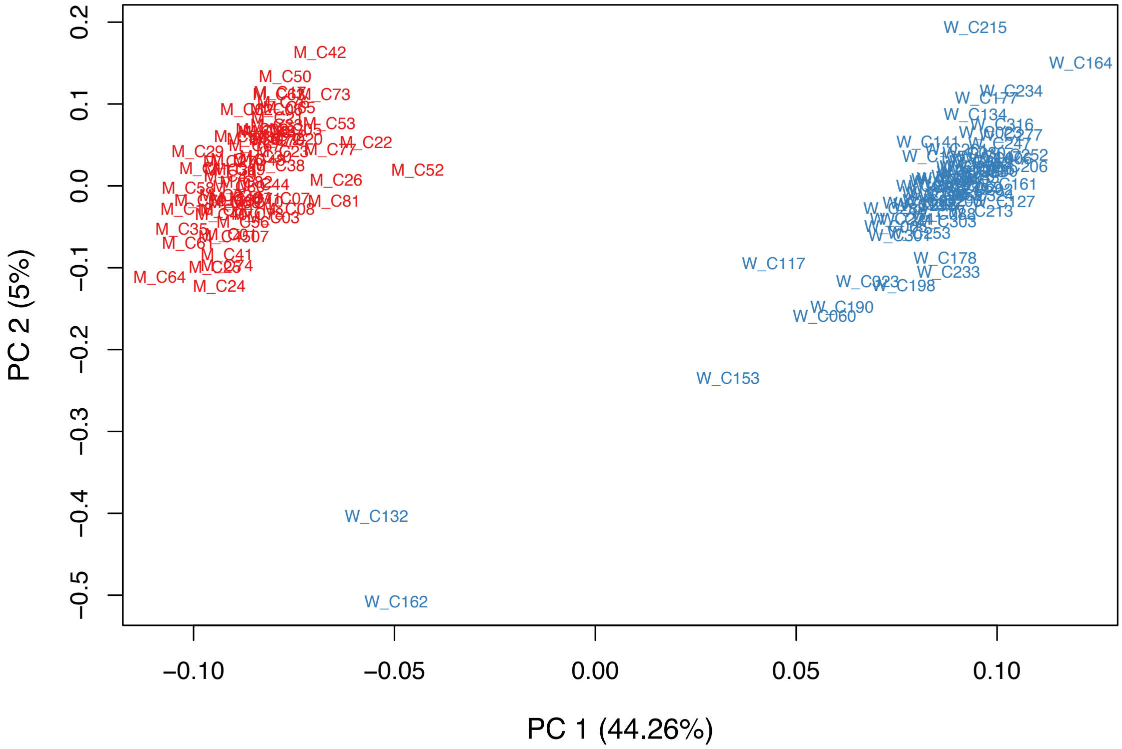


**b)**

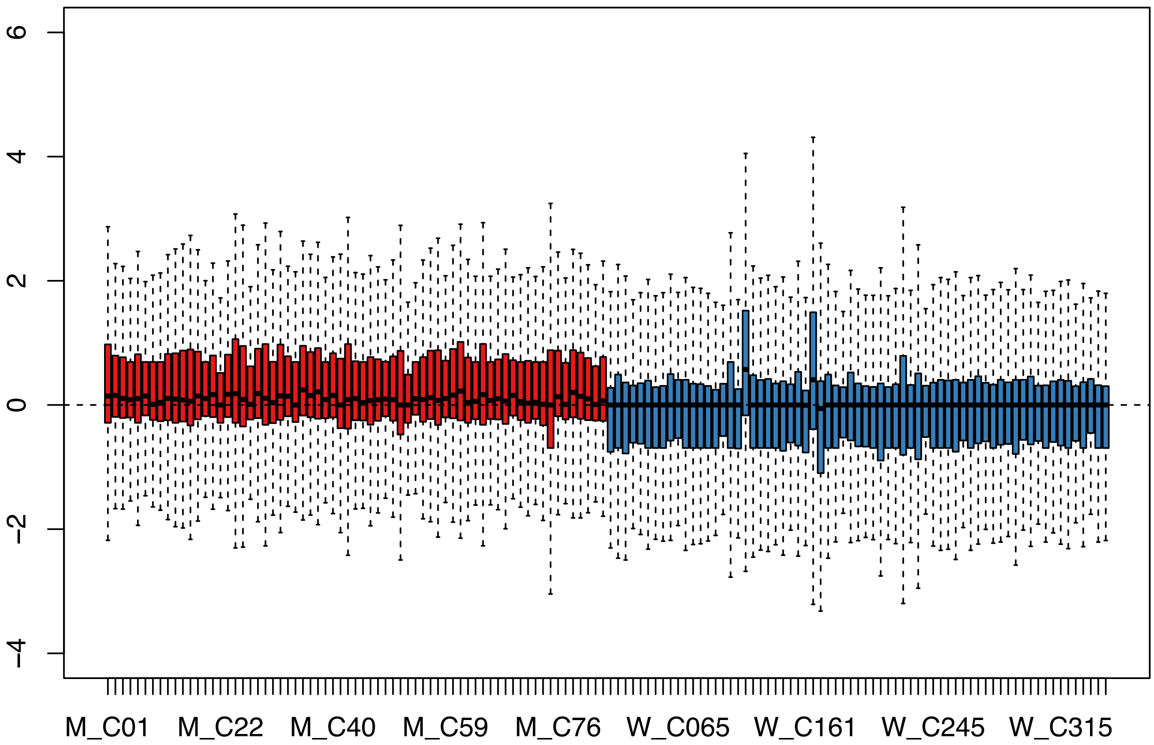


**Supplementary Figure 8: Differential gene expression (DGE) analysis among CRC tumors from 67 Hispanic/Latino patients and 67 Non-Hispanic White patients.** DGE analysis identified unique gene expression patterns in our Hispanic/Latino (H/L) CRC cohort compared to Non-Hispanic White (NHW) CRC cases from two publicly available databases: TCGA-COAD and TCGA-READ. **a) Principal component analysis (PCA) plot.** The plot displays the distribution of these samples along two principal components, PC1 and PC2, which capture the majority of the variance in the genetic data between Hispanic Latino (red) and non-Hispanic white (blue). **b) Relative Log Expression (RLE) plot.** This diagnostic plot was to assess the quality and normalization of gene expression data. It was particularly useful for ensuring the consistency of expression levels across Hispanic/Latino patients (red) and Non-Hispanic White samples (blue).

**Supplementary Table 11: Patient Demographics and Clinical Characteristics of 67 Non-Hispanic Whites (NHW) records.** This table provides information on patient demographics and clinical characteristics, including age at diagnosis, gender, tumor location, and pathological stage. The data is compiled from individual records obtained from public databases projects TCGA-COAD and TCGA-READ.

| Characteristic | TCGA NHW Samples (n = 67)<br>*n (%)* |
|---|---|
| **Age at Diagnosis** | |
| < 50 years | 30 (44.8%) |
| >50 years | 37 (55.2%) |
| **Gender** | |
| Male | 36 (53.7%) |
| Female | 31 (46.3%) |
| **Tumor Location** | |
| Colon | 43 (64.2%) |
| Rectum | 24 (35.8%) |
| **Pathological Stage** | |
| Early (Stage I or II) | 25 (37.3%) |
| Late (Stage III or IV) | 38 (5.97%) |
| No TNM Applicable/Unknown | 4 (28.4%) |

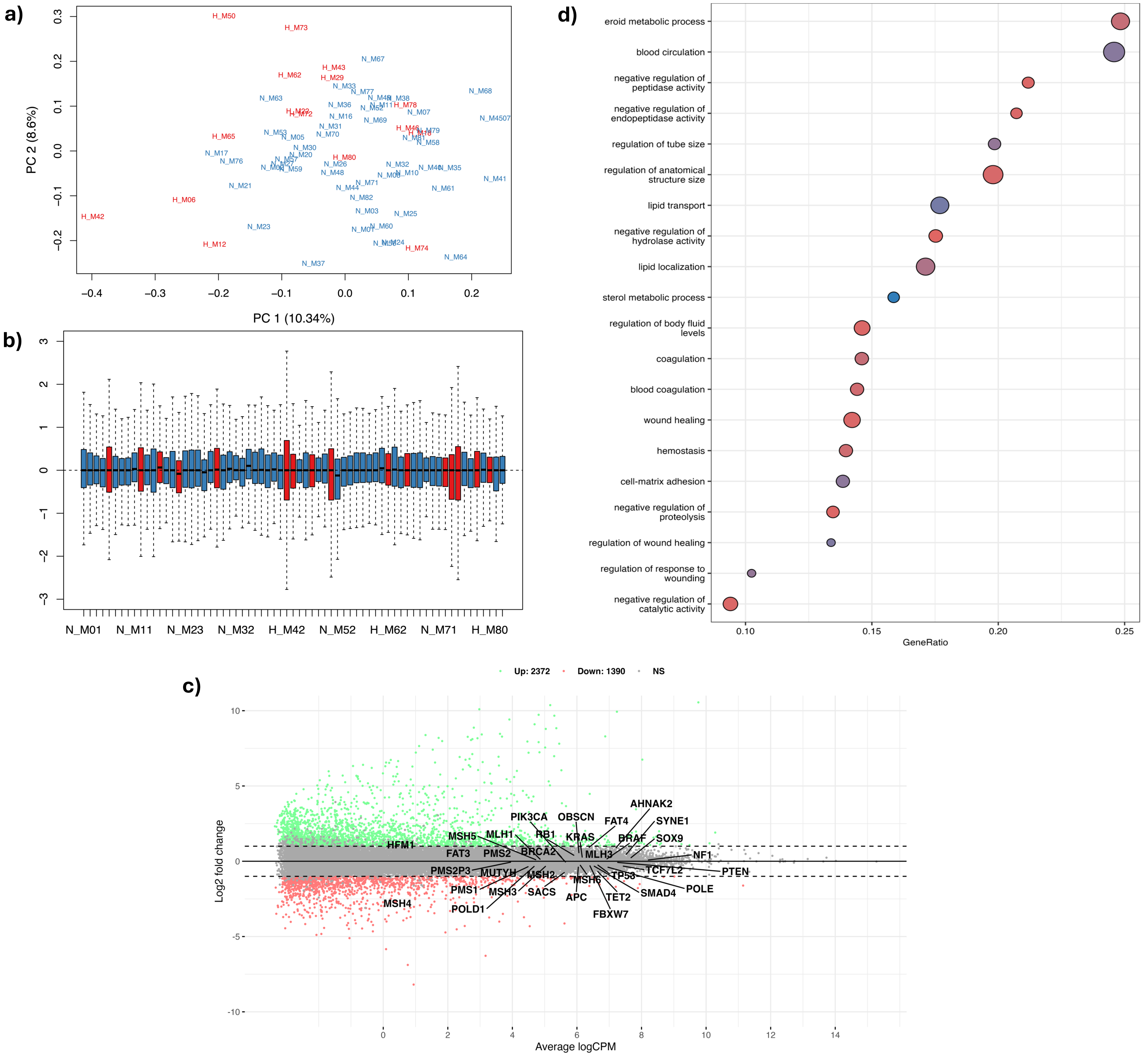


**Supplementary Figure 9: Differential Gene Expression (DGE) Analysis Among CRC Hypermutated and Non-Hypermutated Tumors in Hispanic/Latino Patients.** This figure presents a detailed differential gene expression (DGE) analysis between hypermutated and non-hypermutated colorectal cancer (CRC) tumors in our cohort of Hispanic/Latino (H/L) patients, highlighting unique gene expression patterns in these two groups. **a)** Principal Component Analysis (PCA) Plot: This plot shows the distribution of samples along two principal components, PC1 and PC2, capturing the majority of variance in the genetic data. Hypermutated samples are represented in red, while non-hypermutated samples are in blue. **b)** Relative Log Expression (RLE) Plot: This diagnostic plot assesses the quality and normalization of gene expression data, ensuring consistency of expression levels across hypermutated (red) and non-hypermutated (blue) samples. **c)** Mean-Average (MA) Plot: This scatter plot depicts the upregulated and downregulated CRC-related genes based on log2 fold change, illustrating the differential expression between the two groups. **d)** Pathway Analysis: This graph presents distinct cellular pathways identified through pathway analysis by comparing hypermutated and non-hypermutated CRC tumors in Hispanic/Latino patients. It displays the names of the pathways, adjusted p-values, and the number of genes altered in each pathway.

**Supplementary Table 12: Differentially Expressed CRC-Related Genes in Hypermutated and Non-Hypermutated Tumors from Hispanic/Latino Patients.** It highlights the specific genes that exhibit significant differences in expression levels between these two groups.

| Gene | logFC | logCPM | LR | p-value |
|---|---|---|---|---|
| MSH4 | -2.16 | 1.03 | 84.72 | 3.4314E-20 |
| TET2 | -0.57 | 6.60 | 25.02 | 5.6636E-07 |
| SACS | -0.65 | 5.67 | 15.05 | 1.0466E-04 |
| FAT4 | 0.83 | 6.32 | 13.79 | 2.0458E-04 |
| PMS1 | -0.28 | 4.57 | 5.74 | 0.016590 |
| AHNAK2 | 0.79 | 7.17 | 5.56 | 0.018360 |
| RB1 | 0.31 | 5.97 | 5.06 | 0.024463 |
| MSH2 | -0.38 | 5.32 | 4.79 | 0.028548 |
| OBSCN | 0.45 | 6.07 | 4.74 | 0.029478 |
| APC | -0.27 | 6.05 | 4.67 | 0.030707 |
| POLE | -0.34 | 6.87 | 4.64 | 0.031235 |
| FAT3 | 0.64 | 2.90 | 4.36 | 0.036876 |
| TCF7L2 | -0.29 | 7.33 | 4.33 | 0.037424 |

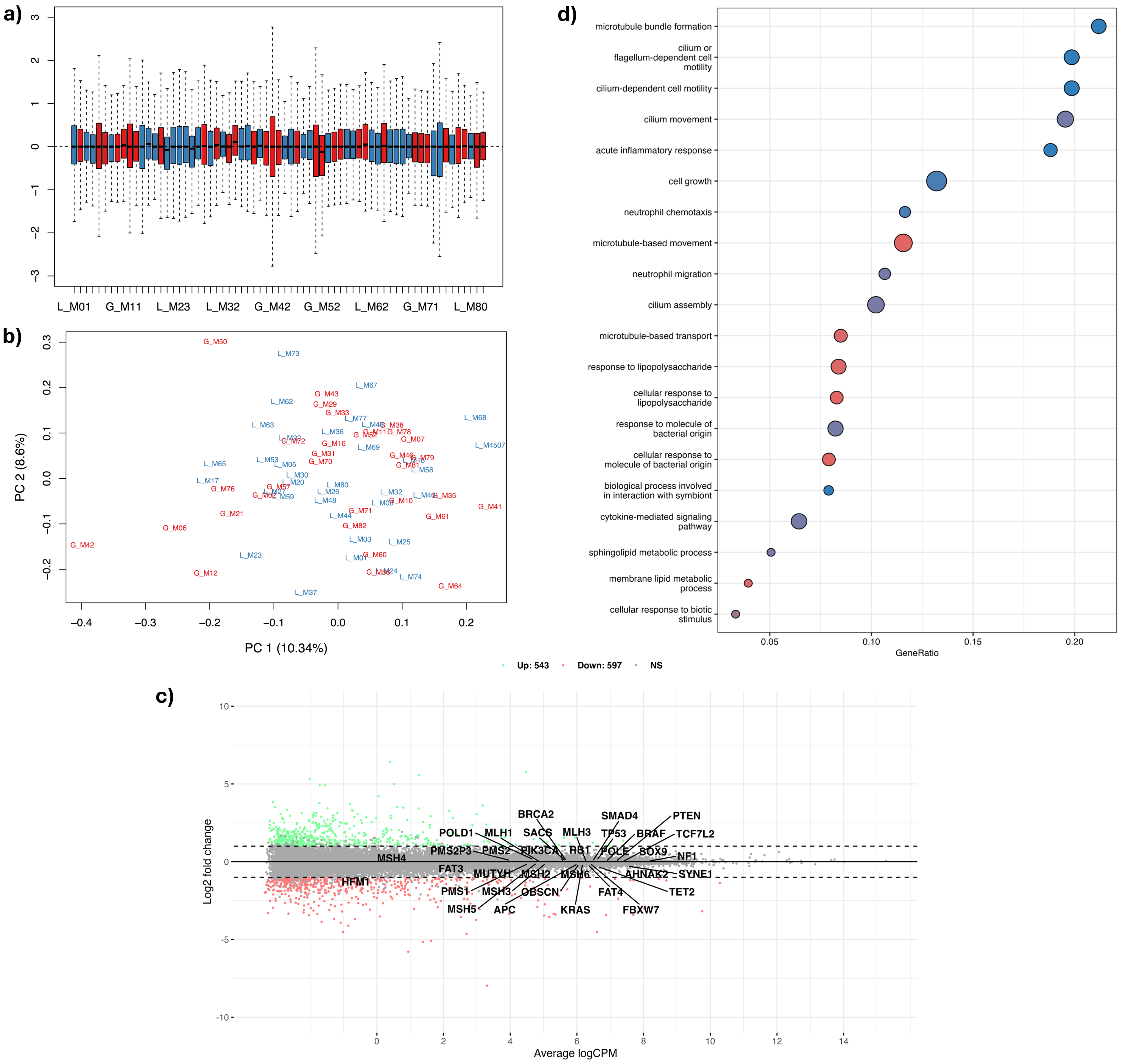


**Supplementary Figure 10: Differential Gene Expression (DGE) Analysis Among CRC Tumors with Varying 1KG-PEL-like Proportions in Hispanic/Latino Patients.** This figure presents a comprehensive differential gene expression analysis among colorectal cancer (CRC) tumors from Hispanic/Latino patients, categorized by 1000 Genomes Project Peruvian-in-Lima-like (1KG-PEL-like) proportions. The analysis compares samples with 1KG-PEL-like proportions >55% and those with 1KG-PEL-like proportions ≤55%. **a)** Principal Component Analysis (PCA) Plot: This plot displays the distribution of samples along two principal components, PC1 and PC2, which capture the majority of the variance in the genetic data. Samples with 1KG-PEL-like >55% are shown in red, while samples with 1KG-PEL-like ≤55% are shown in blue. **b)** Relative Log Expression (RLE) Plot: This diagnostic plot assesses the quality and normalization of gene expression data, ensuring consistency of expression levels across samples with 1KG-PEL-like >55% (red) and 1KG-PEL-like ≤55% (blue). **c)** Mean-Average (MA) Plot: This scatter plot visualizes the upregulated and downregulated CRC-related genes based on log2 fold change. **d)** Pathway Analysis: This graph presents the distinct cellular pathways identified through pathway analysis by comparing CRC tumors with 1KG-PEL-like proportions >55% and 1KG-PEL-like proportions ≤55%. The graph displays the names of the pathways, adjusted

**Supplementary Table 13: Differentially Expressed CRC-Related Genes among CRC Tumors with Varying 1KG-PEL-like Proportions in Hispanic/Latino Patients.** It highlights the specific genes that exhibit significant differences in expression levels between samples with 1KG-PEL-like proportions >55% and those with 1KG-PEL-like proportions ≤55%.

| Gene | logFC | logCPM | LR | p-value |
|---|---|---|---|---|
| TET2 | -0.29 | 6.60 | 6.09 | 0.013612 |
| HFM1 | -0.66 | -0.04 | 5.77 | 0.016315 |
| AHNAK2 | -0.43 | 7.17 | 2.38 | 0.123152 |
| MSH6 | -0.17 | 6.07 | 2.11 | 0.146291 |
| SYNE1 | -0.27 | 7.48 | 1.90 | 0.168190 |
| MSH4 | -0.40 | 1.03 | 1.82 | 0.177668 |
| MSH2 | -0.19 | 5.32 | 1.50 | 0.220839 |
| RB1 | 0.13 | 5.97 | 1.14 | 0.285359 |
| MUTYH | -0.12 | 4.12 | 1.12 | 0.290929 |
| APC | -0.11 | 6.05 | 0.81 | 0.368395 |

**Supplementary Table 14: Gene Fusions.** This table presents the gene fusions identified in 67 Hispanic/Latino colorectal cancer (CRC) patients. The data includes the locations of gene fusions and recurrent deletion peaks identified from somatic copy number alterations (SCNAs) in selected CRC-related genes. Specific details provided are the upstream gene, Ensembl ID for the upstream gene, upstream breakpoint, downstream gene, Ensembl ID for the downstream gene, and downstream breakpoint.

| Gene Upstream | Ensembl Upstream | Breakpoint Upstream | Gene Downstream | Ensembl Downstream | Breakpoint Downstream |
|---|---|---|---|---|---|
| **EML4** | **ENSG00000143924** | **42295516** | **ALK** | **ENSG00000171094** | **29223528** |
| **ERLIN2** | **ENSG00000147475** | **37741818** | **FGFR1** | **ENSG00000077782** | **38457534** |
| **PTPRK** | **ENSG00000152894** | **128520259** | **RSPO3** | **ENSG00000146374** | **127148648** |
| **RAF1** | **ENSG00000132155** | **12590798** | **TMEM40** | **ENSG00000088726** | **12749840** |
| PMS2P9 | ENSG00000233448 | 77043778 | AC105052.5 | ENSG00000286830 | 102703481 |
| AC108865.1 | ENSG00000250829 | 186891368 | AC110772.2 | ENSG00000250971 | 187019821 |
| GNAS | ENSG00000087460 | 58895684 | BCAS1 | ENSG00000064787 | 54058723 |
| PMS2P11 | ENSG00000241350 | 77015855 | CCDC146 | ENSG00000135205 | 77167658 |
| PMS2P9 | ENSG00000233448 | 77043778 | CCDC146 | ENSG00000135205 | 77167658 |
| PPP1R13B | ENSG00000088808 | 103753000 | EML1 | ENSG00000066629 | 99850853 |
| PFKFB3 | ENSG00000170525 | 6226365 | LINC02649 | ENSG00000215244 | 6326546 |
| CLPB | ENSG00000162129 | 72380281 | MYEOV | ENSG00000172927 | 69341510 |
| CDH17 | ENSG00000079112 | 94145928 | RUNX1T1 | ENSG00000079102 | 92017363 |
| FBXO25 | ENSG00000147364 | 435707 | SEPTIN14 | ENSG00000154997 | 55796092 |
| OR51S1 | ENSG00000176922 | 4848669 | TP53I11 | ENSG00000175274 | 44933050 |
| DST | ENSG00000151914 | 56603564 | ABHD16A | ENSG00000204427 | 31701028 |
| ELL | ENSG00000105656 | 18521921 | AC011447.3 | ENSG00000267383 | 20240637 |
| PSD3 | ENSG00000156011 | 18936034 | AC100849.1 | ENSG00000253557 | 19085078 |
| AC127502.3 | ENSG00000283345 | 30513698 | AC127502.1 | ENSG00000215302 | 30474356 |
| AL611929.1 | ENSG00000234768 | 168194688 | AFDN | ENSG00000130396 | 167943129 |
| WARS2 | ENSG00000116874 | 119076350 | AGBL4 | ENSG00000186094 | 49045800 |
| UHRF1BP1L | ENSG00000111647 | 100088915 | ANKS1B | ENSG00000185046 | 99779972 |
| SMAD4 | ENSG00000141646 | 51030623 | ANO4 | ENSG00000151572 | 100901646 |
| EHBP1 | ENSG00000115504 | 62942896 | ANXA4 | ENSG00000196975 | 69652998 |
| VSTM5 | ENSG00000214376 | 93850412 | AP003066.1 | ENSG00000254587 | 96975663 |
| ASH1L | ENSG00000116539 | 155395459 | ARID4B | ENSG00000054267 | 235182793 |
| IGH-@-ext | IGH- | 105588395 | ARL4D | ENSG00000175906 | 43400325 |
| ERC1 | ENSG00000082805 | 1028572 | CACNA1C | ENSG00000151067 | 2448976 |
| SAP30BP | ENSG00000161526 | 75671863 | CAVIN1 | ENSG00000177469 | 42405388 |
| PMS2P6 | ENSG00000174384 | 73097567 | CCDC146 | ENSG00000135205 | 77167658 |
| POGZ | ENSG00000143442 | 151459152 | CDC42SE1 | ENSG00000197622 | 151059773 |
| TANC1 | ENSG00000115183 | 159065971 | DAPL1 | ENSG00000163331 | 158804282 |
| GALK2 | ENSG00000156958 | 49235941 | DENND2A | ENSG00000146966 | 140602542 |
| TMED5 | ENSG00000117500 | 93180054 | DHCR24 | ENSG00000116133 | 54854234 |
| SEC61A2 | ENSG00000065665 | 12129794 | DHTKD1 | ENSG00000181192 | 12117673 |
| MAP2K2 | ENSG00000126934 | 4117419 | EBI3 | ENSG00000105246 | 4236936 |
| VPS41 | ENSG00000006715 | 38817817 | ELMO1 | ENSG00000155849 | 37342763 |
| MSI2 | ENSG00000153944 | 57529724 | GDPD1 | ENSG00000153982 | 59234492 |
| VAV3 | ENSG00000134215 | 107683488 | HMCN1 | ENSG00000143341 | 185965802 |
| MAN2B1 | ENSG00000104774 | 12658063 | HNRNPM | ENSG00000099783 | 8483158 |
| VGLL4 | ENSG00000144560 | 11720394 | HRH1 | ENSG00000196639 | 11259003 |
| PPFIA1 | ENSG00000131626 | 70272436 | IGH@-ext | IGH | 106373964 |
| GINS1 | ENSG00000101003 | 25413854 | ILDR2 | ENSG00000143195 | 166958101 |
| MARK2 | ENSG00000072518 | 63839560 | LNCAROD | ENSG00000231131 | 52561115 |
| DUSP16 | ENSG00000111266 | 12519862 | MANSC1 | ENSG00000111261 | 12330958 |
| AUTS2 | ENSG00000158321 | 70698620 | MDGA2 | ENSG00000139915 | 46882221 |
| NPEPPS | ENSG00000141279 | 47531555 | MED1 | ENSG00000125686 | 39451290 |
| STARD13 | ENSG00000133121 | 33285470 | MTMR6 | ENSG00000139505 | 25274187 |
| CAPRIN1 | ENSG00000135387 | 34086413 | PAMR1 | ENSG00000149090 | 35474744 |
| MCM3AP | ENSG00000160294 | 46236829 | PCBP3 | ENSG00000183570 | 45744231 |
| DHX9 | ENSG00000135829 | 182856578 | PDC-AS1 | ENSG00000229739 | 186519776 |
| CTIF | ENSG00000134030 | 48539312 | RAB31 | ENSG00000168461 | 9775278 |
| WWC1 | ENSG00000113645 | 168431444 | REEP5 | ENSG00000129625 | 112887183 |
| PAK1 | ENSG00000149269 | 77473552 | RIMKLA | ENSG00000177181 | 42409984 |
| AL669831.4 | ENSG00000230092 | 805799 | SEPTIN14 | ENSG00000154997 | 55796092 |
| AL391840.3 | ENSG00000287811 | 79538843 | SH3BGRL2 | ENSG00000198478 | 79673614 |
| TRIM37 | ENSG00000108395 | 59106441 | SKA2 | ENSG00000182628 | 59131367 |
| SPATA13 | ENSG00000182957 | 24160932 | SLC46A3 | ENSG00000139508 | 28718022 |
| MDGA2 | ENSG00000139915 | 46877489 | SNX13 | ENSG00000071189 | 17875790 |
| RAB11FIP3 | ENSG00000090565 | 510800 | SPN | ENSG00000197471 | 29670734 |
| REEP5 | ENSG00000129625 | 112921163 | SSBP2 | ENSG00000145687 | 81461103 |
| C2CD5 | ENSG00000111731 | 22453896 | ST8SIA1 | ENSG00000111728 | 22070625 |
| CTTN | ENSG00000085733 | 70425401 | STARD10 | ENSG00000214530 | 72759381 |
| LDLR | ENSG00000130164 | 11089615 | TMPRSS9 | ENSG00000178297 | 2389761 |
| USP32 | ENSG00000170832 | 60219670 | USH2A | ENSG00000042781 | 216097213 |
| DNAJC1 | ENSG00000136770 | 22003213 | VKORC1 | ENSG00000167397 | 31091342 |
| CLTC | ENSG00000141367 | 59660588 | VMP1 | ENSG00000062716 | 59808796 |
